# Large-scale clustering of longitudinal faecal calprotectin and C-reactive protein profiles in inflammatory bowel disease

**DOI:** 10.1101/2024.11.08.24316916

**Authors:** Nathan Constantine-Cooke, Marie Vibeke Vestergaard, Nikolas Plevris, Karla Monterrubio-Gómez, Clara Ramos Belinchón, Solomon Ong, Alexander T. Elford, Beatriz Gros, Aleksejs Sazonovs, Gareth-Rhys Jones, Tine Jess, Catalina A. Vallejos, Charlie W. Lees

**Affiliations:** Centre for Genomic and Experimental Medicine, Institute of Genetics and Cancer, University of Edinburgh, Edinburgh, UK; MRC Human Genetics Unit, Institute of Genetics and Cancer, University of Edinburgh, Edinburgh, UK; Center for Molecular Prediction of Inflammatory Bowel Disease, PREDICT, Department of Clinical Medicine, Aalborg University, Copenhagen, Denmark; Edinburgh IBD Unit, Western General Hospital, Edinburgh, UK; Faculty of Medicine, The University of Melbourne, Melbourne, Australia; Reina Sofía University Hospital, Gastroenterology and Hepatology. IMIBIC. University of Cordoba. Cordoba, Spain; Centre for Inflammation Research, The Queen’s Medical Research Institute, University of Edinburgh, Edinburgh, UK; Department of Gastroenterology & Hepatology, Aalborg University Hospital, Aalborg, Denmark; Health Data Research UK, London, England, UK

## Abstract

**Background:** Crohn’s disease (CD) and ulcerative colitis (UC) are highly heterogeneous, dynamic and unpredictable, with a marked disconnect between symptoms and intestinal inflammation.

**Objective:** We aimed to describe latent disease heterogeneity in IBD by identifying longitudinal faecal calprotectin (FC) and CRP patterns.

**Design:** In this longitudinal study, patient-level measurements of FC and CRP in two European cohorts were modelled. Latent class mixed models were used to cluster individuals with similar longitudinal profiles. Associations between cluster assignment and baseline characteristics were quantified using multinomial logistic regression. Differences in advanced therapy use across clusters were explored. Finally, we considered the overlap between FC and CRP clusters.

**Results:** We included 1036 patients in the FC discovery analysis (Lothian) with a total of 10545 FC observations (median 9 per subject, IQR 6–13), and 7880 patients in the replication (Denmark). The CRP discovery analysis consisted of 1838 patients with 49364 measurements (median 20 per subject; IQR 10–36), with 10041 patients in the replication cohort. Eight distinct clusters of inflammatory behaviour over time were identified in the FC and CRP analysis. Similar patterns were observed in the replication cohort. The clusters included rapid remitters, delayed remitters, remitting-relapsing disease, and non-remitters The highest use of early advanced therapy was in the group with the lowest overall inflammation. There was broadly poor agreement between FC and CRP cluster assignment.

**Conclusion:** Distinct patterns of inflammatory behaviour over time are evident in patients with IBD. These data pave the way for a deeper understanding of disease heterogeneity in IBD.

## Introduction

Inflammatory bowel disease (IBD), an umbrella term for Crohn’s disease (CD), ulcerative colitis (UC) and inflammatory bowel disease unclassified (IBDU), has a prevalence of almost 1% in the UK.^1,2^ IBD is characterised by chronic relapsing and remitting inflammation of the gastrointestinal (GI) tract, with debilitating symptoms, negatively impacting quality of life.^3,4^ Studies have demonstrated uncontrolled GI tract inflammation increases the risk of adverse outcomes including colorectal cancer,^5^ stricturing/penetrating complications, and surgery.^6,7^ However, IBD is highly heterogeneous with respect to symptoms, inflammatory burden, treatment response, and long-term outcomes.

Current IBD classification methods are mostly based on historic nomenclature which utilise baseline phenotypic characteristics and do not consider the dynamic and unpredictable nature of the disease. Similarly, attempts at developing prediction tools to identify high risk patients have focused on static clinical parameters^8^ and do not include the influence of other factors, such as advanced therapy timing. In the wake of the increasing prevalence of IBD and associated healthcare burden,^9^ new methods to characterise the dynamic disease course and help identify at-risk individuals who require aggressive therapy with close monitoring versus those that need less intense input are essential.

The original IBSEN studies provided the first data on the clinical course of patients with UC and CD during the first 10-years of diagnosis.^10,11^ However, these data were predicated on predefined disease patterns, rather than being data driven, utilising symptoms alone. It is now widely accepted there is a clear disconnect between inflammation and symptoms in IBD.^12^ It is therefore imperative characterisations of disease course include objective parameters of inflammation.

C-reactive protein (CRP) and faecal calprotectin (FC) are well established tools for monitoring patients with IBD, but are typically interpreted in terms of the most recent measurement or short-term trends. Interrogation of long-term trends of inflammation could greatly assist clinical decision making and improve prediction of future events. Moreover, modelling inflammatory behaviour over time might be a key tool for characterising the largely unexplained heterogeneity seen in IBD, and provide new tools for disease sub-phenotyping beyond the current Montreal classification.^13^

In this study, we aimed to 1) identify groups of IBD patients with similar longitudinal patterns of inflammation, 2) determine if these groupings were associated with age, sex, IBD type, Montreal classification, or therapy, and 3) explore whether subjects with similar longitudinal FC profiles also share similar CRP profiles. To assess the robustness of our findings, we performed the analysis separately in two population-level cohorts, one covering the NHS Lothian health board in Scotland and one national-level registry in Denmark.

## Methods

### Study design

This was a cohort study carried out separately in two population-level cohorts, based in Scotland (discovery) and Denmark (validation). Using population-level cohorts reduces potential biases due to recruitment.^14^ Detailed definitions for each cohort are provided in Supplementary Note 1.

In Scotland, patients with a confirmed diagnosis of IBD (Lennard-Jones criteria)^15^ at any age were followed up for seven years from diagnosis. Baseline phenotype data (sex, age at diagnosis, IBD type, date of diagnosis) were obtained from the Lothian IBD registry (LIBDR), a retrospective cohort of patients receiving IBD care from the NHS Lothian health board. The LIBDR is estimated to cover 94.3% of all IBD cases in the area.^1^ Additional phenotyping was extracted by the clinical team from electronic health records (TrakCare; InterSystems, Cambridge, MA). This includes smoking and Montreal classification for disease location, behaviour and extent, all recorded at the time of diagnosis. Data on secondary care prescribing of all advanced therapies, including start/stop dates, were extracted from TrakCare and NHS Lothian pharmacy databases. Primary care prescribing data were not available.

In Denmark, IBD cases were identified from national health registries as in Agrawal et al.^16^ Briefly, patients were required to have at least two IBD-related hospital interactions within a two-year period. As biochemistry data were only nationwide from 2015, a follow-up of five years since diagnosis was used. Montreal classification and smoking status were not available for the Danish cohort. However, this cohort had data on all redeemed primary care prescriptions for corticosteroids and immunosuppressants, in addition to secondary care advanced therapy prescribing.

### Inclusion/exclusion criteria

Figure S1 summarises the inclusion/exclusion criteria for diagnosis dates. Further exclusions were then separately applied based on the availability of longitudinal FC and CRP measurements. Subjects were required to have a diagnostic measurement (± three months of diagnosis, Figures S2, S3) and have a further two measurements within the cohort-specific longitudinal follow up. For FC, only measurements within the limits of detection of the assays were considered for this (see the *Longitudinal measurements and preprocessing* section). For CRP, this filtering was applied after preprocessing and only in the LIBDR; individuals for which all measurements were outside the limit of detection were excluded. If any measurements were observed within three months prior to the recorded diagnosis date, it was assumed there was a delay for the diagnosis record.^17^ In such cases, measurements were realigned (Figure S4).

**Figure 1.**
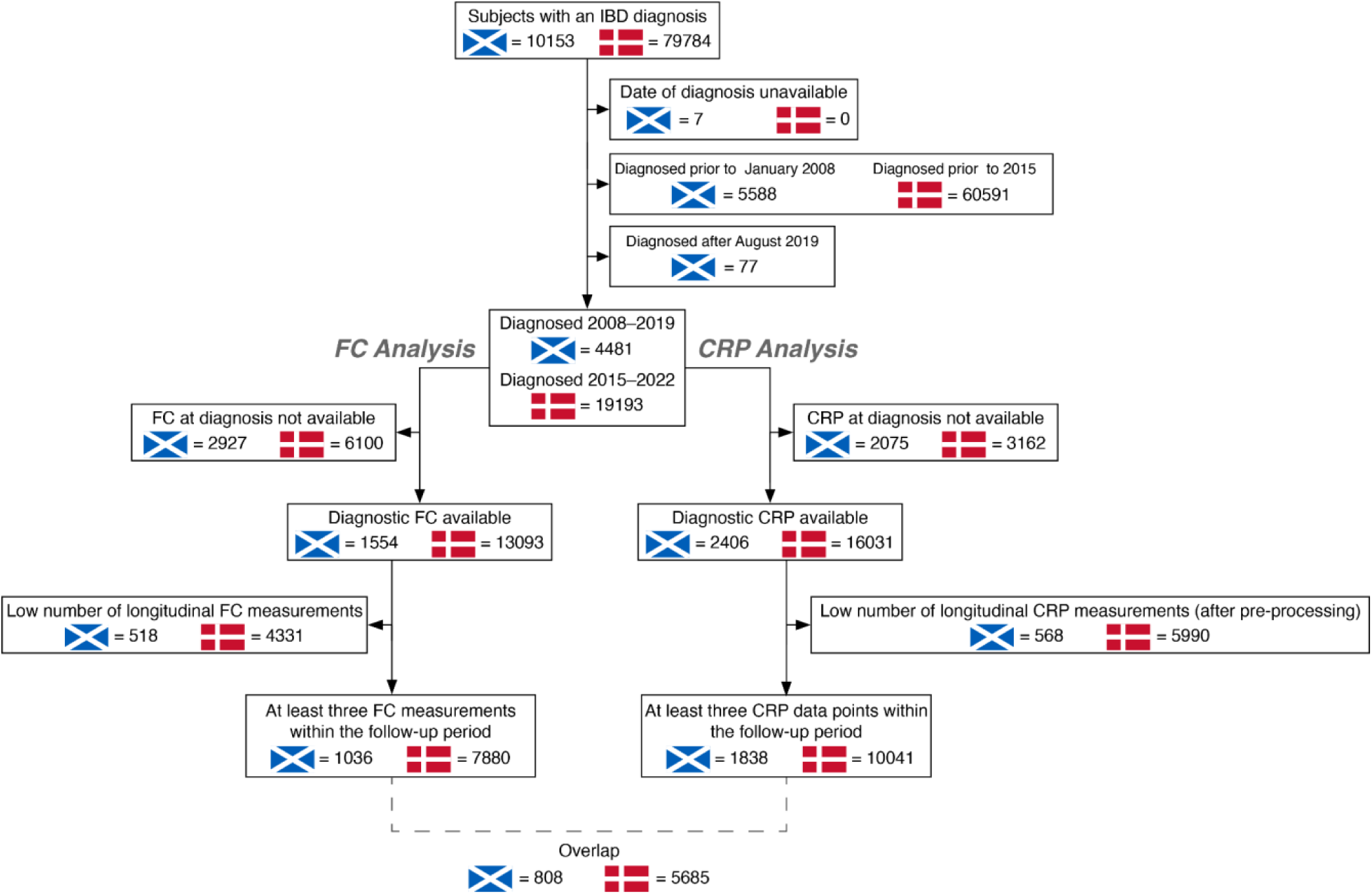
Derivation of the study cohorts based on separate faecal calprotectin (FC) and C-reactive protein (CRP) inclusion/exclusion criteria. A 7-year follow-up period was used for LIBDR, a 5-year follow-up period was used for the Danish cohort. For the LIBDR analysis, patients with a diagnosis date prior to April 2008 were excluded as FC was was not routinely used to monitor disease activity in that period. For the Danish analysis, patients required a diagnosis in 2015 or later as this is when nationwide data are available from. Of the 2927 subjects in the LIBDR excluded due to lacking a diagnostic FC measurement, 659 subjects had a date of diagnosis recorded as January 1st. This largely corresponds to individuals for which an exact diagnosis was unknown (see Supplementary Note 1.1). The same occurred for 606 subjects out of 3162 excluded due to lack of a diagnostic CRP measurement in the LIBDR cohort. For FC, we included individuals with at least 3 measurements recorded to be within the limits of detection of the assay (<20 or >1250 for LIBDR, >1800 for Denmark). For CRP, individuals were excluded if they had less than 3 longitudinal measurements after pre-processing (see Supplementary Note 2.1). For the LIBDR only, subjects were excluded if all CRP measurements (after pre-processing) were outside the detection threshold (<1).

### Statistical analysis

#### Cohort description

Continuous variables were summarised as their median and interquartile range (IQR). Categorical variables were summarised as counts and percentages.

#### Longitudinal measurements and preprocessing

All observations were log-transformed. The availability of longitudinal measurements is summarised in Figure S1.

For LIBDR individuals, all measurements within seven years from diagnosis were considered. Failed tests, for example due to contamination, were discarded. All FC tests were performed from stool samples using the same ELISA technology.^18^ CRP was measured from blood samples. Due to limits of detection, observations <20 μg/g or >1250 μg/g were mapped as 20 μg/g and 1250 μg/g, respectively. Similarly, CRP observations for which only an upper bound was available, e.g. <1 mg/L, were mapped to the corresponding upper bound. For the Danish cohort, all measurements within five years from the date of diagnosis were considered. For FC and CRP, observations >1800μg/g and <4mg/L were mapped to the corresponding threshold, respectively.

To smooth out short-term fluctuations that do not reflect IBD behaviour (e.g. due to infection), CRP measurements were further processed by considering median values within fixed time intervals, after log transformation (Supplementary Note 2.1).

#### Longitudinal biomarker clustering

The Scottish and Danish data were analysed separately. FC and CRP trajectories were modelled independently using latent class mixed models (LCMMs),^18^ which can identify clusters of individuals with similar longitudinal patterns. LCMM includes two submodels (Supplementary Note 2.2). The longitudinal submodel captures temporal trends, with natural cubic splines as fixed effects and a patient-level random intercept. The cluster assignment submodel used IBD type as a covariate..

As the number of clusters is not known a priori, the optimal model was found using a grid search approach, considering 2–10 clusters. This considered Akaike information criterion (AIC)^19^, Bayesian information criterion (BIC), and visual inspection of cluster trajectories (Supplementary Note 2.3). The optimal number of clusters was chosen based on the LIBDR dataset (discovery), as it has more precise phenotyping (date of diagnosis and IBD type). The chosen number of clusters was then applied when analysing the Danish cohort (validation).

For each subject, LCMM estimates the probability of being assigned to each cluster. These estimates are static, based on all the longitudinal measurements available for a subject. Each individual was assigned to the cluster with the highest probability. The distribution of cluster assignment probabilities was used to assess uncertainty in these allocations with respect to follow-up length. This was defined as the time difference between diagnosis and the last available biomarker measurement (FC or CRP, depending on the analysis). For each cluster, the average of individual-specific probabilities of cluster assignment were reported.

To avoid displaying potentially identifiable individual-level data, exemplar trajectories were visualised as aggregated trends, summarised as the median across six randomly selected individuals. For the LIBDR data, clusters were ordered by the area under the biomarker trend inferred for each cluster, used as a proxy for cumulative inflammatory burden (Supplementary Note 2.4). The clusters found in the Danish cohort were ordered manually to resemble similar trajectories.

#### Associations with respect to cluster assignments

To facilitate the interpretation of each cluster, associations between cluster assignments and patient phenotypes were considered. Violin plots and bar plots were used as a visual summary when considering continuous (age) and discrete (sex, IBD type and, for LIBDR, additional phenotyping) covariates, respectively.

Multinomial logistic regression was used to quantify these associations using patient phenotypes as covariates. Multivariate models were considered, and the associated 95% confidence intervals are reported. Individuals with missing covariate values were excluded when fitting each model. As a sensitivity analysis, the analysis was repeated after excluding individuals with a low probability for cluster assignment (<0.5).

#### Prescribing trends

To compare patterns of advanced therapy (AT) across clusters, the cumulative distribution of first-line advanced therapy use was calculated for both the Scottish and Danish cohorts. As primary care prescribing data were available for Danish subjects, patterns for systemic corticosteroid and immunomodulator use were also investigated. Results are reported stratified by IBD type (CD and UC only).

#### Comparison between FC and CRP cluster assignments

For subjects meeting the criteria for both FC and CRP analyses, the relationship between FC and CRP cluster assignment was visualised using alluvial plots and side-by-side comparisons of mean cluster trajectories for the optimal models. Stratified results for CD and UC subjects are also reported.

#### Software

All analysis was performed in the R statistical software^20^; software/package versions are shown in Supplementary Note 3. Analytical reports for the LIBDR have been generated using the Quarto scientific publishing system and are hosted online (https://vallejosgroup.github.io/IBD-Inflammatory-Patterns/) with code used to perform the LIBDR and Danish analyses publicly available under an open source license (https://github.com/VallejosGroup/IBD-Inflammatory-Patterns).

#### Ethics

Usage of the Scottish dataset was approved by the local Caldicott Guardian (Project ID: CRD18002, registered NHS Lothian information asset #IAR-954). Studies based on Danish registry data alone are not required to obtain permission from the regional ethics committees as confirmed by The Central Denmark Region Committees on Health Research Ethics (legislation: 1–10–72-148-19).

#### Patient and public involvement

Patients or the public were not involved in the design, conduct, reporting or dissemination plans of our research.

#### Role of the funding source

Funders were not involved in the study design, collection, analysis, or interpretation of the data, writing, or decision to submit the paper for publication.

## Results

### Cohort derivation and description

In the Scottish cohort, 10545 FC and 49364 (9898 after preprocessing) CRP observations were available. In the Danish cohort, 67986 FC and 207141 (40648 after preprocessing) CRP observations were available. Figure 1 outlines the derivation of the cohorts; Table 1 describes key demographic factors. The distribution of log-transformed FC and CRP values is shown in Figure S5.

**Table 1.** Demographic and clinical data at diagnosis for subjects meeting the faecal calprotectin (FC) or C-reactive protein (CRP) study inclusion criteria. Continuous data are presented as median and interquartile range. Categorical data are presented as counts and percentages. Missingness is only directly reported if values were missing. The column labelled as “Overlap” denotes subjects which met the inclusion criteria for both FC and CRP modelling. Missing observations for upper gastrointestinal inflammation were assumed to be “not present” (Supplementary Note 1). Missingness was not inferred to be a value for any other variable. There are no IBDU diagnoses reported for the Danish cohort as these were mapped to either CD or UC (Supplementary Note 1).

|  | The Lothian IBD Registry |  |  | Danish national registry |  |  |
| --- | --- | --- | --- | --- | --- | --- |
|  | FC analysis<br>(n = 1036) | CRP analysis<br>(n = 1838) | Overlap<br>(n = 808) | FC analysis<br>(n = 7880) | CRP analysis<br>(n = 10041) | Overlap<br>(n = 5685) |
| Age at diagnosis | 33 (25–75) | 37 (24–55) | 32 (20–50) | 33 (22–53) | 38 (24–57) | 32 (22–52) |
| Male sex | 503 (48.6%) | 956 (52.0%) | 397 (49.1%) | 3648 (46.3%) | 4693 (46.7%) | 2594 (45.6%) |
| IBD type |  |  |  |  |  |  |
| Crohn's disease (CD) | 544 (52.5%) | 805 (43.8%) | 451 (55.8%) | 3931 (49.9%) | 4415 (44.0%) | 2954 (52.0%) |
| Ulcerative colitis (UC) | 380 (36.7%) | 847 (46.1%) | 276 (34.2%) | 3949 (50.1%) | 5626 (56.0%) | 2731 (48.0%) |
| IBDU | 112 (10.8%) | 186 (10.1%) | 81 (10.0%) | .. | .. | .. |
| Baseline FC (µg/g) | 740 (320–1070) | .. | 760 (330–1081) | 574 (148–1613) | .. | 652 (171–1737) |
| FC observations | 9 (6–13) | .. | 9 (6–14) | 7 (4–11) | .. | 8 (5–12) |
| Baseline CRP (µg/mL) | .. | 8 (3–24) | 8 (3–20) | .. | 12 (3–45) | 20 (3–38) |
| Total unprocessed CRP observations | .. | 20 (10–36) | 26 (14–36) | .. | 19 (9–32) | 22 (12–34) |
| Total CRP observations after pre-processing | .. | 6 (4–7) | 6 (5–7) | .. | 4 (3–5) | 4 (3–5) |
| **Advanced therapies (AT)** |  |  |  |  |  |  |
| AT within follow-up for CD | 270 (49.6%) | 374 (46.5%) |  | 2108 (53.6%) | 2246 (50.9%) |  |
| AT within one year for CD | 129 (23.7%) | 167 (20.7%) |  | 1517 (38.6%) | 1549 (35.1%) |  |
| AT within follow-up for UC | 108 (28.4%) | 85 (10.0%) |  | 1293 (32.7%) | 1702 (30.3%) |  |
| AT within one year for UC | 42 (11.1%) | 35 (4.1%) |  | 789 (20.0%) | 978 (17.4%) |  |
| **Crohn's disease** |  |  |  |  |  |  |
| Montreal location |  |  |  |  |  |  |
| L1 | 168 (30.9%) | 249 (30.9%) | 136 (30.2%) |  |  |  |
| L2 | 200 (36.8%) | 287 (35.7%) | 157 (34.8%) |  |  |  |
| L3 | 169 (31.1%) | 246 (30.6%) | 153 (33.9%) |  |  |  |
| Missing | 7 (1.3%) | 23 (2.9%) | 5 (1.1%) |  |  |  |
| **Upper GI inflammation** |  |  |  |  |  |  |
| Present | 77 (14.2%) | 87 (10.8%) | 68 (15.1%) |  |  |  |
| Not Present | 467 (85.8.4%) | 718 (89.2%) | 383 (84.9%) |  |  |  |
| **Montreal behaviour** |  |  |  |  |  |  |
| B1 | 443 (81.4%) | 607 (75.4%) | 364 (80.7%) |  |  |  |
| B2 | 63 (11.5%) | 110 (13.7%) | 54 (12.0%) |  |  |  |
| B3 | 28 (5.1%) | 61 (7.6%) | 25 (5.5%) |  |  |  |
| Missing | 10 (1.8%) | 27 (3.4%) | 8 (1.8%) |  |  |  |
| **Perianal disease** |  |  |  |  |  |  |
| Present | 79 (14.5%) | 113 (14.0%) | 68 (15.1%) |  |  |  |
| Not Present | 459 (84.4%) | 673 (83.6%) | 379 (84.0%) |  |  |  |
| Missing | 6 (1.1%) | 19 (2.4%) | 4 (0.9%) |  |  |  |
| Smoking status |  |  |  |  |  |  |
| Current or previously | 166 (30.5%) | 274 (34.0%) | 138 (30.6%) |  |  |  |
| Never | 352 (64.7%) | 484 (60.1%) | 293 (65.0%) |  |  |  |
| Missing | 26 (4.8%) | 47 (5.8%) | 20 (4.4%) |  |  |  |
| Ulcerative colitis |  |  |  |  |  |  |
| Montreal extent |  |  |  |  |  |  |
| E1 | 50 (13.2%) | 155 (18.3%) | 27 (9.8%) |  |  |  |
| E2 | 155 (40.8%) | 330 (39.0%) | 111 (40.2%) |  |  |  |
| E3 | 170 (44.7%) | 341 (40.3%) | 133 (48.1%) |  |  |  |
| Missing | 5 (1.3%) | 21 (2.5%) | 5 (1.8%) |  |  |  |
| Smoking status |  |  |  |  |  |  |
| Current or previously | 121 (31.8%) | 336 (39.7%) | 89 (32.2%) |  |  |  |
| Never | 237 (62.4%) | 444 (52.4%) | 167 (60.5%) |  |  |  |
| Missing | 22 (5.8%) | 67 (7.9%) | 20 (7.2%) |  |  |  |

### Modelling of FC trajectories

For the LIBDR cohort, AIC suggested the 10-cluster model (Figure S6), whilst BIC suggested the 9-cluster model (Figure S7) was most appropriate (Table S1, Figure S8 (A)). However, the 8-cluster model was chosen as a parsimonious choice, as it captures the main observed longitudinal patterns without generating very small clusters (<50 individuals) which could be difficult to interpret.

Figure 2 (A) shows representative cluster profiles for the 8-cluster model in LIBDR, ordered from lowest (FC1) to highest (FC8) cumulative inflammatory burden. FC2 (n=67; ∼6%) represents low FC values throughout the whole observation period. FC1 (n=140; ∼14%), FC3 (n=157; ∼15%), and FC7 (n=244; ∼24%) were characterised by initially high FC values (>250 μg/g) which decreased over time at different rates. Whilst FC1 exhibited a sharp decrease within the first year, the decrease was more gradual for FC3 and FC7, where FC was normalised (<250 μg/g) approximately around two and five years post diagnosis, respectively. Furthermore, FC4 (n=103; ∼10%), FC5 (n=67; ∼6%) and FC6 (n=64; ∼6%) captured relapsing and remitting patterns of gastrointestinal inflammation. Finally, FC8 (n=194; ∼19%) represented individuals with consistently high FC values.

**Figure 2.**
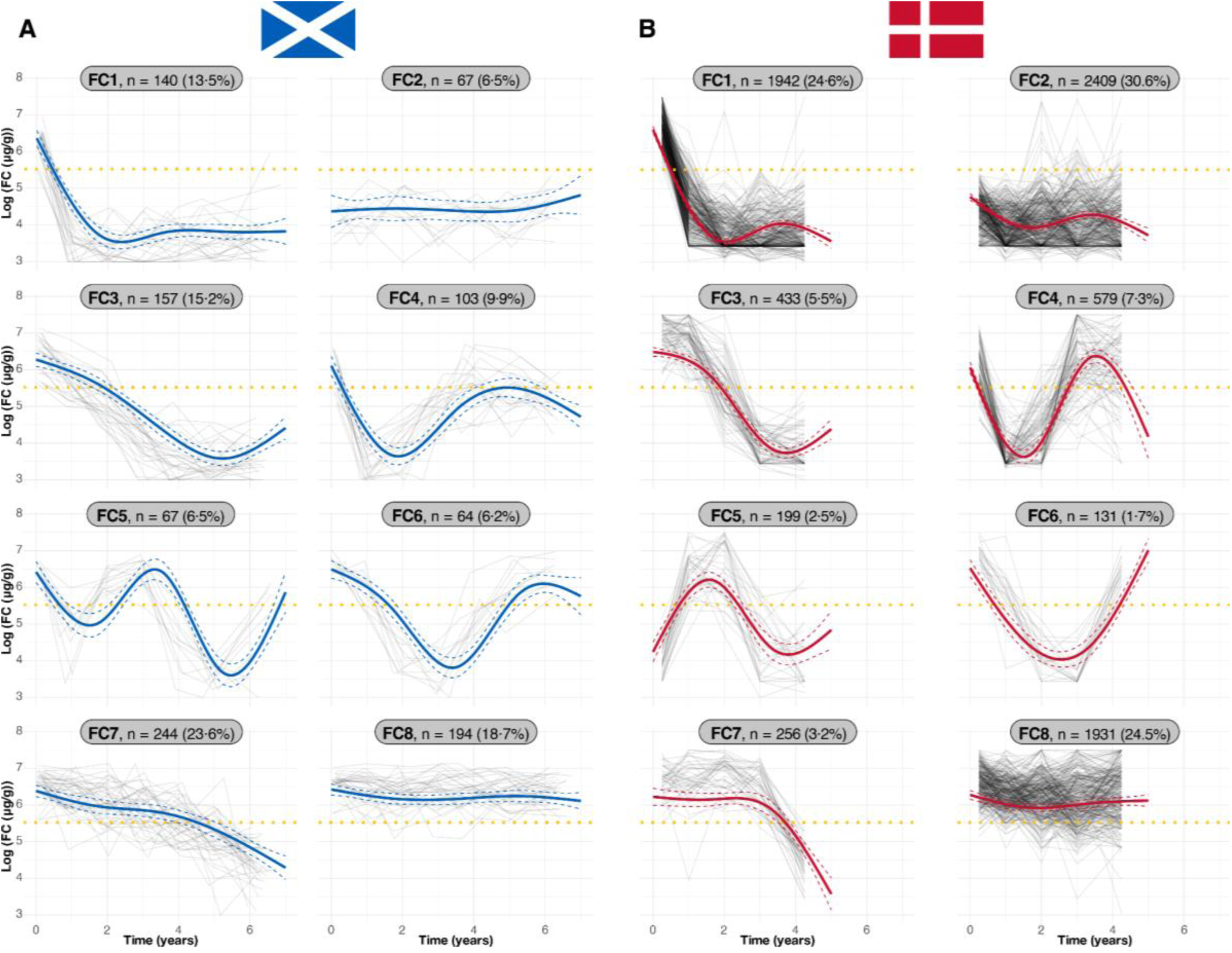
Cluster trajectories obtained from LCMM assuming eight clusters fitted to FC data (log-transformed) in (A) the Lothian IBD Registry and (B) Danish national registry data. The blue and red lines indicate predicted mean cluster profiles with 95% confidence intervals for the Lothian IBD Registry and Danish data respectively. The yellow dotted lines indicate log(250μg/g). For visualisation purposes, pseudo subject-specific trajectories have been generated by amalgamating observations from randomly selected groups of six subjects. Clusters are ordered from lowest (FC1) to highest (FC8) cumulative inflammatory burden in the Lothian data, calculated from the area under the cluster trajectories. Danish cluster labels chosen based on visual similarity to the former clusters. Cluster sizes are shown as panel titles.

An 8-cluster model was then applied to the Danish data. Despite being an independent analysis, using data from another healthcare system, and with a shorter follow-up period, the shape of the cluster-specific trajectories largely matched those found for the LIBDR cohort (Figure 2 (B)). The most marked difference is within the relapsing-remitting clusters (FC4-6), which may be partly explained by the smaller size of these clusters in the LIBDR cohort. The proportion of individuals allocated to each cluster differed between cohorts. For example, whilst FC2 represented ∼6.5% in the Scottish cohort, nearly one third of patients (∼30.1%) were assigned to FC2 in the Danish cohort.

### Associations with respect to FC cluster assignments

Associations with respect to age, sex and IBD type were considered in both cohorts with the relapsing-remitting clusters (FC4–6) grouped for ease of comparison. Effect sizes were largely consistent between cohorts (Figures S9–S13). The effect of age differed across clusters (Figure S9), but this may be partially due to differences in treatment between younger and older individuals. Males were more likely to be assigned to the clusters with the highest inflammatory burden, FC7 and FC8 (Figures S9–S11). IBD type was associated with cluster assignments (Figure S13). For example, FC1 (rapid remitters) was enriched by patients with CD in both cohorts (62.9% in LIBDR and 63.0% in the Danish data vs 50.9% and 45.6% respectively elsewhere). However, whilst the relapsing-remitting clusters (FC4–6) mostly consists of individuals with UC in the Danish cohort (87.7%), this was not the case for LIBDR.

For the LIBDR, a similar analysis was performed after stratifying by IBD type (UC and CD only) and considering additional phenotyping (Figures S14–S20). In most cases, effect sizes were not statistically significant (in some cases this was due to low counts and small cluster sizes). However, amongst CD patients, those without upper GI inflammation were more likely to be assigned to the clusters with the lowest inflammatory burden, FC1 or FC2 (2.4% and 3.03% L4 vs 18.1% elsewhere; Figure S16). Finally, UC patients with ulcerative proctitis were less likely to be assigned to FC3 (2.44% E1 versus 14.2% elsewhere; Figure S20).

### FC cluster assignment and IBD therapy usage

In Scottish and Danish patients with CD, 23.7% and 38.6% received an advanced therapy (AT) within one year of diagnosis, respectively (Table 1). This increased to 49.6% and 53.6% by the end of the observation period (seven and five years, respectively). For patients with UC, 11.1% and 20.0% received an AT within one year of diagnosis, respectively for Scottish and Danish patients. This increased to 28.4% and 32.7% by the end of follow-up. Overall AT rates and the distribution of time to first AT were not homogeneous across clusters (Figure 3). For example, whilst AT prescription rates in FC1 largely matched the overall FC cohort, prescriptions were generally earlier in this cluster, especially in patients with CD. In both FC7 (the substantially delayed remitting cluster) and in FC8 (consistently high inflammation), the cumulative AT rates for CD were high. By the end of follow-up in Scotland and Denmark, 55.1% and 80.9% were on AT in FC7 and 56.8% and 56.7% in FC8. However, early prescriptions of AT were noticeably lower than in other clusters including FC1.

**Figure 3.**
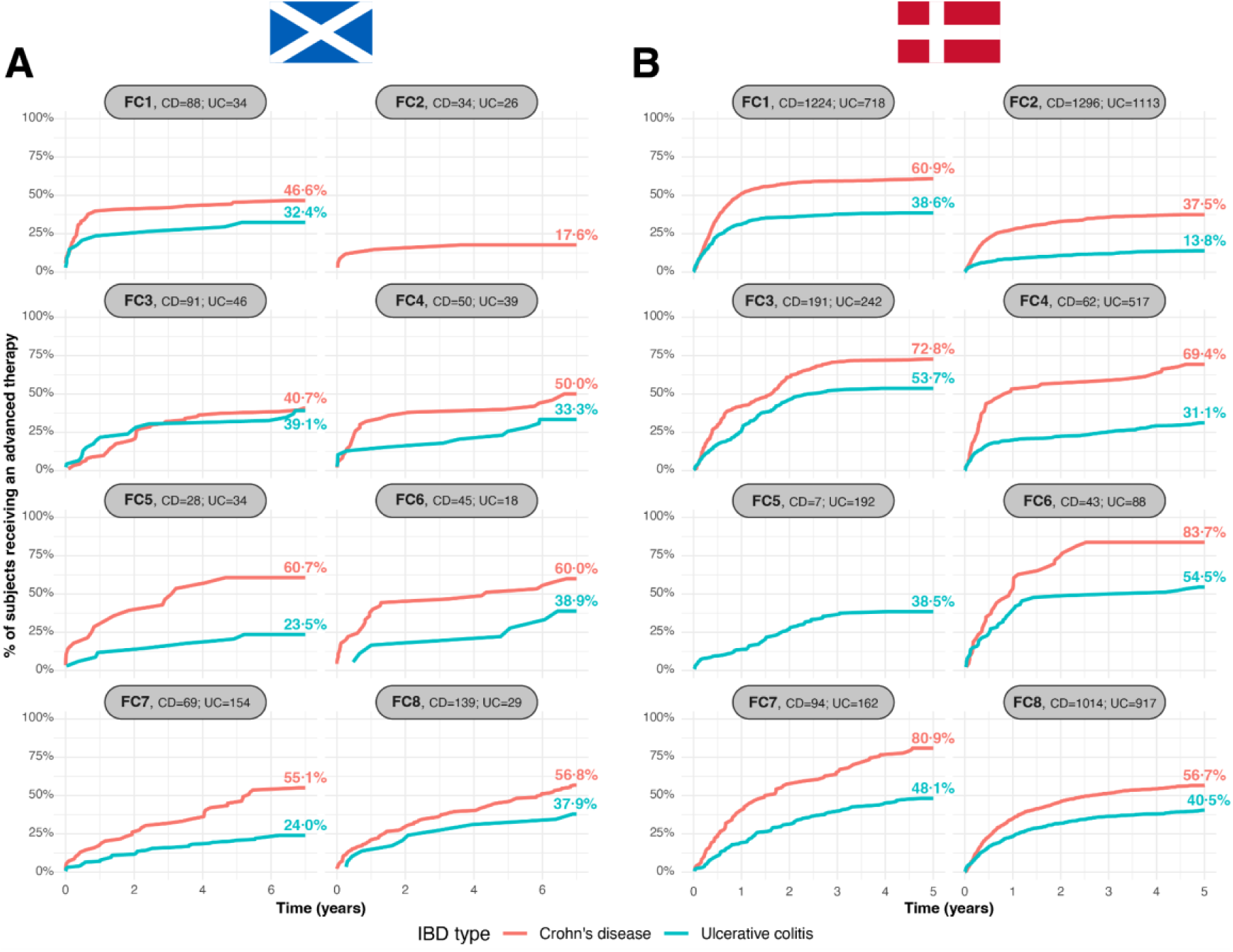
FC cluster-specific cumulative distribution for first-line advanced therapy prescribing for Crohn’s disease (red) and ulcerative colitis (teal) subjects in (A) the Lothian IBD Registry and (B) national Danish registry data. Clusters are ordered from lowest (FC1) to highest (FC8) cumulative inflammatory burden. The number of CD and UC subjects present in each cluster is displayed as panel titles. Total advance therapy prescribing (as a percentage of the corresponding group) within seven/five years from diagnosis is shown next to each distribution curve. Curves which would describe fewer than ten subjects are not shown.

The detailed therapy data available from the Danish registries allows for a more granular exploration of multiple lines of advanced therapy, immunosuppressant use and courses of corticosteroids separately for CD (Figure S21) and UC (Figure S22).

In CD, FC1 is characterised by early use of steroids, AT and immunosuppressants, with therapy remaining remarkably stable throughout (no additional lines of AT and no further courses of steroids). FC2 has the highest number of patients not on AT after 1 (72.3%) and 5 years (62.5%). In FC3 (delayed remitters), FC4 and FC6 (the two main relapsing remitting clusters for patients with CD), second, third and fourth line AT prescriptions are noted along with multiple courses of corticosteroids. These observations would be consistent with the challenges of treating patients with IBD, where for some patients it takes time to find the right drug to induce deep remission, requiring multiple steroid courses in the interim. A similar pattern is seen in FC7, where 25.5% had 3 or more different advanced therapies, 23.7% had 5 or more steroid courses and 87.2% received an immunomodulator within five years. In FC8, 29.3% and 13.1% of patients had tried ≥2 and ≥3 different ATs respectively, suggesting the persistently elevated inflammation levels during follow-up are at least in part due to non-response to multiple lines of therapy. In patients with UC, FC2 has the highest percentage of patients without treatment, FC1 is relatively stable over time, and FC8 has the highest percentage of multiple types of treatment.

Surgical resections for patients with CD are shown in Figure S23. It is noteworthy that some of the patients assigned to a delayed remitting group (FC3 and FC7) received surgical resections later in follow-up. Overall colectomy rates in UC were very low (Figure S24), although higher rates were observed in clusters with a higher cumulative inflammatory burden (10.5% in FC8 compared to 2.8% and 1.4% in FC1 and FC2).

### Modelling of CRP trajectories

AIC and BIC both suggested the 8-cluster model was the most appropriate (Table S2) for the LIBDR cohort. Visual inspection supported this finding as the 9-cluster model did not identify new trajectories when compared to the 8-cluster model, producing two trajectories with consistently low CRP (Figure S25). In contrast, the 7-cluster model (Figure S26) lacks one of the clinically interesting trajectories, characterised by an initially elevated CRP which then decreases to slightly above biochemical remission after one year, when compared to the 8-cluster model.

Figure 4 (A) presents exemplar cluster profiles for the 8-cluster model. Over a third of subjects (n=702; ∼38%) were assigned to CRP1, defined by consistently low CRP (< 5μg/mL). CRP2 (n=225; ∼12%) was characterised by high CRP at diagnosis which rapidly decreased shortly thereafter remaining low. CRP3 (n=51; ∼3%) and CRP4 (n=60; ∼3%) are both small clusters with the former described by low CRP until the last year of follow-up and the latter presenting low inflammation within the first year before increasing until the third year where the inflammation then decreases again. CRP5 (n=110; 6%) is characterised by elevated CRP at diagnosis which then decreases gradually over time. CRP6 (n=434; 24%) consists of trajectories which are elevated at diagnosis which then falls slightly for the first two years after diagnosis, remaining elevated across the remaining duration of follow-up. CRP8 (n=172; 9%) is consistently elevated and does not change over time.

**Figure 4.**
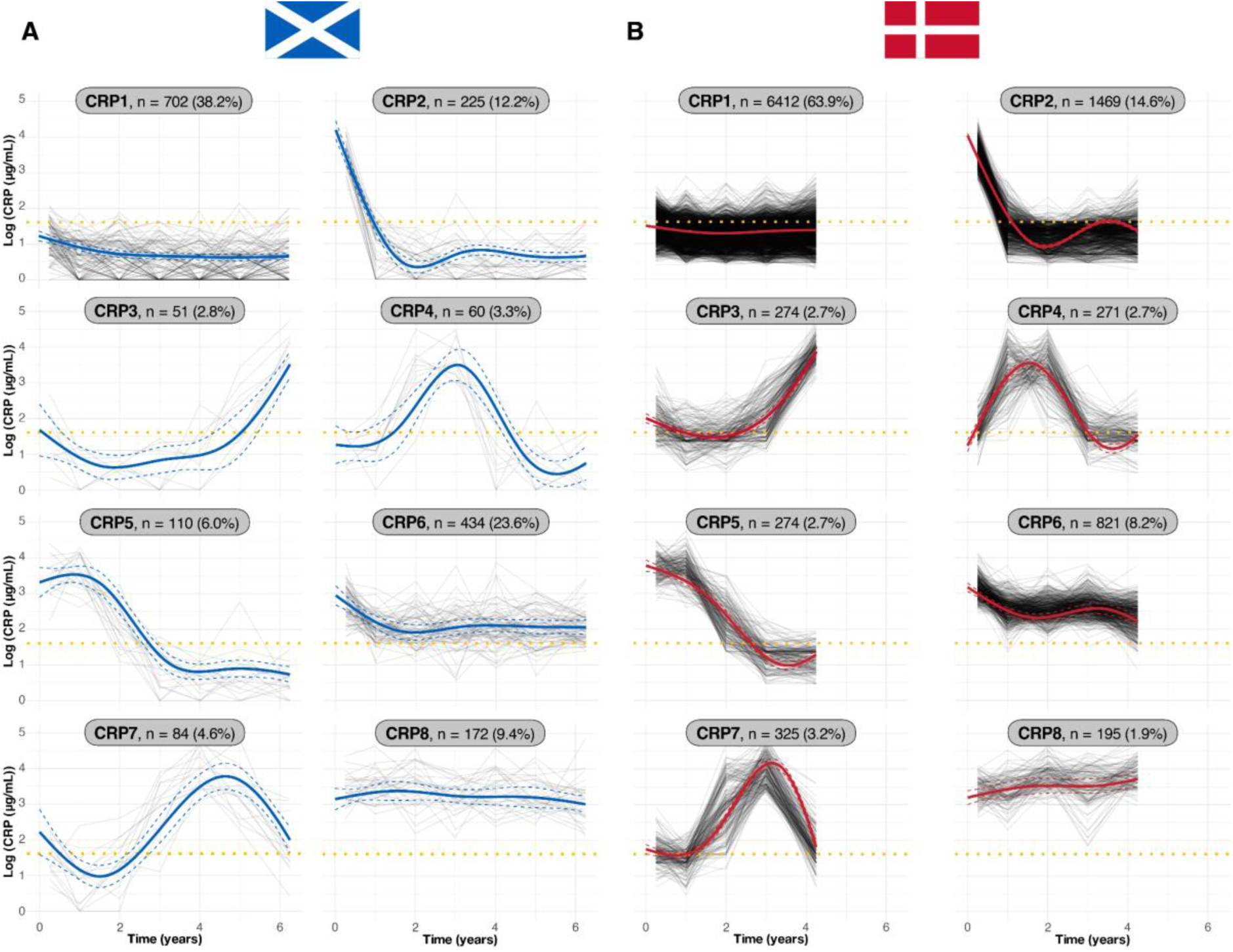
Cluster trajectories obtained from LCMM assuming eight clusters fitted to processed CRP data (log-transformed) for subjects in (A) the Lothian IBD Registry and (B) national Danish registry data. Blue and red lines indicate predicted mean cluster profiles with 95% confidence intervals. The yellow dotted lines indicate log(5μg/mL). For visualisation purposes, pseudo subject-specific trajectories have been generated by amalgamating observations from randomly selected groups of six subjects. Clusters are ordered from lowest (CRP1) to highest (CRP8) cumulative inflammatory burden, calculated from the area under the cluster trajectories. Danish cluster labels chosen based on visual similarity to the former clusters. Cluster sizes are shown as panel titles.

As in the FC analysis, the shape of CRP clusters inferred for the LIBDR cohort was largely recapitulated when the 8-cluster model was applied to the Danish cohort (Figure 4 (B)).

### Associations with CRP cluster assignments

Figures S27–S29 visualise the distribution of age, sex and IBD type within each CRP cluster. Results were largely consistent across the cohorts. Older patients were more likely to be assigned to a CRP cluster with higher cumulative inflammatory burden (Figure S27). IBD type was not evenly distributed among clusters (Figure S29), with CRP1, CRP3, CRP4 and CRP7 enriched for UC patients (59.3%, 56.8%, 61.7% and 72.6% vs 32.3% elsewhere in LIBDR, with similar proportions for the Danish data.The association plots for CD and UC disease sub-phenotypes and CRP cluster assignment are shown in Figures S30–S35. Advanced therapy (AT) prescribing patterns in CRP clusters are shown in Figure S36.

### Uncertainty in cluster assignments

Whilst cluster assignments are static, uncertainty in cluster assignments can depend on the amount of information available for each individual. In the LIBDR FC analysis, with the exception of FC2, cluster assignments were on average more uncertain for subjects with a short follow-up (Figure 5 (A)). This is particularly the case for clusters sharing similar early trends. For example, individuals assigned to FC1 (rapid remitters) had a low average probability of being assigned to FC8 (non-remitters) and vice-versa, even for those with a short follow-up. This is not the case for FC3 and FC6, both of which capture similar FC trajectories within the first two years. On average, cluster assignments were less uncertain in the CRP analysis, even for individuals with a short follow-up (Figure 5 (B)). This is expected as CRP clusters are associated with more distinct early trajectories.

**Figure 5.**
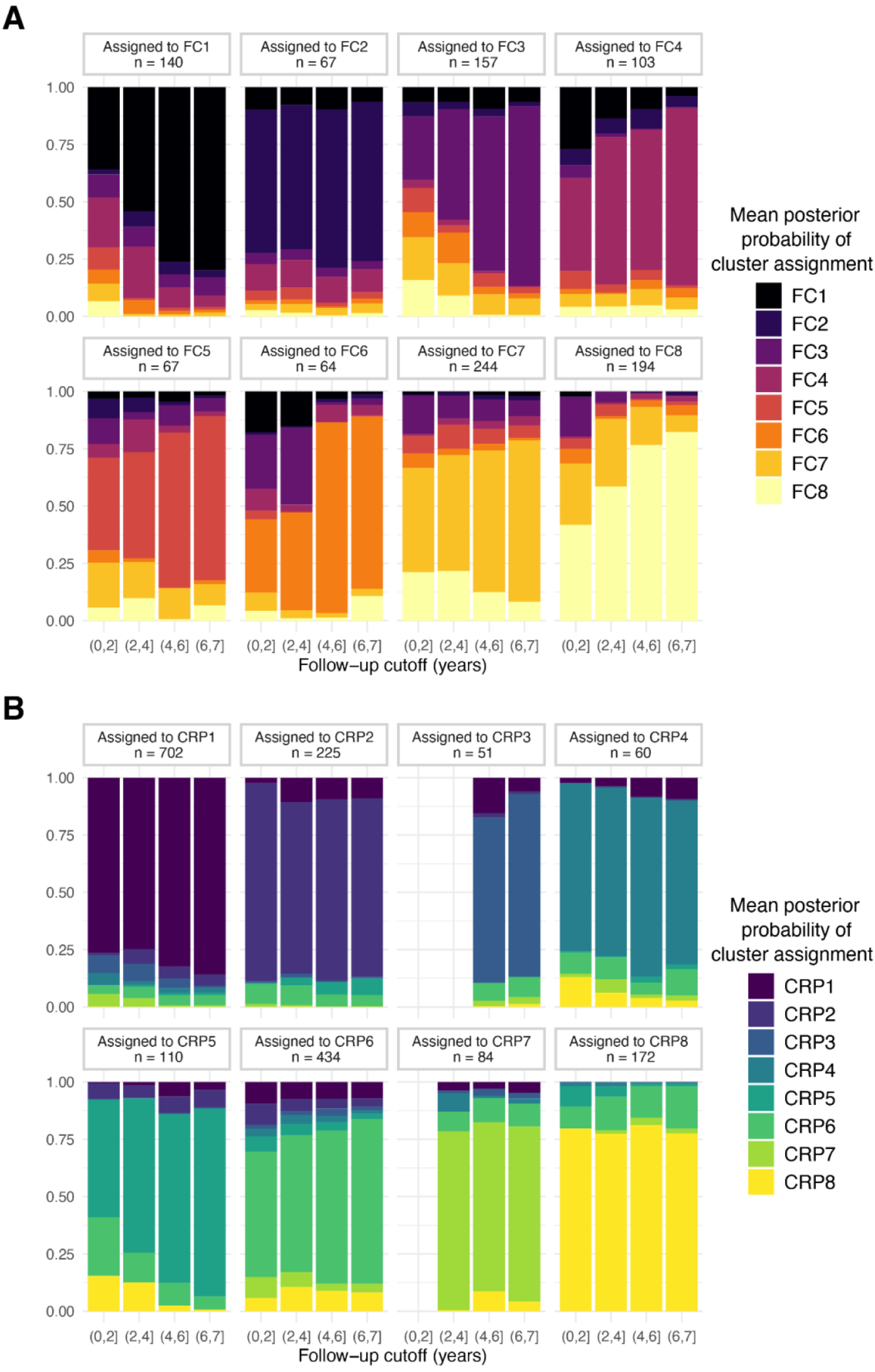
Exploration of cluster assignment uncertainty for A) faecal calprotectin (FC) and B) CRP clusters in the Lothian IBD Registry cohort. LCMM assigns individuals to the cluster with the highest estimated probability. For individuals assigned to a given cluster, bars show the average probability of cluster assignment to each possible cluster. Results are stratified according to follow-up length, defined as the time difference between diagnosis and the last available biomarker measurement (FC or CRP for A) and B), respectively). Clusters are ordered from lowest (FC1 and CRP1) to highest (FC8 and CRP8) cumulative inflammatory burden, with adjacent clusters coloured sequentially in the plots for FC (black to yellow) and CRP (blue to yellow).

### Comparison of FC and CRP clustering

In the LIBDR cohort, all FC clusters were well represented amongst the 808 subjects included in both analyses (the *overlap sub-cohort*), but the proportion in CRP8 was low (∼27% of CRP8 was in the overlap sub-cohort vs ∼46% elsewhere; Figure S37). As shown in Figure 6 (A), there was low agreement between FC and CRP clustering (the same occurred in the Danish data, Figure S38). Whilst most subjects in FC1 (71.6%) were also in CRP1 or CRP2, this relationship was not mirrored as 81.2% of subjects in the latter two CRP clusters were assigned to substantially different FC clusters. However, CRP8 did overlap with elevated FC as the majority of subjects in this cluster (80.8%) were assigned to either FC7 or FC8.

**Figure 6.**
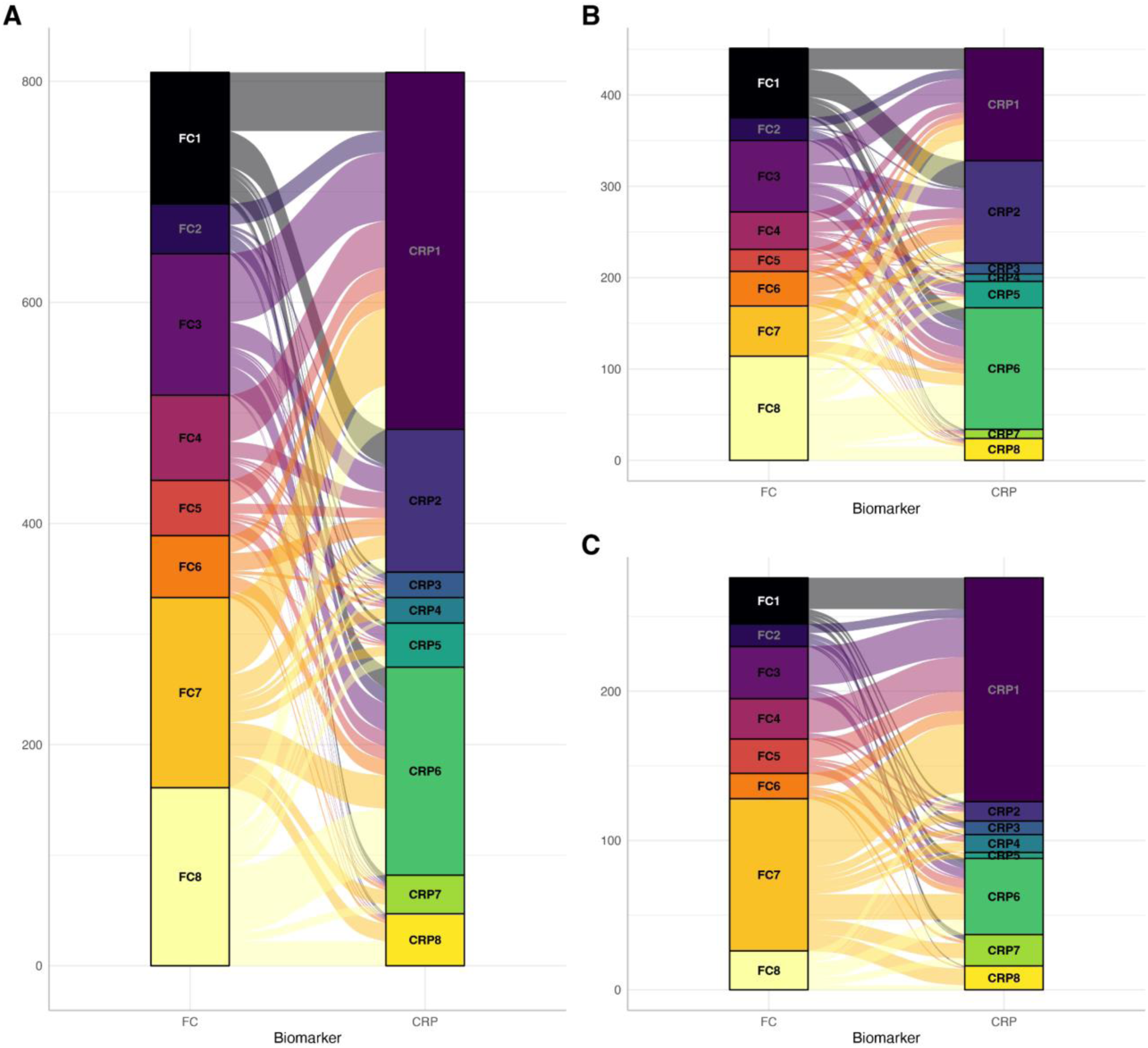
Comparison between faecal calprotectin (FC) and processed CRP for models with chosen specification (three NCS) assuming eight clusters for subjects in the Lothian IBD Registry cohort. Results are reported based on the overlap cohort, consisting of 808 subjects included in both the FC and CRP analysis. (A) all subjects; (B) Crohn’s disease; and (C) ulcerative colitis. Each segment denotes the size of the cluster whilst the alluvial segments connecting the nodes visualises the number of subjects shared between clusters.

## Discussion

We have characterised IBD behaviour using long-term longitudinal trends of objective inflammatory markers routinely collected for clinical care. Our analysis has uncovered eight clusters with distinct inflammatory profiles based on FC and CRP, respectively. This was developed in the Lothian IBD Registry and then validated in Danish national registry data. Our data highlights the heterogeneity of the disease course, and presents a novel approach for understanding real-world inflammatory activity patterns in IBD patients. For the first time, we are capturing the dynamic nature of the disease in a more biologically nuanced way than traditional behaviour endpoints such as treatment escalation, surgery and Montreal progression. This represents a marked shift from the traditional symptom-based behaviour profiles exemplified by the IBSEN cohorts over a decade ago.^10,11^

This study builds on our earlier proof-of-concept work, where we first demonstrated the feasibility of using long-term individualised profiles of FC to cluster a subset of patients with CD.^21^ In our previous analysis of Lothian patients, we identified four distinct FC clusters: one cluster with persistently high FC (non-remitters) and three clusters with different downward longitudinal trends. Subjects who underwent surgery, hospitalisation, or Montreal progression within a year of diagnosis were excluded in addition to subjects with an FC below 250 at diagnosis. Here, we expand upon this by considering a substantially larger Lothian IBD cohort (FC cohort, n=1036; CRP cohort, n=1838), with longer follow-up, not excluding patients based on indicators of disease severity, and including patients with UC or IBDU. This has generated results with greater representativeness across IBD phenotypes, uncovering more granular structure with eight distinct FC clusters rather than four. It is noteworthy FC2 could not have been found in the previous study due to exclusion criteria. In addition, we introduced modelling CRP.

Whilst the modelling suggested further partitioning of the FC data was possible (Figure S5, Figure S7 (A)), we selected the eight-cluster model as a parsimonious choice that captured the key inflammatory patterns (Figure 2). Notably, the CRP data also partitioned into eight distinct clusters. Together, the clusters broadly fall into four patterns of inflammatory behaviour mirroring those recognised by gastroenterologists managing IBD patients (i) rapid remitters (FC1 and CRP2), (ii) delayed remitters (FC3, FC7 and CRP5), (iii) relapsing-remitters (FC4–6 and CRP4 and CRP7), and (iv) non-remitters (FC8 and CRP6 and CRP8). These classifications provide new insights into the inflammatory time-course of IBD patients.

Critically, replication in an independent IBD cohort demonstrates the robustness of our findings, indicating the longitudinal patterns described above characterise inherent properties of the disease course. Both cohorts represent real-world IBD populations with complementary properties. The Danish cohort is substantially larger, has nation-wide coverage, and has more detailed treatment information (including primary care prescribing). However, phenotyping is more limited with Montreal classification unavailable. Instead, LIBDR represents a single Scottish health board and is more moderate in size, but has longer follow-up. Moreover, extensive data curation provides more accurate diagnostic data and additional phenotyping, helping to understand how the clusters relate to well-established IBD classifications.

We observed broadly poor agreement between FC and CRP clusters. The lack of overlap between the two is perhaps unsurprising. CRP is a marker of systemic inflammation, whilst FC is more specific for detecting inflammation at a mucosal level, making both biomarkers complimentary for monitoring patients with IBD^22^. Studies have also shown a proportion of UC patients will have lower CRP values at diagnosis,^23^ as is seen in CRP1 which is significantly enriched for UC cases.

We observed several observations regarding cluster assignment. Cluster assignment was unevenly distributed across IBD types (Figures S13 and S29). Although CD patients with L4 disease were less represented in FC1 and FC2, there was no association with ileal versus colonic disease location. Males were slightly overrepresented in the FC8 cluster, which is characterised by persistently elevated values. In the recent SEXEII study, male patients with UC were reported as being more likely to have extensive colonic involvement and require abdominal surgery, which may account for this finding.^24^ CRP cluster membership was also associated with smoking and older age, with both more likely to have higher inflammatory burdens.

Multiple lines of evidence, including the recent PROFILE study,^25^ have clearly demonstrated improved disease control and outcomes in Crohn’s patients receiving early AT. As such, we anticipated the biggest driver of inflammatory patterns over time would be AT and this effect would be more pronounced in CD versus UC given the era of our cohort. Indeed, rates of AT use were not homogenous across clusters or IBD subtypes. Overall, in the FC LIBDR cohort, AT were used in 49.6% of CD patients of whom 47.7% started AT in the first year. In FC1, similar rates of AT were used for CD patients, however they were used earlier. Rates of AT prescriptions for patients with CD in FC8 (persistently high levels of inflammatory behaviour) were similar, but started later in the disease course. This cluster may represent a more refractory group of patients, with a high-risk phenotype. Whilst this may provide additional support for the use of early AT, a causal interpretation of these effects is not possible in this observational study. Additional work with alternative cohorts where treatment assignment is randomised is planned.

In this study we have modelled FC and CRP separately. We anticipate a multivariate approach which simultaneously considers other biomarkers of disease activity, such as haemoglobin, albumin, and platelet count, could further increase the robustness of cluster assignments. Such analysis may also consider pre-diagnostic biomarker measurements, following recent observations in Danish registry data,^26^ as well as metabolomics, genetics or microbiome data to inform cluster assignments, but this is beyond the scope of the current study.

Critically, this study is limited by the use of observational data. As samples were requested after every IBD clinic appointment, the frequency of biomarker measurements is likely to be related to disease severity, and very mild cases may be excluded from our cohort. Moreover, the FC and CRP clusters cannot be directly used in a prognostic way. Further work is needed to assess associations between clusters and IBD related complications (such as steroid use, hospitalisations and surgery) but also non-conventional complications associated with high cumulative inflammatory burden, such as major cardiovascular adverse events^27^, neuropsychiatric illness^28^, and malignancy^29^.

Classifying patients by their inflammatory behaviour is a paradigm shift in thinking about disease behaviour compared with the previous symptom based profiles first reported by the IBSEN cohorts. Moreover, this approach - rooted in data widely available in clinical settings^30^ and probabilistic modelling - paves the way for predictive analytics integrated into a clinical support tool for population wide risk stratification and individual patient level prognostication and treatment

## Data sharing statement

As the data collected for this study has been derived from unconsented patient data, it is not possible to share subject-level data with external entities. Detailed summary level data is available online at https://vallejosgroup.github.io/IBD-Inflammatory-Patterns. Analysis code is also publicly available (https://github.com/VallejosGroup/IBD-Inflammatory-Patterns).

Analyses relating to the DK cohort used data from Danish National Health registries (https://sundhedsdatastyrelsen.dk). These data are protected by the Danish Act on Processing of Personal Data and access requires an application to the Danish Data Protection Agency and the Danish Health Data Authority.

## Author contributions

CAV, CWL, and NCC were involved in conceptualising the study. CRB, SO, ATE, GRJ, and NP curated the LIBDR data. NCC and CAV implemented computer code, tested existing code components, and conducted the formal analysis and visualisation for the LIBDR. TJ obtained funding for and built the Danish PREDICT data infrastructure on which the Danish analyses were performed. MVV curated the Danish data and conducted the validation analyses and visualization on the Danish data. The original draft was written by NCC, NP, ATE, CMR, MVV, CWL and CAV with all authors involved in reviewing and editing the manuscript. KMG, CWL, CAV, AS and TJ provided supervisory support.

## Competing interests

NP has served as a speaker for Janssen, Takeda and Pfizer. BG has acted as consultant to Galapagos and Abbvie and as speaker for Abbvie, Jansen, Takeda, Pfizer and Galapagos. GRJ has served as a speaker for Takeda, Janssen, Abbvie, Fresnius and Ferring. TJ has served as a speaker and/or consultant for Ferring and Pfizer. CWL has acted as a speaker and/or consultant to AbbVie, Janssen, Takeda, Pfizer, Galapagos, GSK, Gilead, Vifor Pharma, Ferring, Dr Falk, BMS, Boehringer Ingelheim, Eli Lilly, Merck, Novartis, Sandoz, Celltrion, Cellgene, Amgen, Samsung Bioepis, Fresenius Kabi, Tillotts, Kuma Health, Trellus Health and Iterative Health. None of the other authors report any conflicts of interest.

## Funding

CWL is funded by a UKRI (UK Research and Innovation) Future Leaders Fellowship ‘Predicting outcomes in IBD’ (MR/S034919/1). G-RJ is funded by a Wellcome Trust Clinical Research Career Development Fellowship. NC-C was partially supported by the Medical Research Council and The University of Edinburgh via a Precision Medicine PhD studentship (MR/N013166/1). MVV, AS, and TJ are funded by a Center of Excellence Grant (DNRF148) to TJ from the Danish National Research Foundation.

## Data Availability

As the data collected for this study has been derived from unconsented patient data, it is not possible to share subject-level data with external entities. Detailed summary level data is available online at https://vallejosgroup.github.io/Lothian-IBDR. The code used to conduct the analysis is also publicly available (https://github.com/VallejosGroup/Lothian-IBDR).

## Acknowledgements

This work uses data provided by patients and collected by the NHS as part of their care and support. The Lothian component of this work has made use of the resources provided by the Edinburgh Compute and Data Facility (ECDF). The analysis of the Danish cohort was performed using the National Genome Center platform.

## Supplemental display items

### Supplemental figures

**Figure S1.**
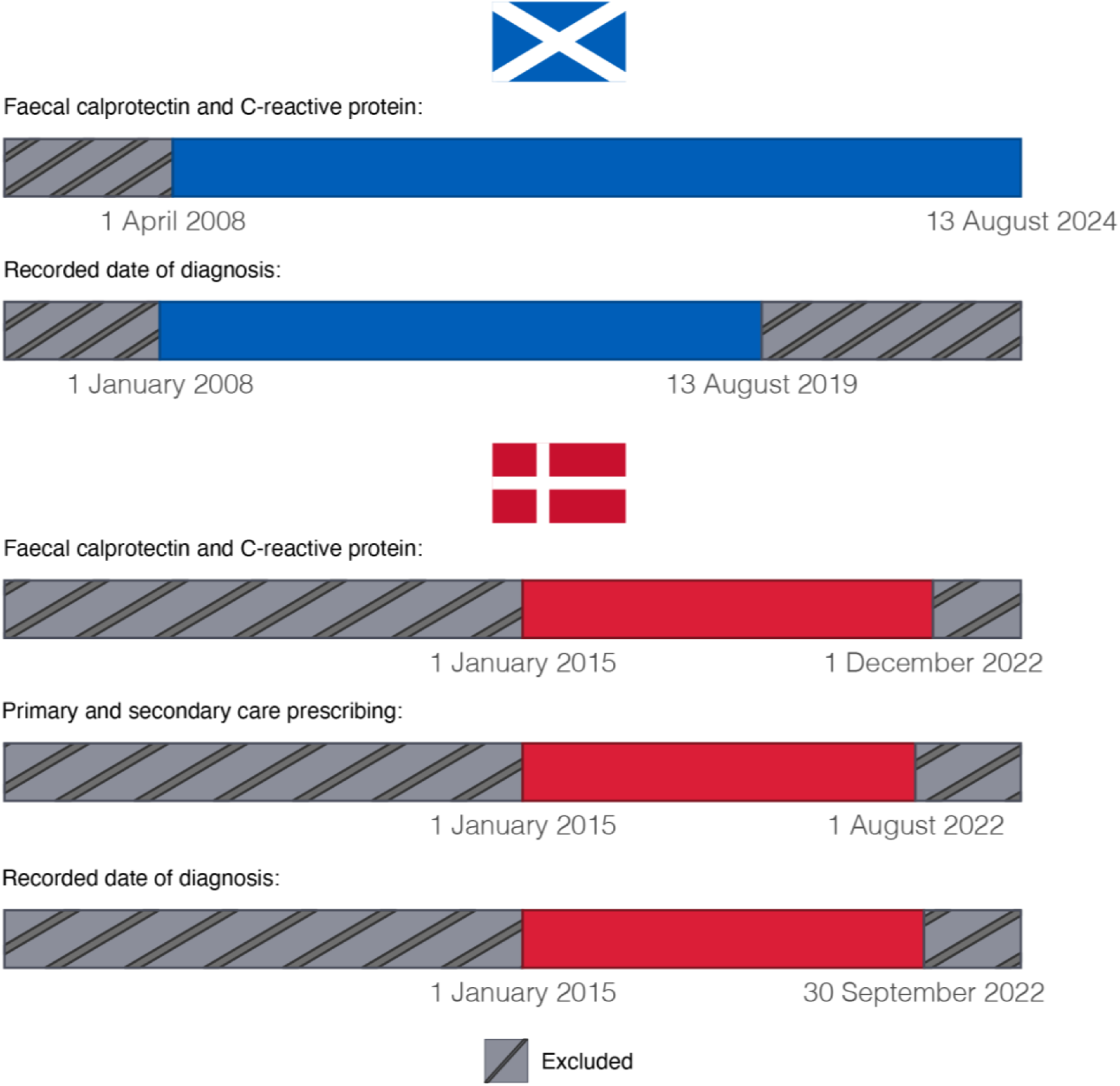
Data availability for the LIBDR (Scotland) and national Danish registry data. Longitudinal biomarker measurements were extracted by the local biochemistry teams. All FC and CRP measurements were extracted by corresponding local biochemistry teams. For LIBDR, the lower bound for diagnosis dates was established as FC testing was not routinely performed in the NHS Lothian health board prior to this date. The upper bound ensured subjects had the possibility of at least five years of follow-up at the time of data extraction. CRP was unavailable prior to April 2008 due to a system migration. For the Danish cohort, the lower bound was set based on when the Danish nationwide Register of Laboratory Results for Research became nationwide. The upper bounds were defined by the date of data extraction.

**Figure S2.**
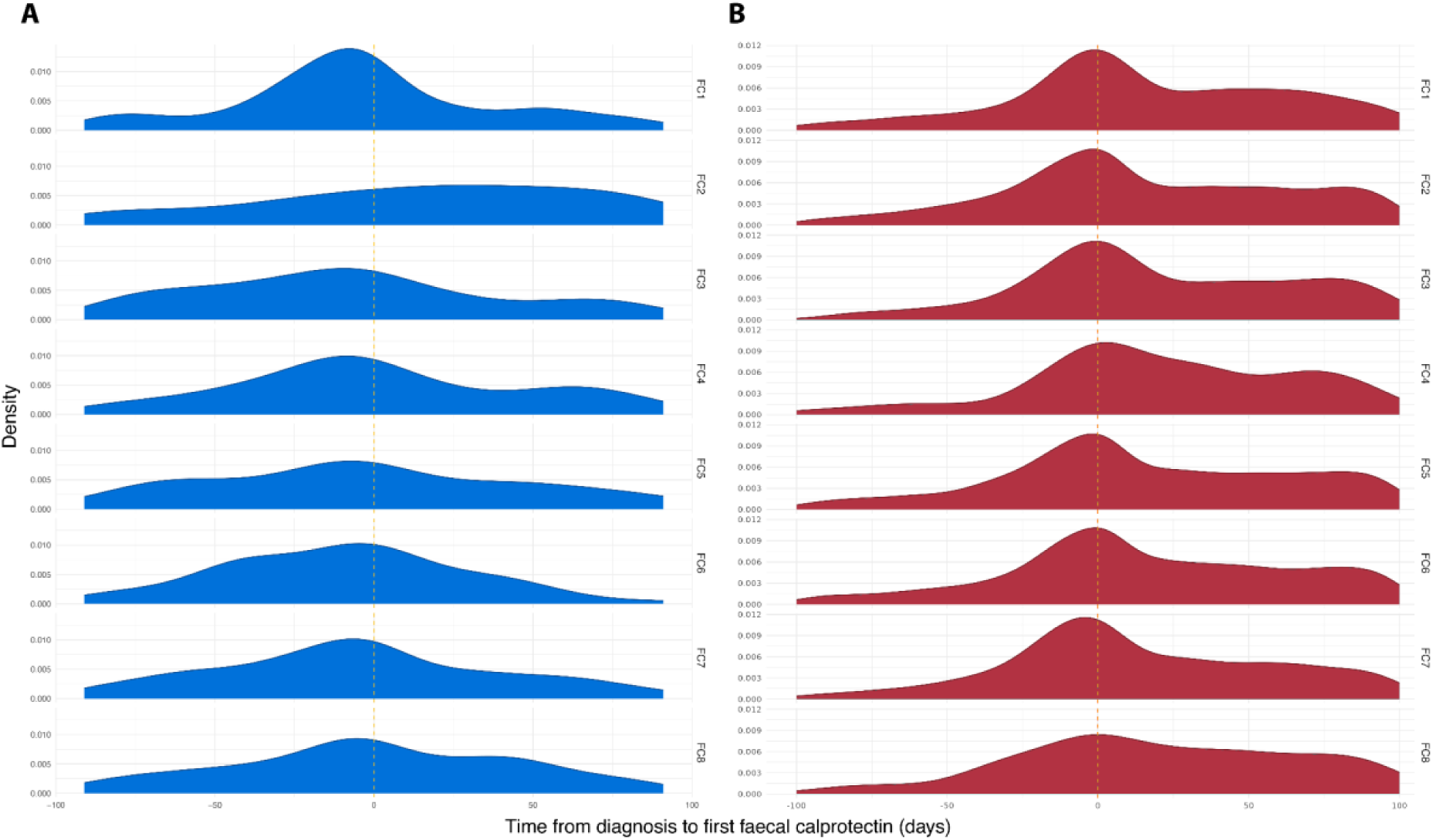
Distribution of observation times for diagnostic faecal calprotectin (FC) relative to reported date of diagnosis for (A) the Lothian IBD registry cohort and (B) the Danish national registry cohort. Stratified by FC cluster assignment. Diagnostic FC was defined as the first FC test within ±90 days of diagnosis.

**Figure S3.**
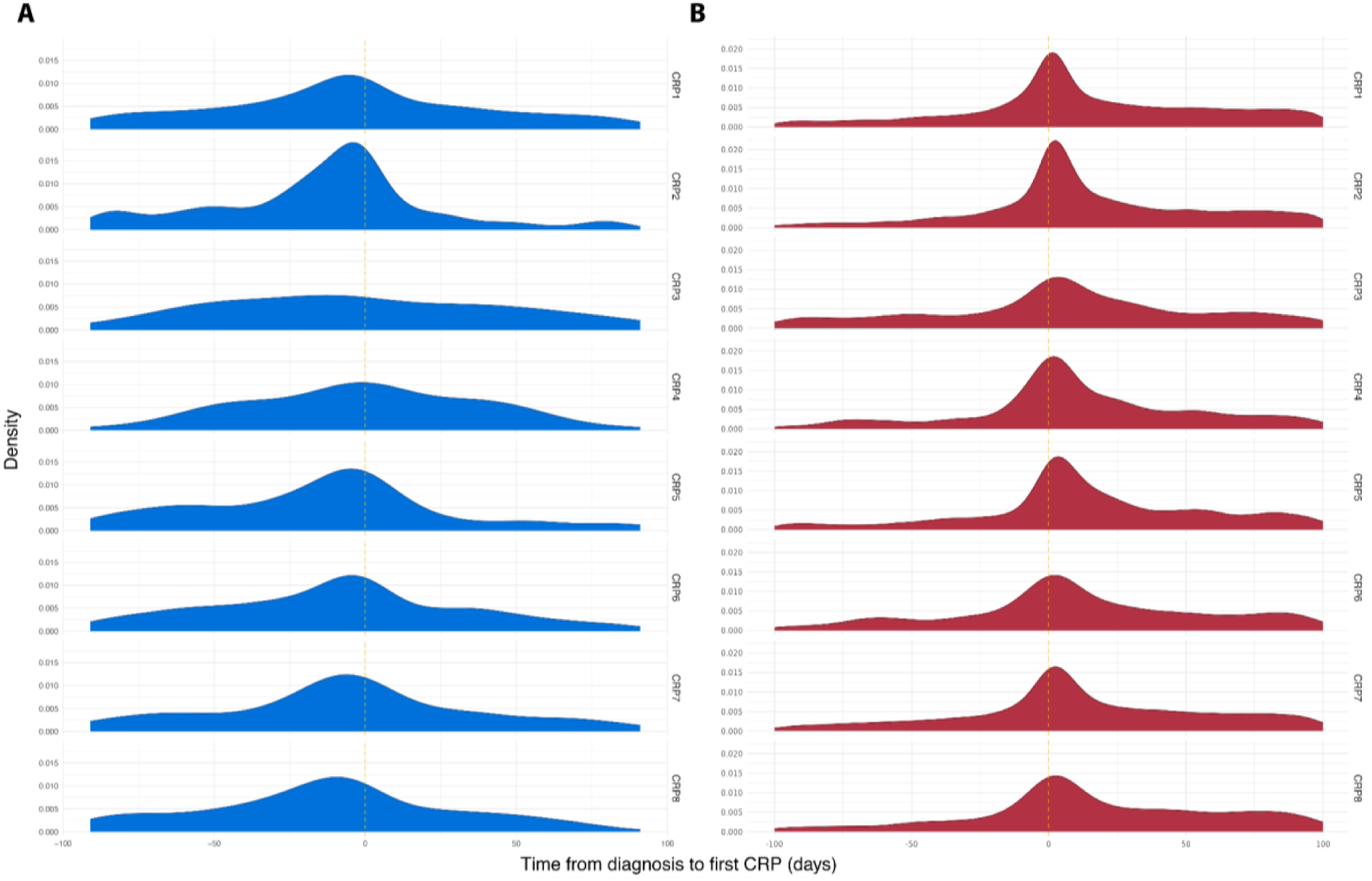
Distribution of observation times for diagnostic C-reactive protein (CRP) relative to reported date of diagnosis for (A) the Lothian IBD registry cohort and (B) the Danish national registry cohort. Stratified by CRP cluster assignment. Diagnostic CRP was defined as the first CRP test within ±90 days of diagnosis.

**Figure S4.**
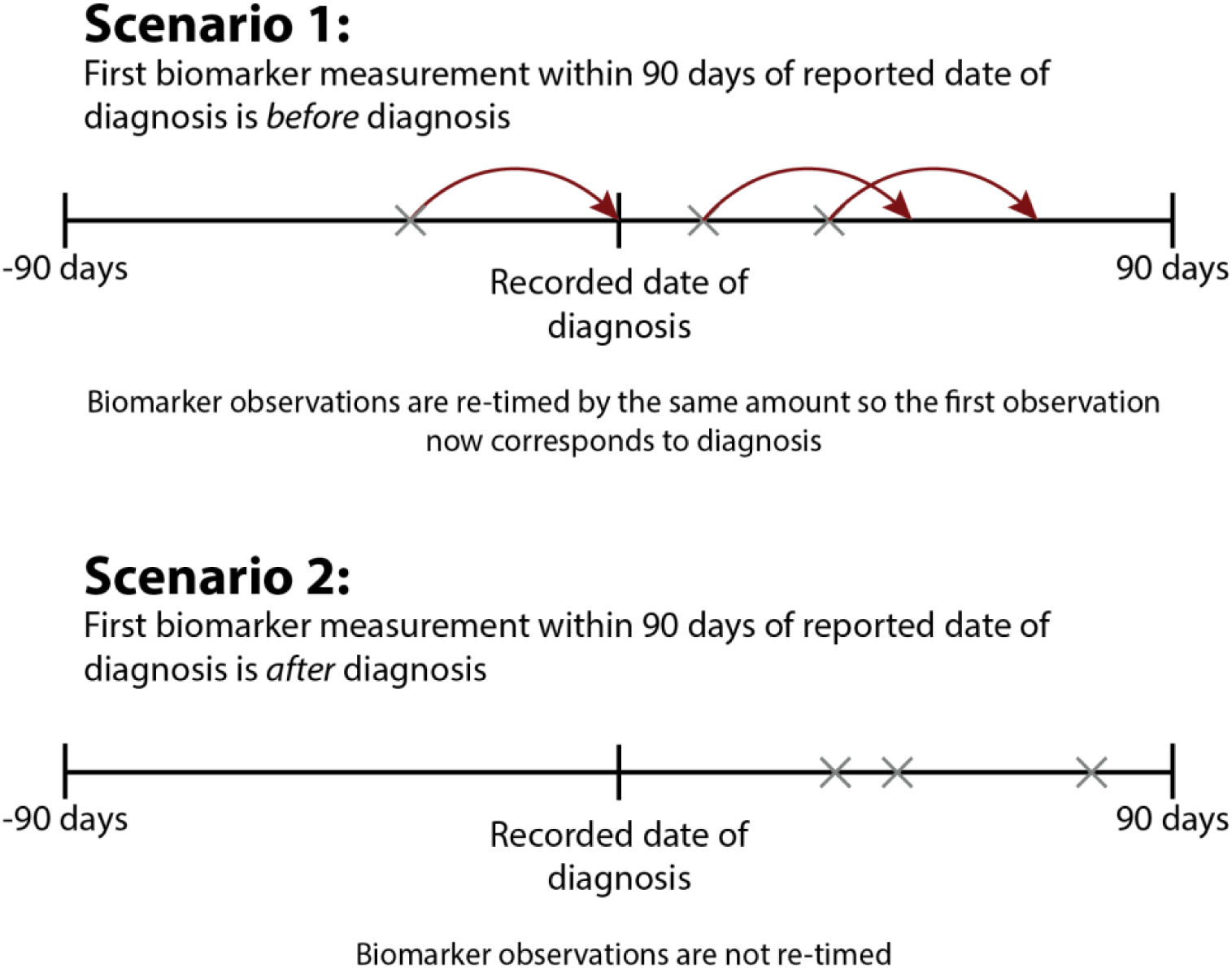
Illustration of how biomarker measurements (FCAL or CRP) were realigned in cases where measurements were observed within three months prior to the recorded diagnosis date. In such cases (Scenario 1), observation times were adjusted such that the timing of the first measurement was treated as the date of diagnosis (t=0 in subsequent analyses). To retain the time lag between observations, the timing of all other observations of that biomarker for the same subject were also adjusted. If an observation is outside the follow-up period after this adjustment, the observation is discarded.

**Figure S5.**
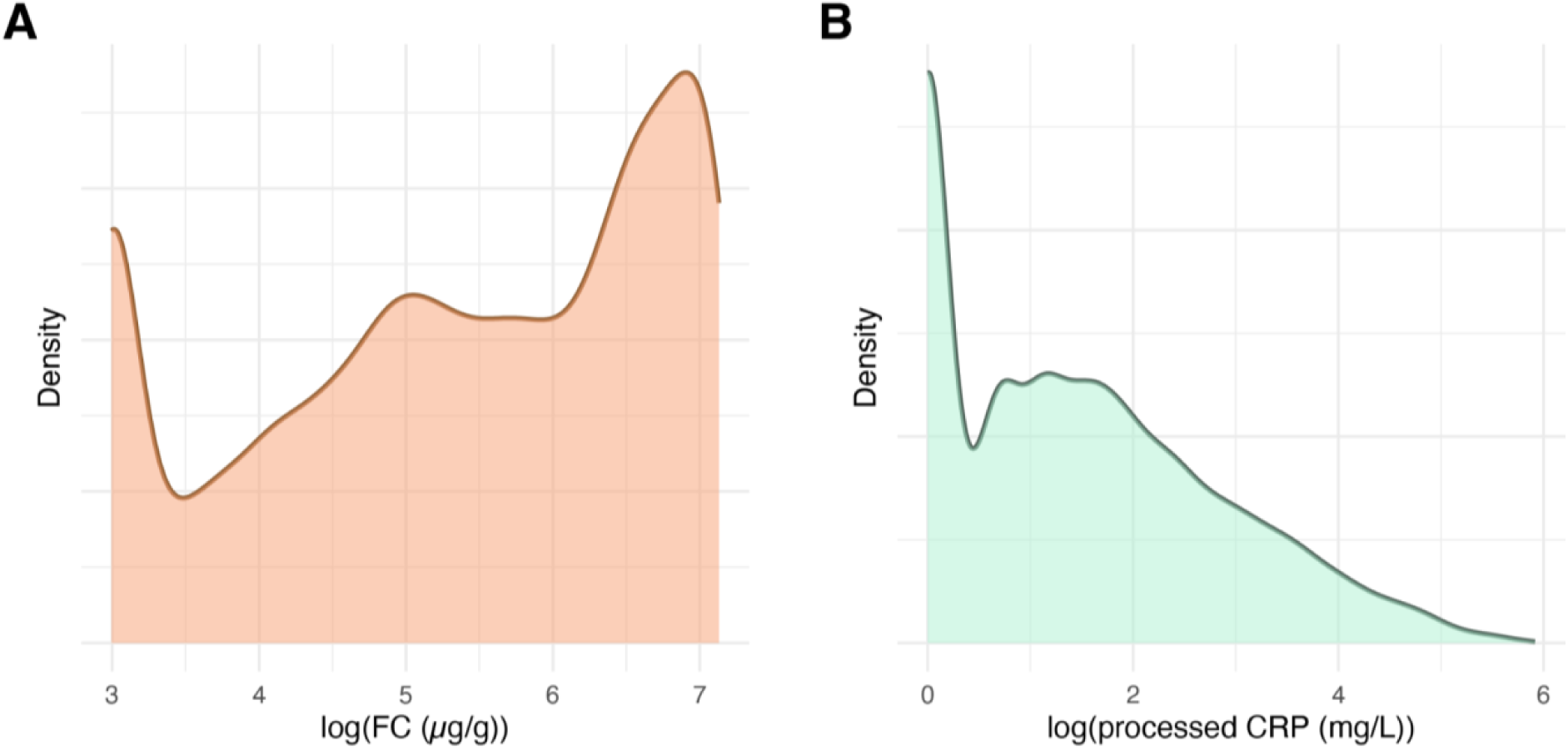
Distribution of diagnostic biomarker measurements across the LIBDR study cohort. (A) faecal calprotectin after applying a logarithmic transformation; (B) pre-processed (grouped into time intervals with the median used for multiple measurements) CRP after logarithmic transformation.

**Figure S6.**
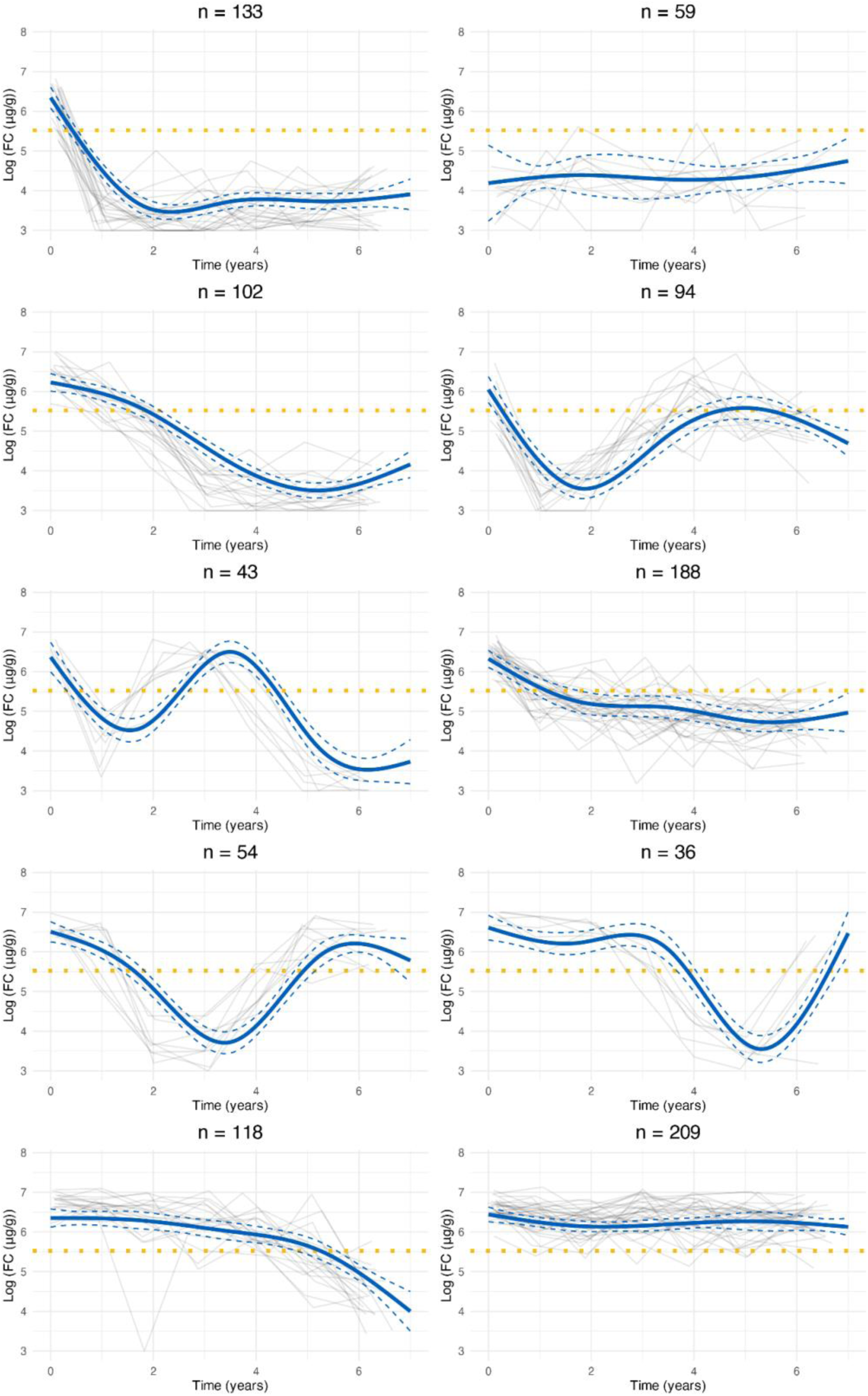
Cluster trajectories obtained from LCMM assuming ten clusters fitted to faecal calprotectin data in the Lothian IBD Registry. Red lines indicate predicted mean cluster profiles with 95% confidence intervals. Dotted horizontal lines indicate log(250μg/g). For visualisation purposes, pseudo subject-specific trajectories have been generated by amalgamating observations from groups of six subjects.

**Figure S7.**
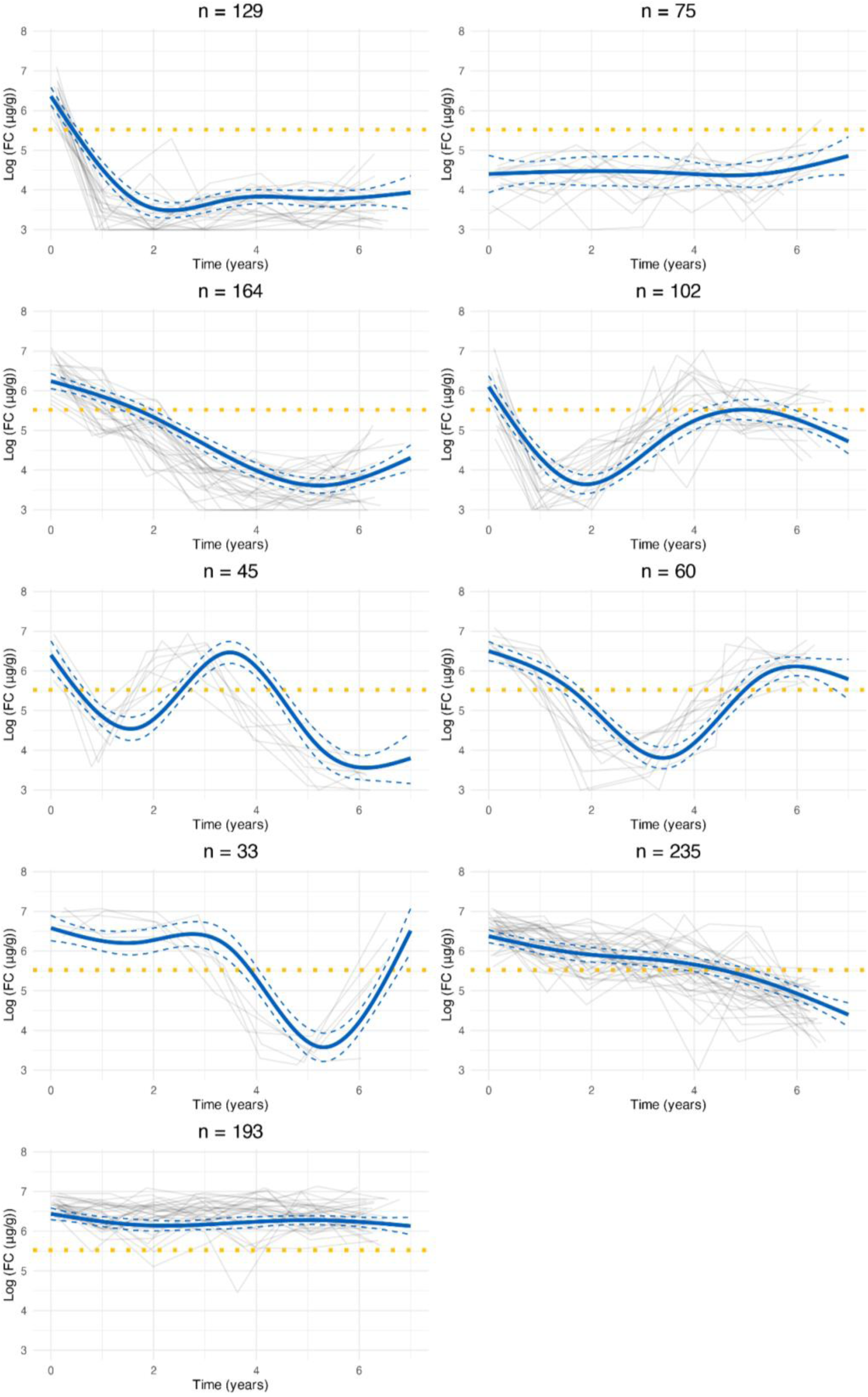
Cluster trajectories obtained from LCMM assuming nine clusters fitted to faecal calprotectin data in the Lothian IBD Registry. Red lines indicate predicted mean cluster profiles with 95% confidence intervals. Dotted horizontal lines indicate log(250μg/g). For visualisation purposes, pseudo subject-specific trajectories have been generated by amalgamating observations from groups of six subjects.

**Figure S8.**
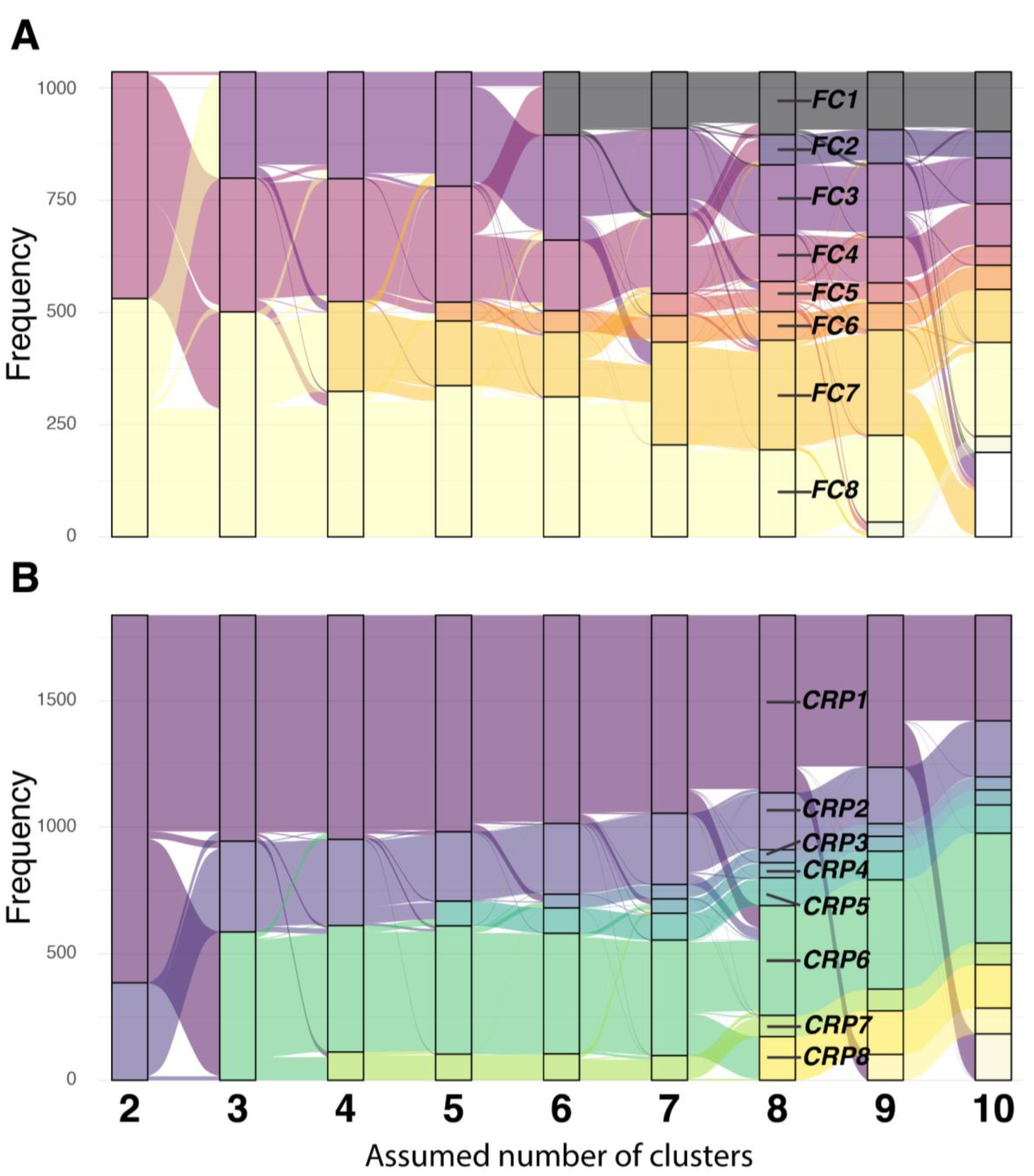
Alluvial plot demonstrating how cluster assignment changes for the Lothian IBD registry cohort as the number of assumed clusters increases for the chosen models for (A) faecal calprotectin and (B) C-reactive protein. The clusters found by the 8-cluster models are labelled.

**Figure S9.**
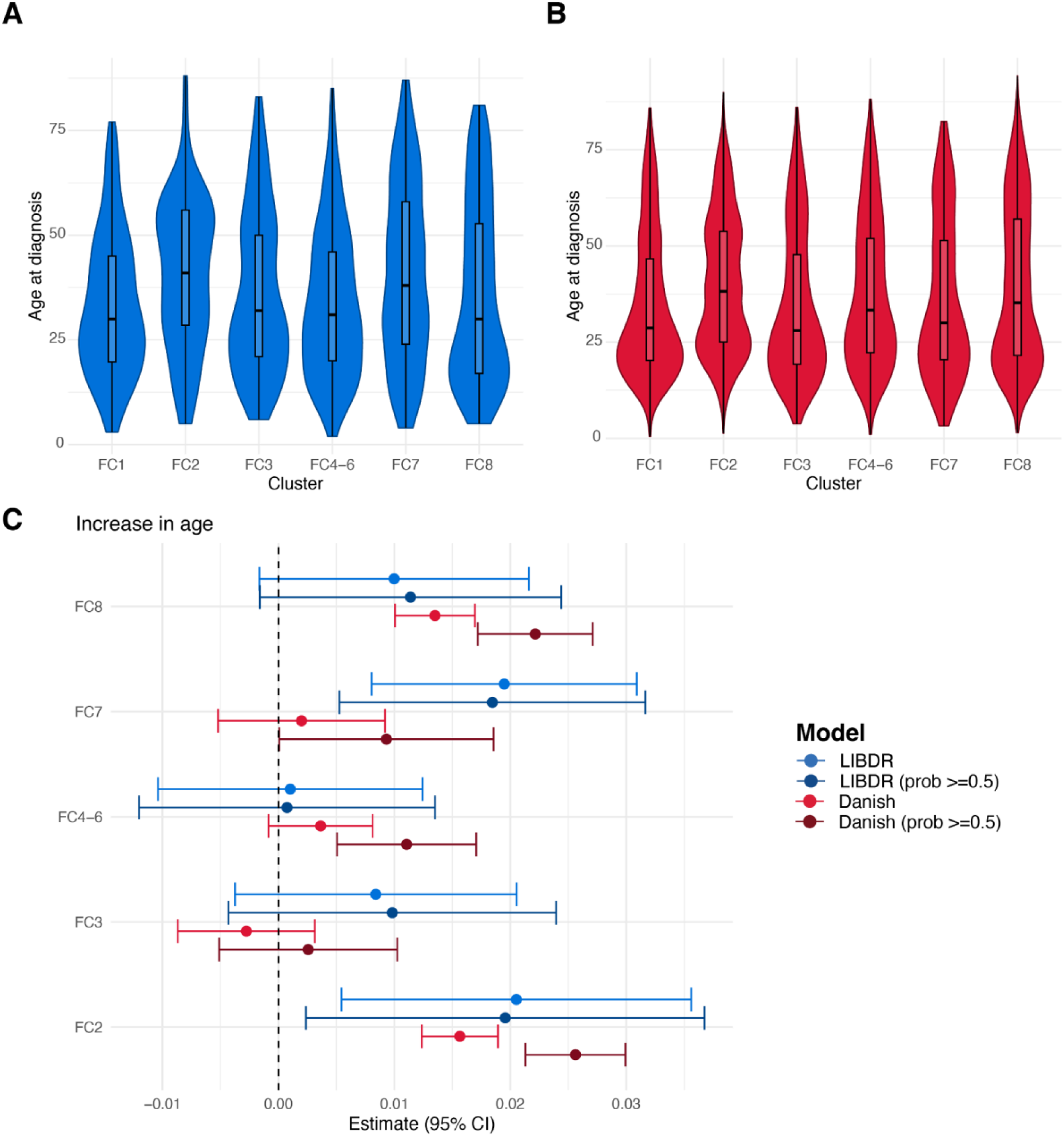
For each FC cluster, violin plots show the distribution of age at diagnosis across subjects, highlighting median and interquartile ranges for (A) the Lothian IBD Registry and (B) Danish national registry data. (C) Forest plot showing the estimated effect sizes and associated 95% confidence intervals for age in a multinomial logistic regression model that uses FC cluster assignment as outcome. Subjects with a posterior probability of belonging to their assigned cluster greater than 0.5 were considered in addition to all subjects in the cohort. Effect sizes are with respect to the reference cluster (in this case FC1). The multivariate model includes age, sex and IBD type as covariates. The dashed vertical lines are used as a reference to indicate no effect.

**Figure S10.**
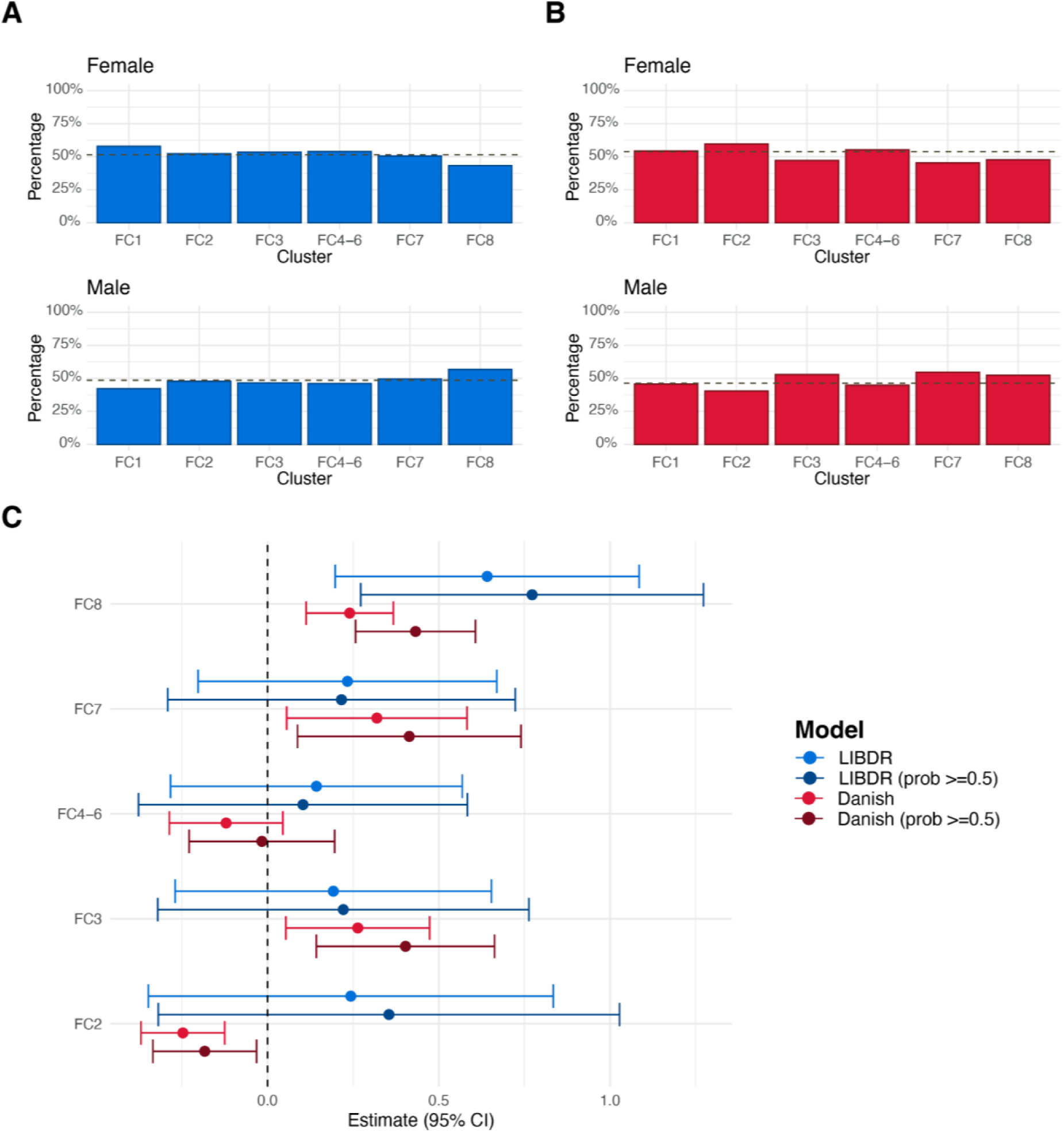
For each FC cluster, barplots show the proportion of individuals with female and male sex. The dashed horizontal line represents overall proportions for (A) the Lothian IBD Registry and (B) Danish national registry data. (C) Forest plot showing the estimated effect sizes and associated 95% confidence intervals for male sex versus females (baseline category) in a multinomial logistic regression model that uses FC cluster assignment as outcome. Subjects with a posterior probability of belonging to their assigned cluster greater than 0.5 were considered in addition to all subjects in the cohort. In (C), effect sizes are with respect to the reference cluster (in this case FC1). The multivariate model includes age, sex and IBD type as covariates. The dashed vertical line is used as a reference to indicate no effect.

**Figure S11.**
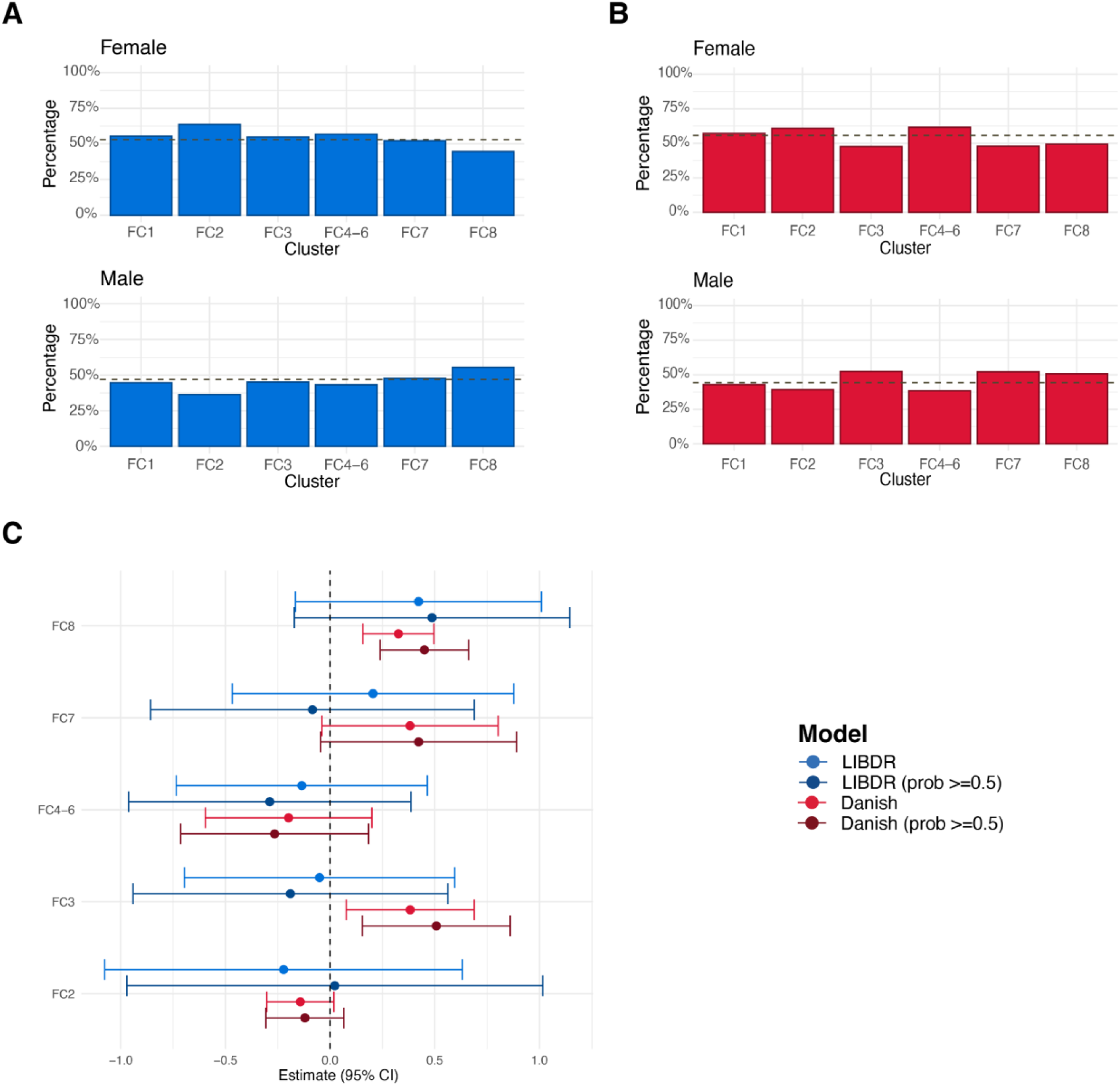
Crohn’s disease subjects only. For each FC cluster, barplots show the proportion of individuals with female and male sex. The dashed horizontal line represents overall proportions for (A) the Lothian IBD Registry and (B) Danish national registry data. (C) Forest plot showing the estimated effect sizes and associated 95% confidence intervals for male sex versus females (baseline category) in a multinomial logistic regression model that uses FC cluster assignment as outcome. Subjects with a posterior probability of belonging to their assigned cluster greater than 0.5 were considered in addition to all subjects in the cohort, In (C), effect sizes are with respect to the reference cluster (in this case FC1). The multivariate model includes age, sex and IBD type as covariates. The dashed vertical lines are used as a reference to indicate no effect.

**Figure S12.**
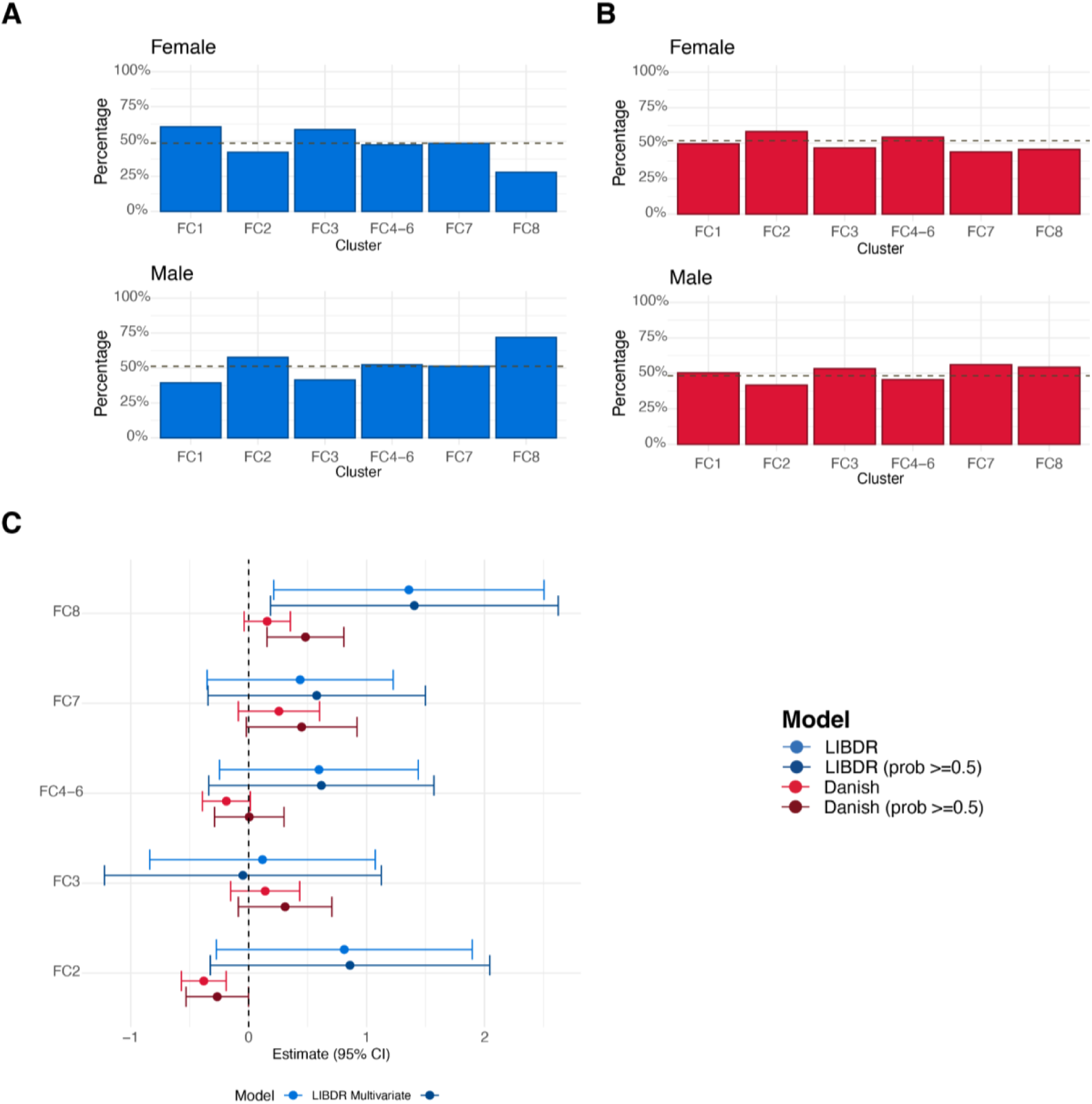
Ulcerative colitis subjects only. For each FC cluster, barplots show the proportion of individuals with female and male sex. The dashed horizontal line represents overall proportions for (A) the Lothian IBD Registry and (B) Danish national registry data. (C) Forest plot showing the estimated effect sizes and associated 95% confidence intervals for male sex versus females (baseline category) in a multinomial logistic regression model that uses FC cluster assignment as outcome. Subjects with a posterior probability of belonging to their assigned cluster greater than 0.5 were considered in addition to all subjects in the cohort. In (C), effect sizes are with respect to the reference cluster (in this case FC1). The multivariate model includes age, sex and IBD type as covariates. The dashed vertical lines are used as a reference to indicate no effect.

**Figure S13.**
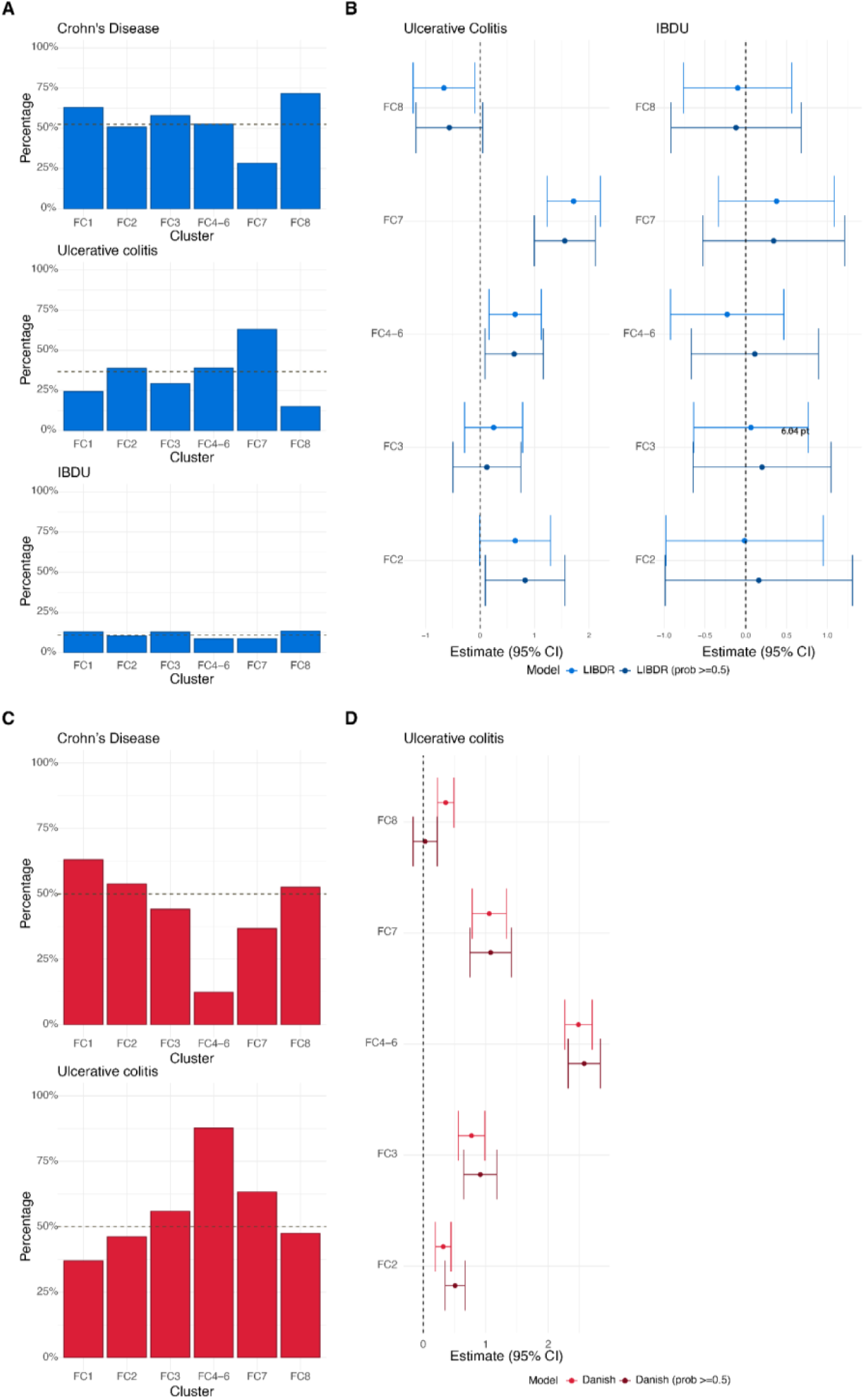
(A) For each FC cluster, panels show the proportion of individuals with Crohn’s disease, ulcerative colitis and IBDU respectively in the Lothian IBD Registry. (B) Forest plot showing the estimated effect sizes and associated 95% confidence intervals for IBD type in the Lothian IBD Registry: ulcerative colitis and IBDU versus Crohn’s disease (baseline category) in a multinomial logistic regression model that uses FC cluster assignment as outcome. (C) For each FC cluster, panels show the proportion of individuals with Crohn’s disease and ulcerative colitis respectively in the Danish cohort. (D) Forest plot showing the estimated effect sizes and associated 95% confidence intervals for IBD type in the Danish cohort: ulcerative colitis versus Crohn’s disease (baseline category) in a multinomial logistic regression model that uses FC cluster assignment as outcome. There are no IBDU diagnoses reported for the Danish cohort as these were mapped to either CD or UC (Supplementary Note 1). Subjects with a posterior probability of belonging to their assigned cluster greater than 0.5 were considered in addition to all subjects in the cohort. In (A) and (C), the dashed horizontal line represents overall proportions across the FC cohorts. In (B) and (D), effect sizes are with respect to the reference cluster (in this case FC1). In both cases, the multivariate model includes age, sex and IBD type as covariates. The dashed vertical lines are used as a reference to indicate no effect.

**Figure S14.**
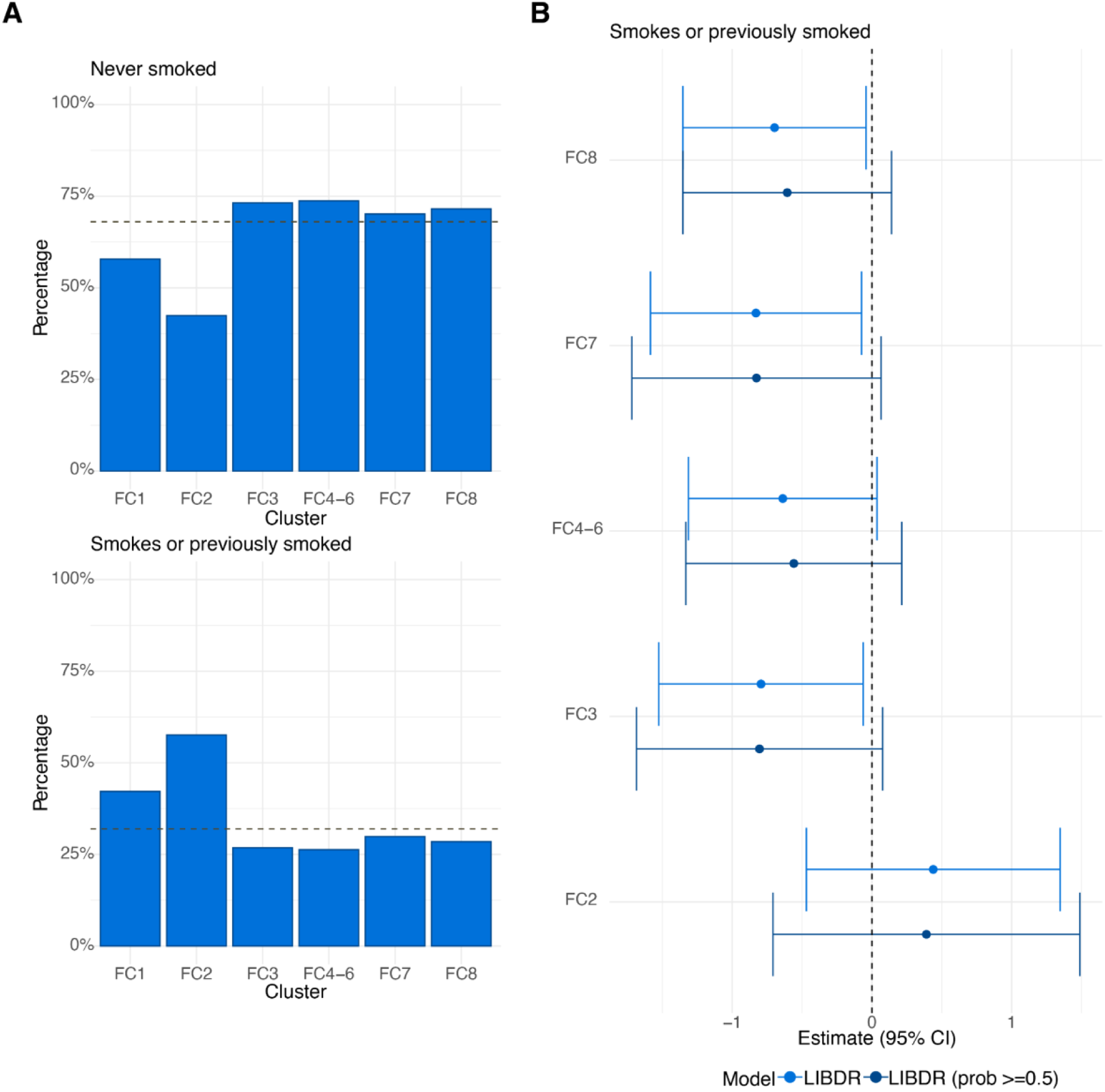
Crohn’s disease patients in the Lothian IBD Registry only. (A) For each FC cluster, panels show the proportion of individuals with smoking behaviour recorded as “no” (no and never) and “yes” (current or previously smoked) at diagnosis respectively. The dashed horizontal line represents overall proportions across the entire FC cohort. (B) Forest plot showing the estimated effect sizes and associated 95% confidence intervals for smoking: yes versus no (baseline category) in a multinomial logistic regression model that uses FC cluster assignment as outcome. Subjects with a posterior probability of belonging to their assigned cluster greater than 0.5 were considered in addition to all subjects in the cohort, Effect sizes are with respect to the reference cluster (in this case FC1). The multivariate model includes age, sex, smoking, Montreal location (L1, L2, L3), upper gastrointestinal inflammation (L4), perianal disease (yes, no) and Montreal behaviour (B1, B2/B3) as covariates. The dashed vertical lines are used as a reference to indicate no effect.

**Figure S15.**
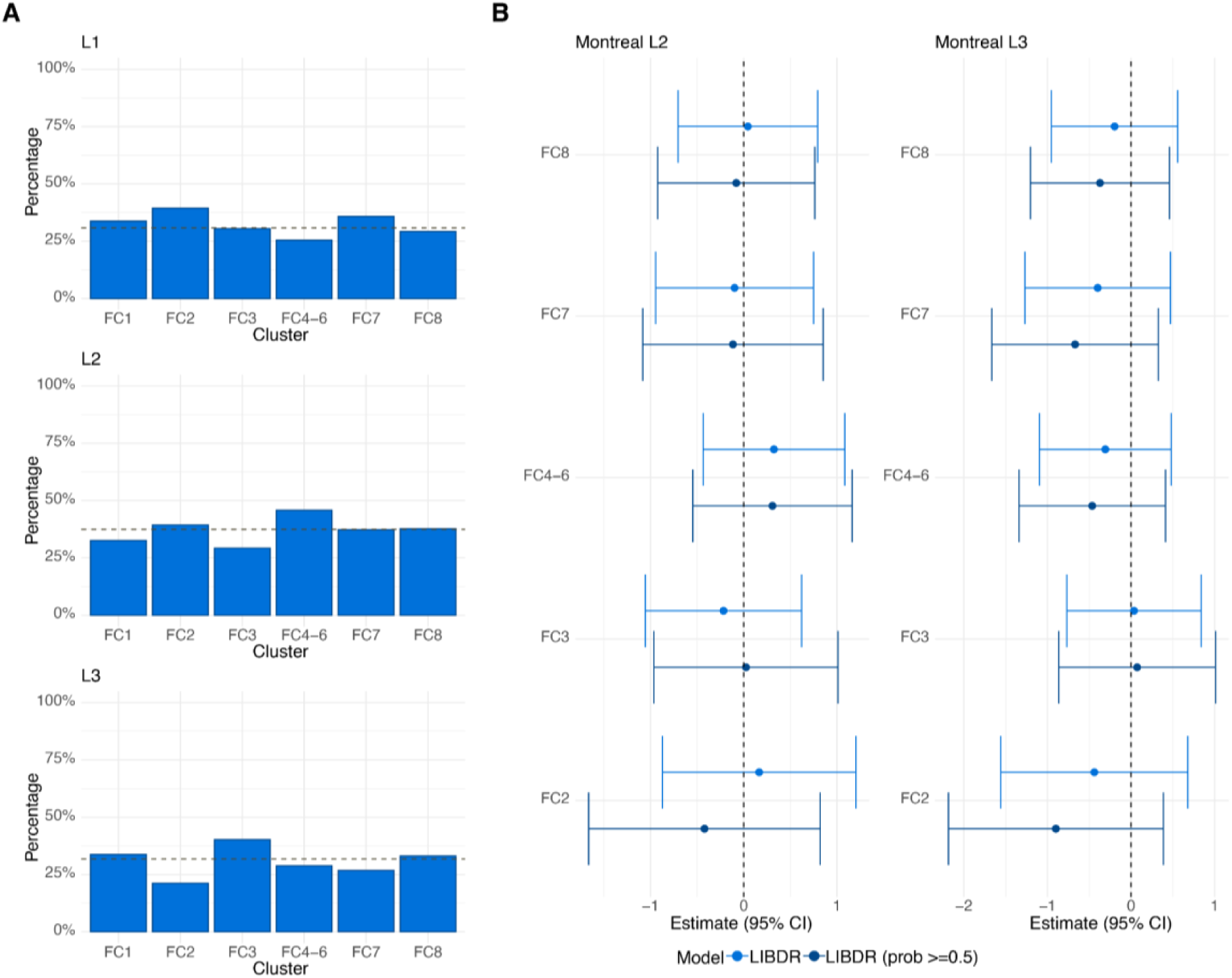
Crohn’s disease patients in the Lothian IBD Registry only. (A) For each FC cluster, panels show the proportion of individuals with Montreal Location recorded as L1, L2 and L3 respectively. The dashed horizontal line represents overall proportions across the entire FC cohort. (B) Forest plot showing the estimated effect sizes and associated 95% confidence intervals for Montreal Location: L2 and L3 versus L1 (baseline category) in a multinomial logistic regression model that uses FC cluster assignment as outcome. Subjects with a posterior probability of belonging to their assigned cluster greater than 0.5 were considered in addition to all subjects in the cohort. Effect sizes are with respect to the reference cluster (in this case FC4). The multivariate model includes age, sex, smoking, Montreal location (L1, L2, L3), upper gastrointestinal inflammation (L4), perianal disease (yes, no) and Montreal behaviour (B1, B2/B3) as covariates. The dashed vertical lines are used as a reference to indicate no effect.

**Figure S16.**
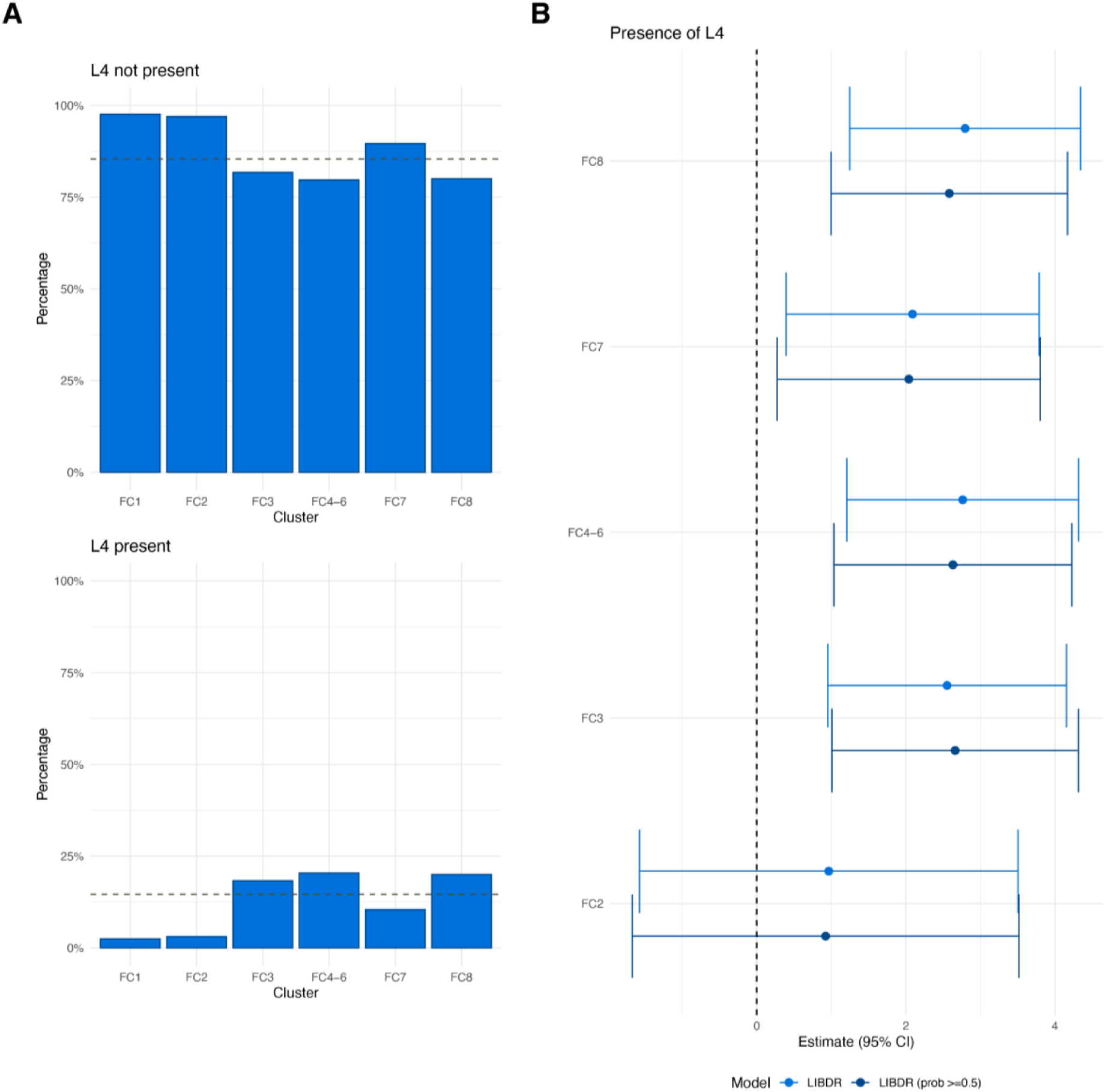
Crohn’s disease patients in the Lothian IBD Registry only. (A) For each FC cluster, panels show the proportion of individuals with Montreal L4 (upper gastrointestinal inflammation), recorded as present or non present. The dashed horizontal line represents overall proportions across the entire FC cohort. (B) Forest plot showing the estimated effect sizes and associated 95% confidence intervals for L4: present versus not present (baseline category) in a multinomial logistic regression model that uses FC cluster assignment as outcome. Subjects with a posterior probability of belonging to their assigned cluster greater than 0.5 were considered in addition to all subjects in the cohort. Effect sizes are with respect to the reference cluster (in this case FC1). The multivariate model includes age, sex, smoking, Montreal location (L1, L2, L3), upper gastrointestinal inflammation (L4), perianal disease (yes, no) and Montreal behaviour (B1, B2/B3) as covariates. The dashed vertical lines are used as a reference to indicate no effect.

**Figure S17.**
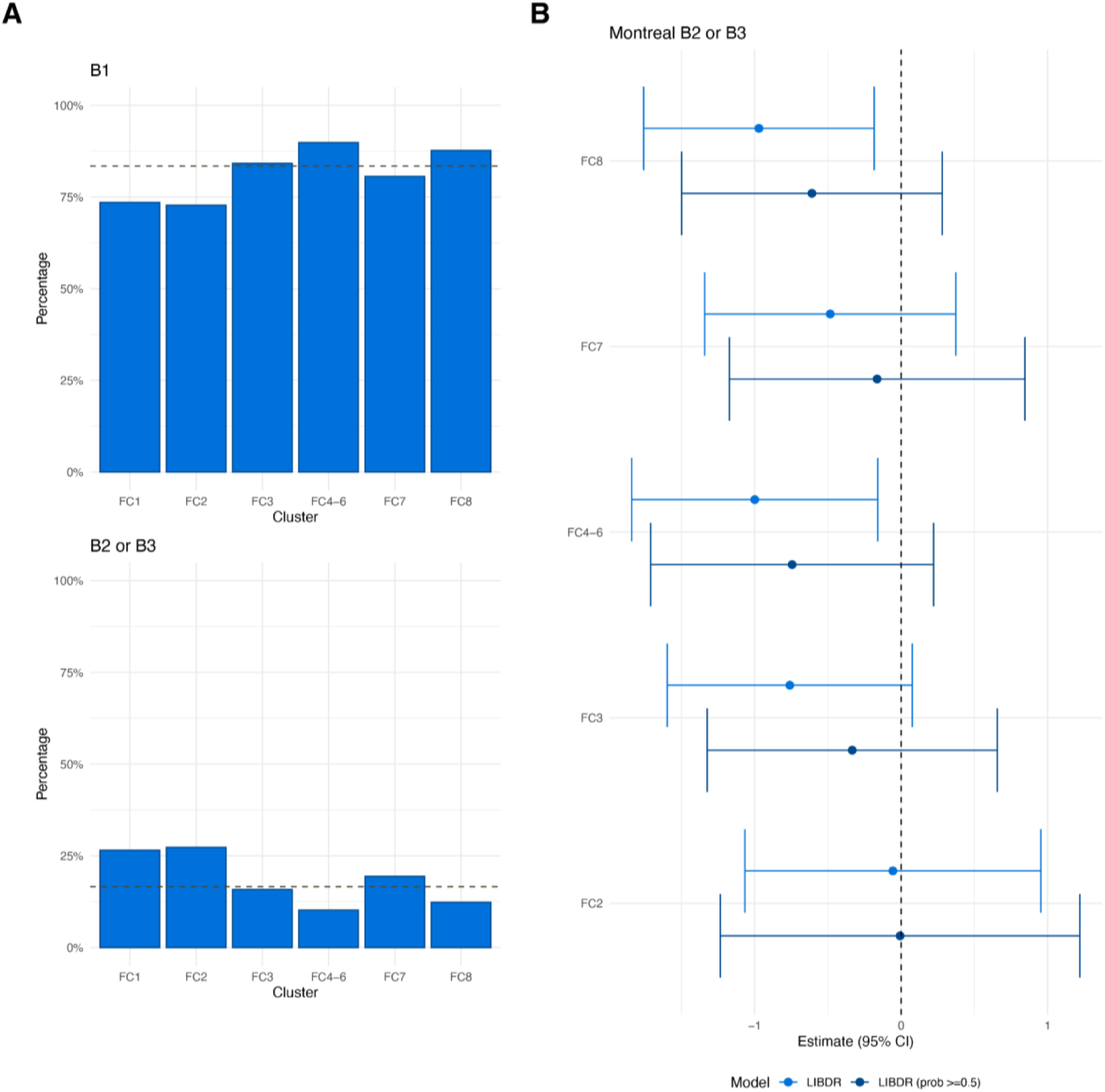
Crohn’s disease patients in the Lothian IBD Registry only. (A) For each FC cluster, panels show the proportion of individuals with Montreal behaviour recorded as B1 or B2/B3 respectively. The dashed horizontal line represents overall proportions across the entire FC cohort. (B) Forest plot showing the estimated effect sizes and associated 95% confidence intervals for Montreal behaviour: B2/B3 versus B1 (baseline category) in a multinomial logistic regression model that uses FC cluster assignment as outcome. Subjects with a posterior probability of belonging to their assigned cluster greater than 0.5 were considered in addition to all subjects in the cohort. Effect sizes are with respect to the reference cluster (in this case FC4). The multivariate model includes age, sex, smoking, Montreal location (L1, L2, L3), upper gastrointestinal inflammation (L4), perianal disease (yes, no) and Montreal behaviour (B1, B2/B3) as covariates. The dashed vertical lines are used as a reference to indicate no effect.

**Figure S18.**
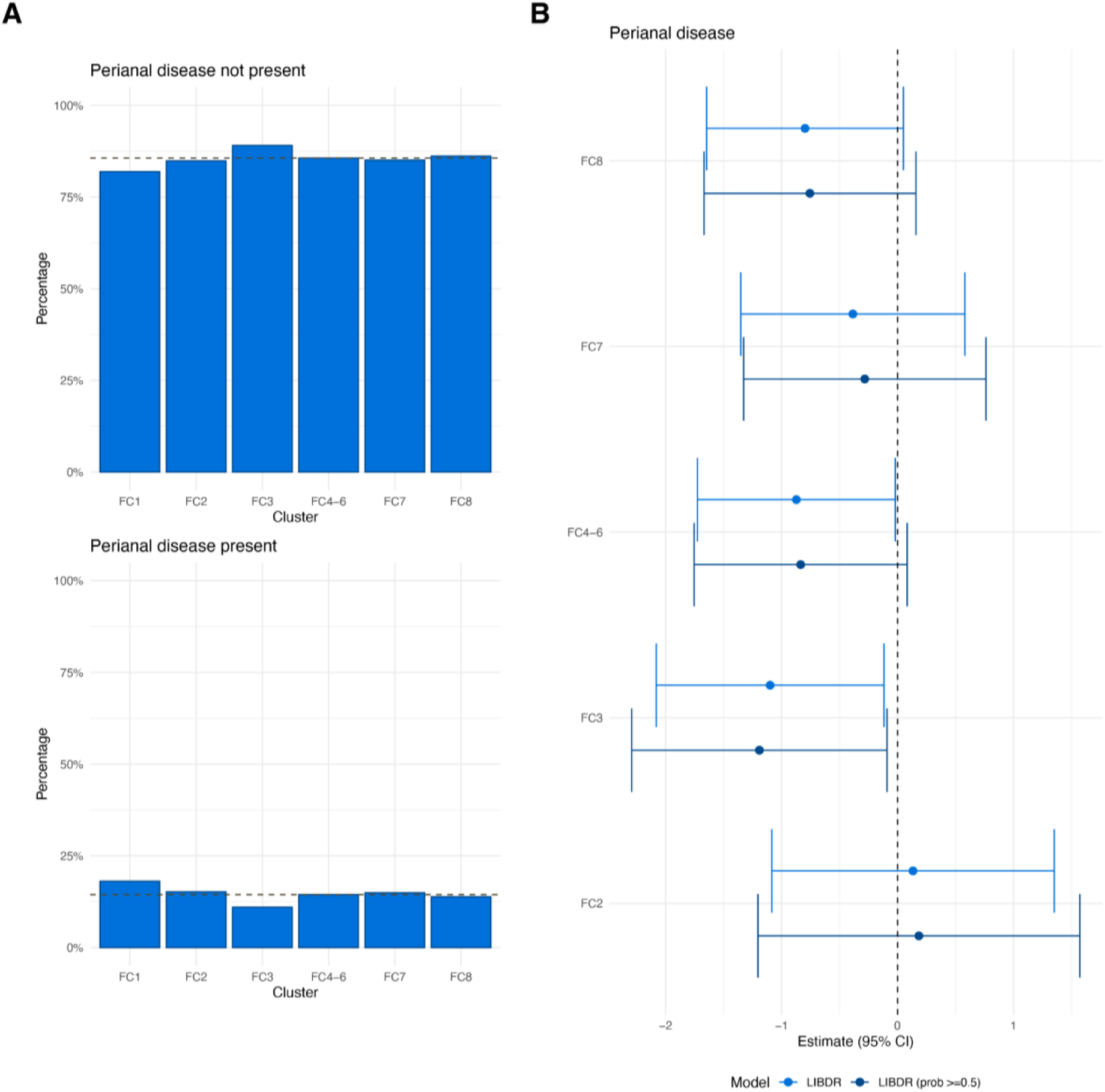
Crohn’s disease patients in the Lothian IBD Registry only. (A) For each FC cluster, panels show the proportion of individuals with perianal disease recorded as present or not present. The dashed horizontal line represents overall proportions across the entire FC cohort. (B) Forest plot showing the estimated effect sizes and associated 95% confidence intervals for perianal disease: present versus not present (baseline category) in a multinomial logistic regression model that uses FC cluster assignment as outcome. Subjects with a posterior probability of belonging to their assigned cluster greater than 0.5 were considered in addition to all subjects in the cohort. Effect sizes are with respect to the reference cluster (in this case FC1). The multivariate model includes age, sex, smoking, Montreal location (L1, L2, L3), upper gastrointestinal inflammation (L4), perianal disease (yes, no) and Montreal behaviour (B1, B2/B3) as covariates. The dashed vertical lines are used as a reference to indicate no effect.

**Figure S19.**
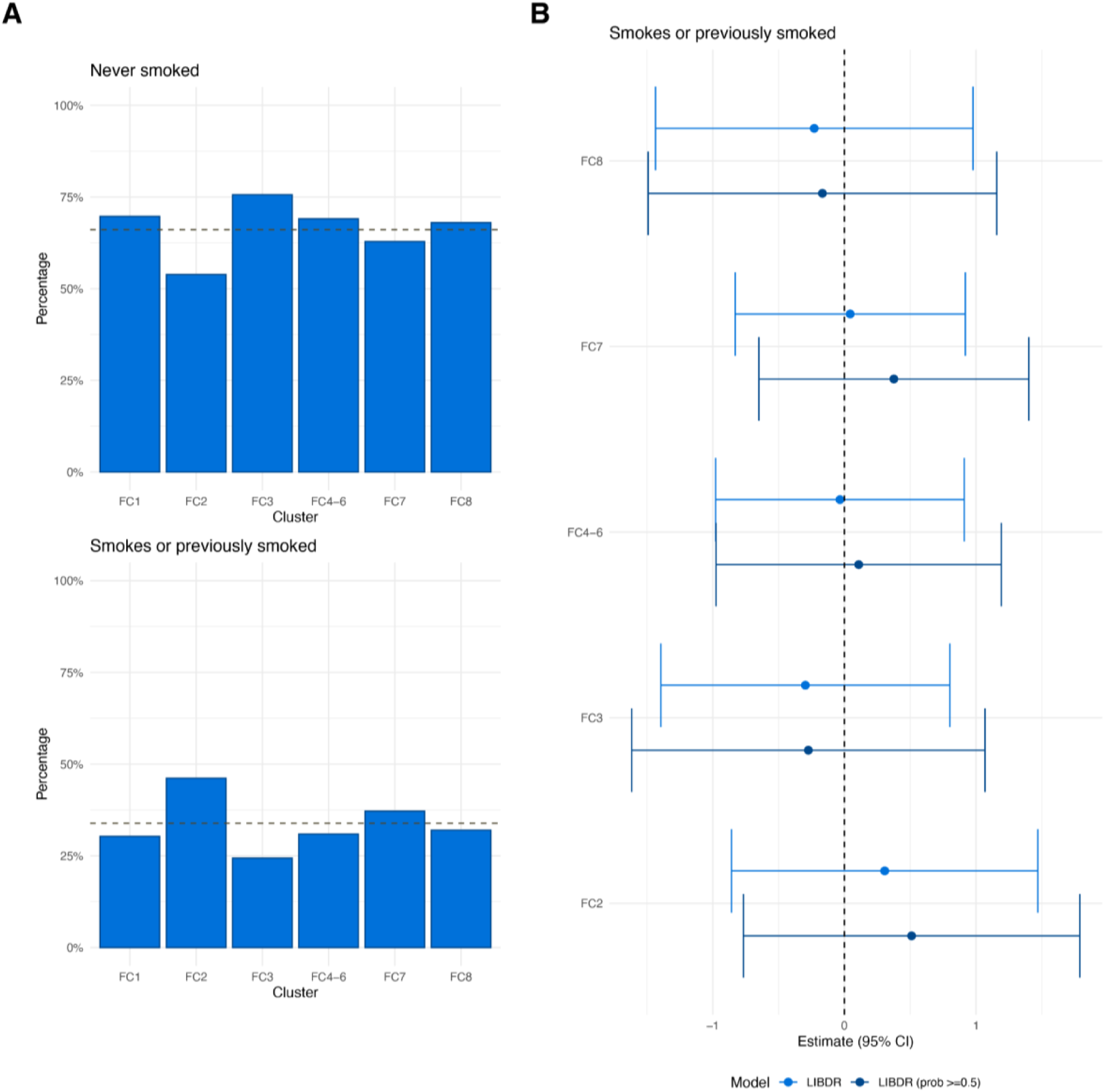
Ulcerative colitis patients in the Lothian IBD Registry only. (A) For each FC cluster, panels show the proportion of individuals with smoking behaviour recorded as “no” (no and never) and “yes” (current or previously smoked) at diagnosis respectively. The dashed horizontal line represents overall proportions across the entire FC cohort. (B) Forest plot showing the estimated effect sizes and associated 95% confidence intervals for smoking: yes versus no (baseline category) in a multinomial logistic regression model that uses FC cluster assignment as outcome. Subjects with a posterior probability of belonging to their assigned cluster greater than 0.5 were considered in addition to all subjects in the cohort. Effect sizes are with respect to the reference cluster (in this case FC1). The multivariate model includes age, sex, smoking and Montreal extent (E1, E2, E3). The dashed vertical lines are used as a reference to indicate no effect.

**Figure S20.**
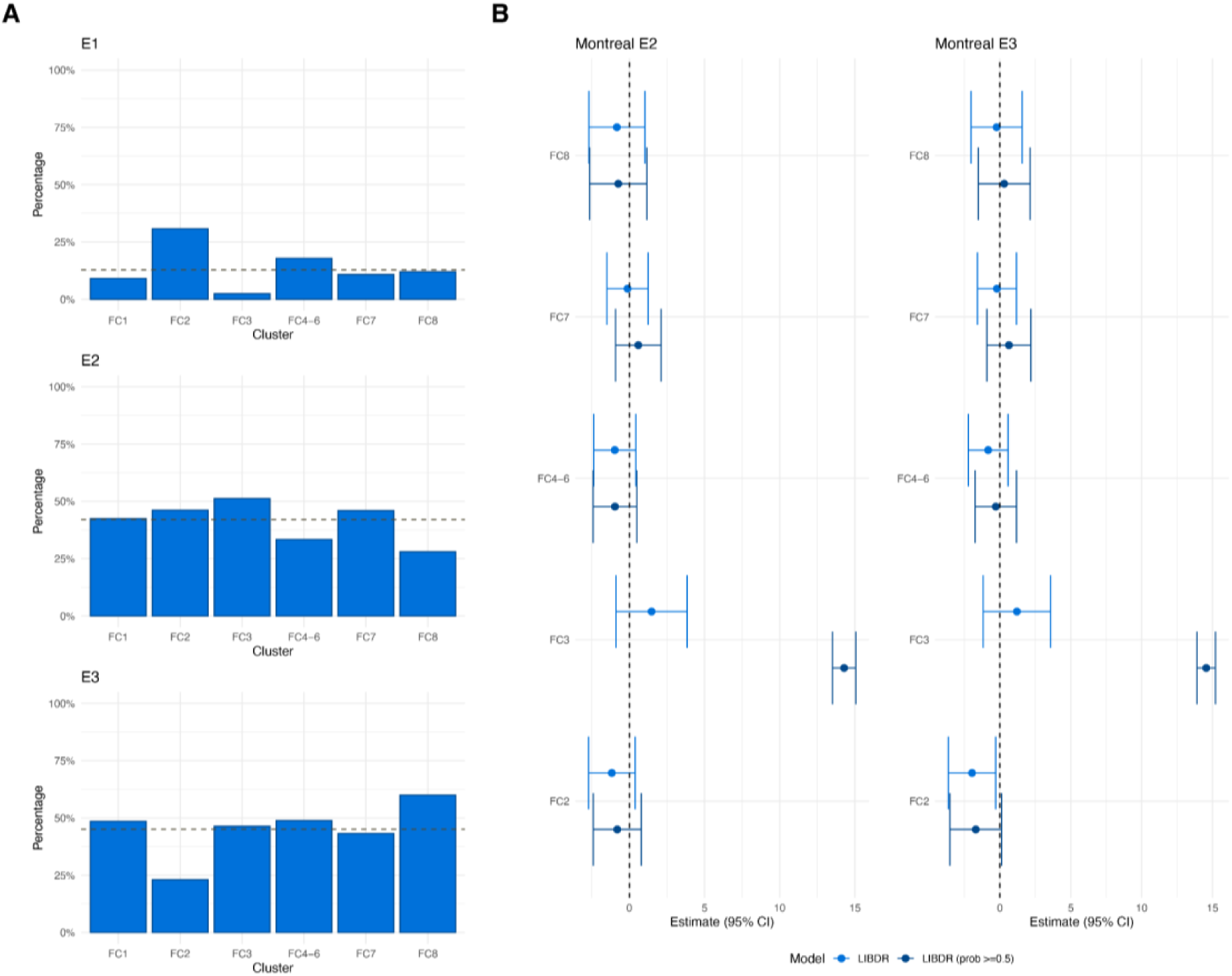
Ulcerative colitis patients in the Lothian IBD Registry only. (A) For each cluster, panels show the proportion of individuals with Montreal extent recorded as E1, E2 and E3 respectively. The dashed horizontal line represents overall proportions across the entire FC cohort. (B) Forest plot showing the estimated effect sizes and associated 95% confidence intervals for Montreal extent: E2 and E3 versus E1 (baseline category) in a multinomial logistic regression model that uses FC cluster assignment as outcome. Subjects with a posterior probability of belonging to their assigned cluster greater than 0.5 were considered in addition to all subjects in the cohort. Effect sizes are with respect to the reference cluster (in this case FC1). The multivariate model includes age, sex, smoking and Montreal extent (E1, E2, E3). The dashed vertical lines are used as a reference to indicate no effect.

**Figure S21.**
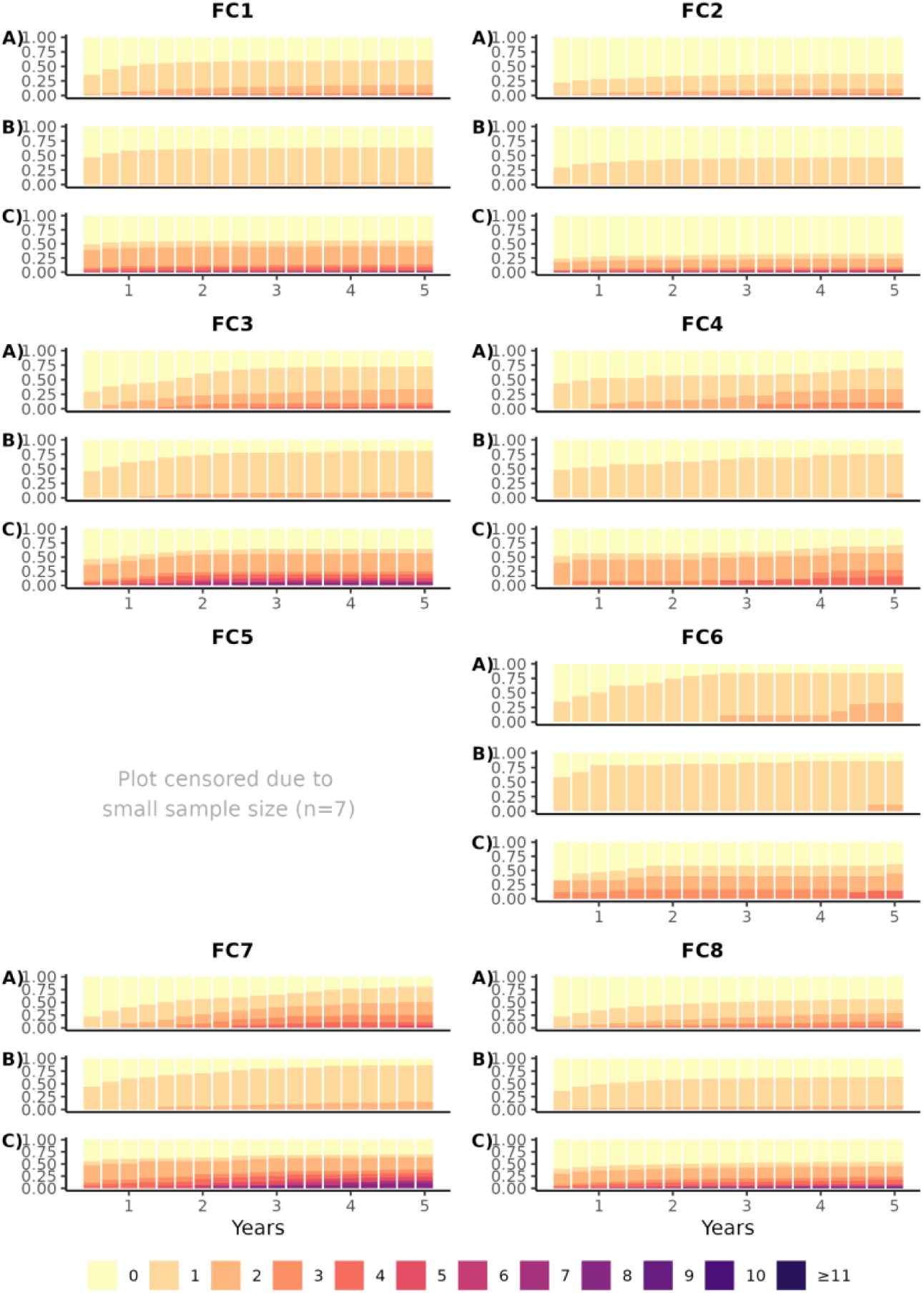
Prescribing trends over time for Crohn’s disease subjects in the Danish faecal calprotectin cohort for (A) commencement of an advanced therapy, (B) commencement of an immunomodulator, and (C) a course of corticosteroids (defined as 1500mg total). Subjects are stratified by FC cluster assignment. For confidentiality reasons, categories with <5 individuals at each time interval were merged with the category with one fewer treatment.

**Figure S22.**
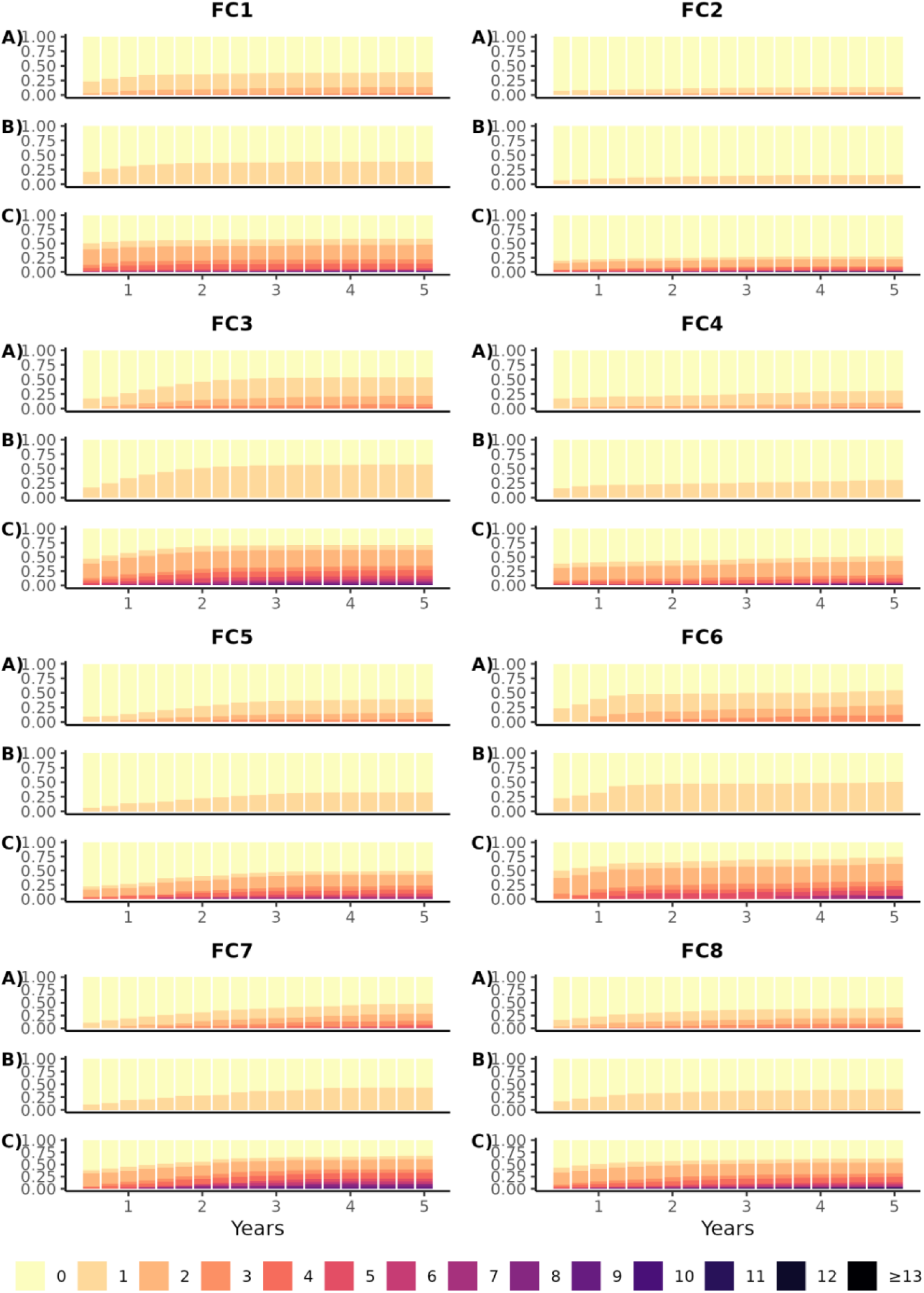
Prescribing trends over time for ulcerative colitis subjects in the Danish faecal calprotectin cohort for (A) commencement of an advanced therapy, (B) commencement of an immunomodulator, and (C) a course of corticosteroids (defined as 1500mg total). Subjects are stratified by FC cluster assignment. For confidentiality reasons, categories with <5 individuals at each time interval were merged with the category with one fewer treatment.

**Figure S23.**
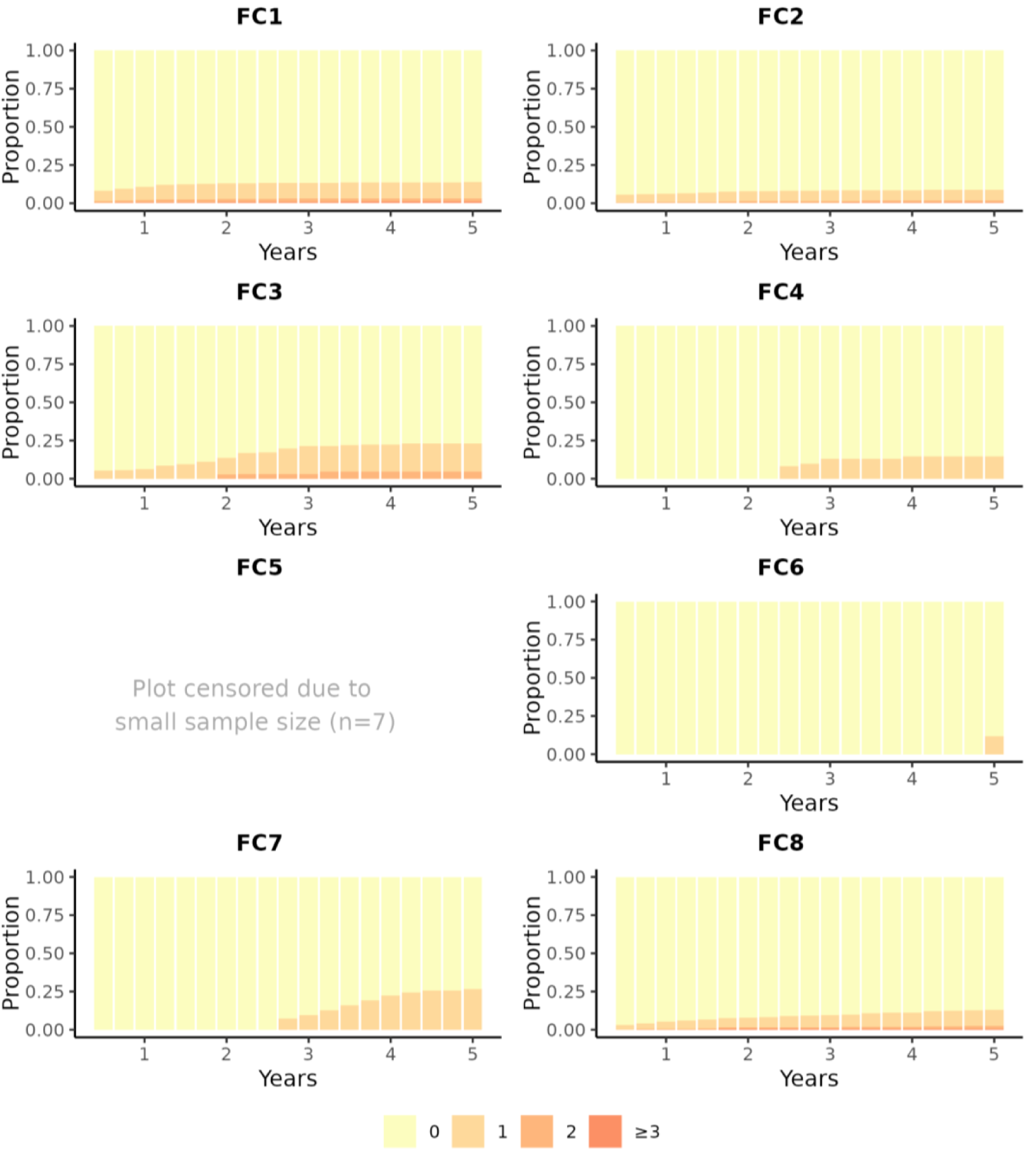
Number of surgical bowel resections over time for Crohn’s disease subjects in the Danish cohort. Stratified by faecal calprotectin cluster assignment.

**Figure S24.**
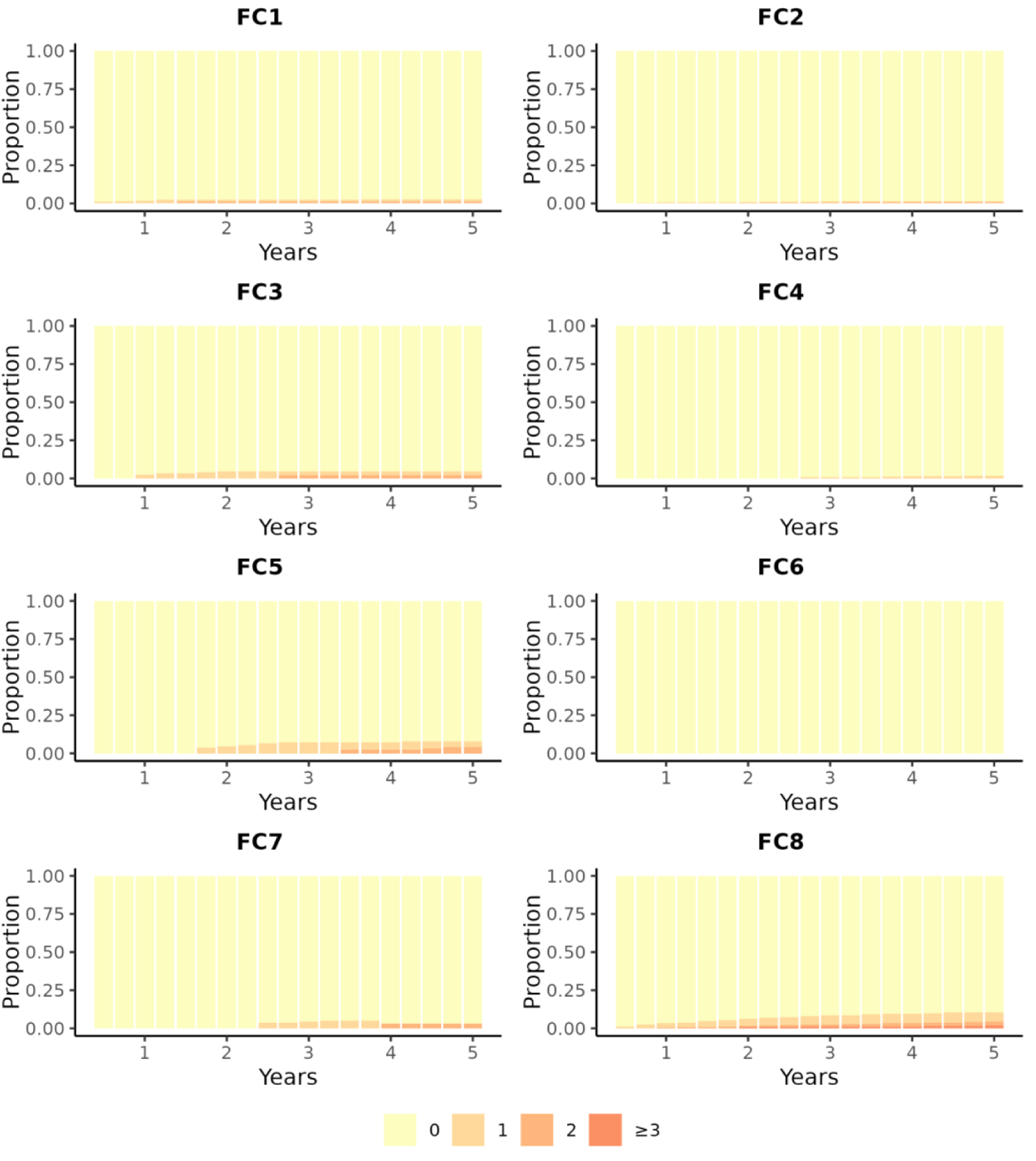
Number of surgical bowel resections over time for ulcerative colitis subjects in the Danish cohort. Stratified by faecal calprotectin cluster assignment.

**Figure S25.**
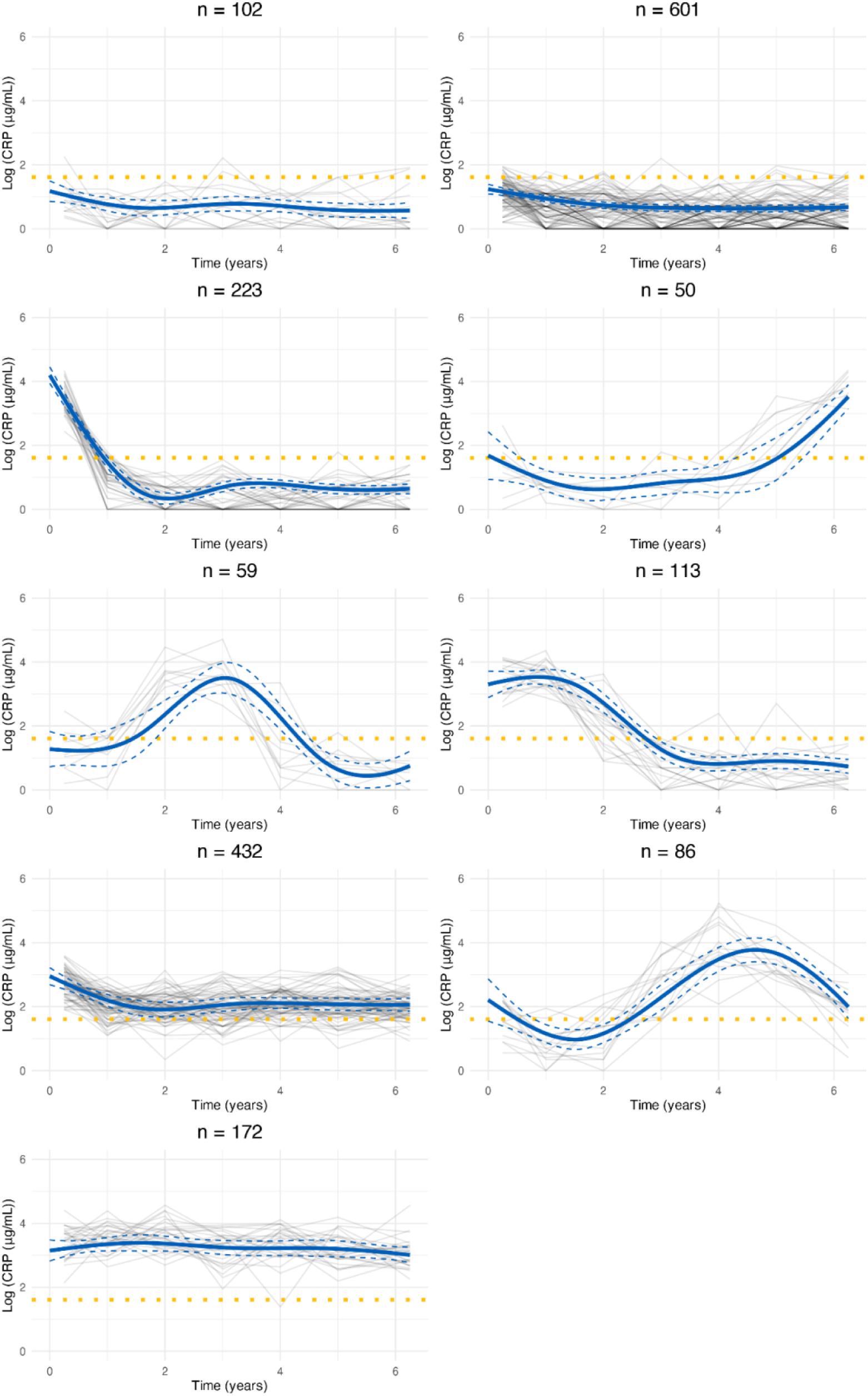
Cluster trajectories obtained from LCMM assuming nine clusters fitted to C-reactive protein data in the Lothian IBD Registry. Red lines indicate predicted mean cluster profiles with 95% confidence intervals. Dotted horizontal lines indicate log(5μg/mL). For visualisation purposes, pseudo subject-specific trajectories have been generated by amalgamating observations from groups of six subjects.

**Figure S26.**
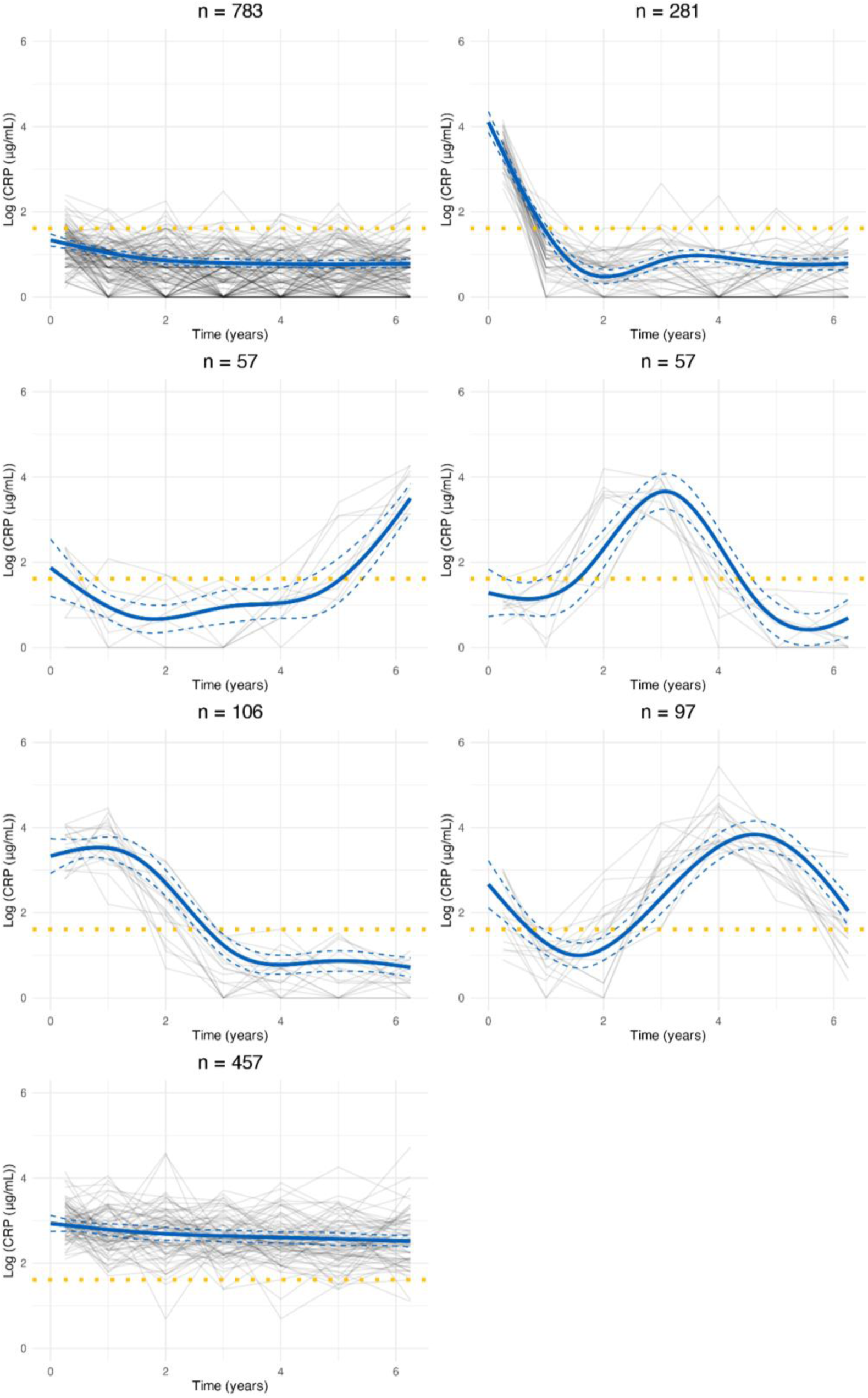
Cluster trajectories obtained from LCMM assuming seven clusters fitted to C-reactive protein data in the Lothian IBD Registry. Red lines indicate predicted mean cluster profiles with 95% confidence intervals. Dotted horizontal lines indicate log(5μg/mL). For visualisation purposes, pseudo subject-specific trajectories have been generated by amalgamating observations from groups of six subjects.

**Figure S27.**
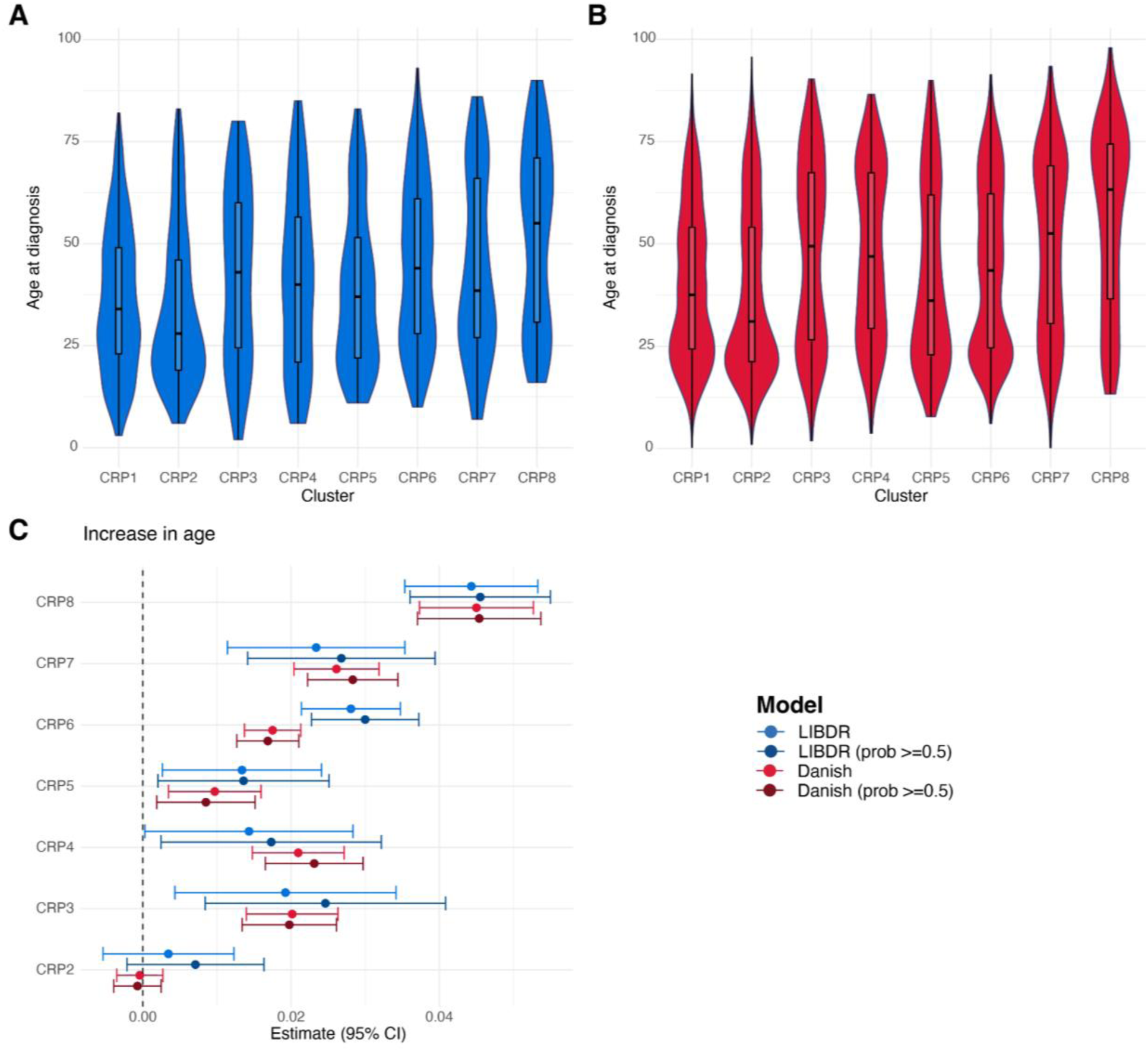
For each CRP cluster, violin plots show the distribution of age at diagnosis across subjects, highlighting median and interquartile ranges for (A) the Lothian IBD Registry and (B) Danish national registry data. (C) Forest plot showing the estimated effect sizes and associated 95% confidence intervals for age in a multinomial logistic regression model that uses CRP cluster assignment as outcome. Subjects with a posterior probability of belonging to their assigned cluster greater than 0.5 were considered in addition to all subjects in the cohort. Effect sizes are with respect to the reference cluster (in this case CRP1). The multivariate model includes age, sex and IBD type as covariates. The dashed vertical lines are used as a reference to indicate no effect.

**Figure S28.**
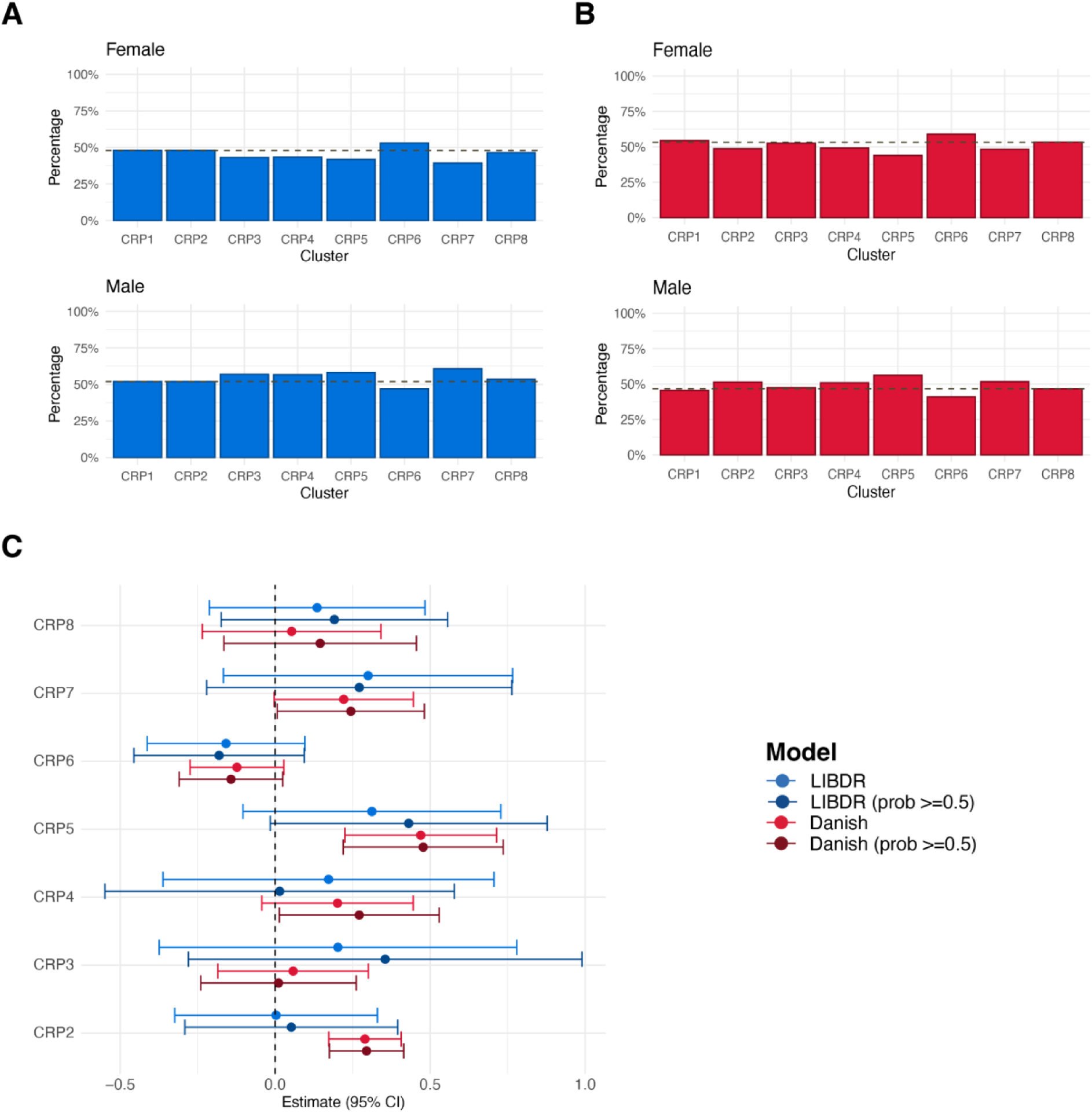
For each CRP cluster, barplots show the proportion of individuals with female and male sex. The dashed horizontal line represents overall proportions for (A) the Lothian IBD Registry and (B) Danish national registry data. (C) Forest plot showing the estimated effect sizes and associated 95% confidence intervals for male sex versus females (baseline category) in a multinomial logistic regression model that uses CRP cluster assignment as outcome. Subjects with a posterior probability of belonging to their assigned cluster greater than 0.5 were considered in addition to all subjects in the cohort. In (C), effect sizes are with respect to the reference cluster (in this case CRP1). The multivariate model includes age, sex and IBD type as covariates. The dashed vertical line is used as a reference to indicate no effect.

**Figure S29.**
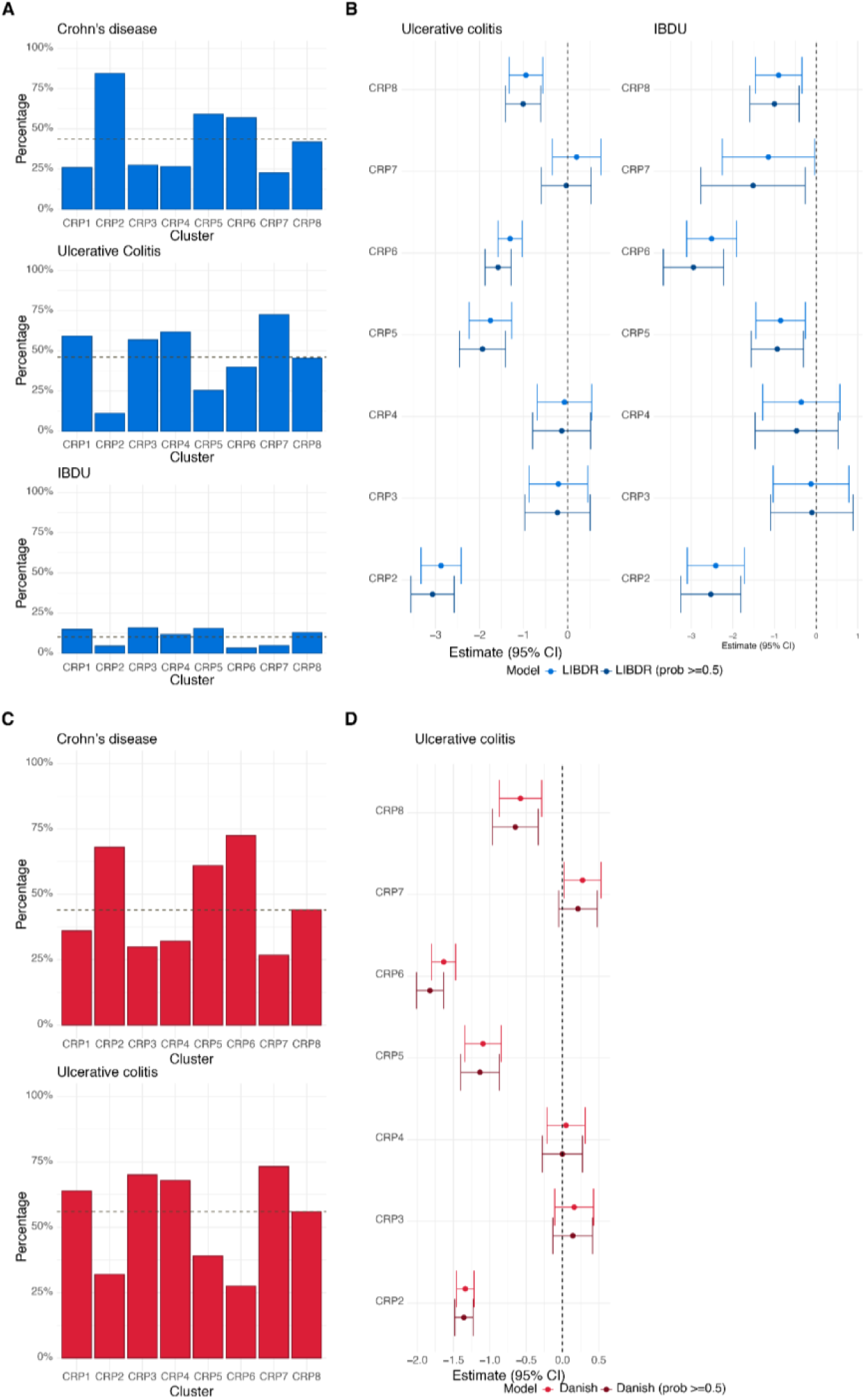
(A) For each CRP cluster, panels show the proportion of individuals with Crohn’s disease, ulcerative colitis and IBDU respectively. The dashed horizontal line represents overall proportions across the entire CRP cohort. (B) Forest plot showing the estimated effect sizes and associated 95% confidence intervals for IBD type: ulcerative colitis and IBDU versus Crohn’s disease (baseline category) in a multinomial logistic regression model that uses CRP cluster assignment as outcome. (C) For each FC cluster, panels show the proportion of individuals with Crohn’s disease and ulcerative colitis respectively in the Danish cohort. (D) Forest plot showing the estimated effect sizes and associated 95% confidence intervals for IBD type in the Danish cohort: ulcerative colitis versus Crohn’s disease (baseline category) in a multinomial logistic regression model that uses FC cluster assignment as outcome. There are no IBDU diagnoses reported for the Danish cohort as these were mapped to either CD or UC (Supplementary Note 1). Subjects with a posterior probability of belonging to their assigned cluster greater than 0.5 were considered in addition to all subjects in the cohort. Effect sizes are with respect to the reference cluster (in this case CRP1). In both cases, the multivariate model includes age, sex and IBD type as covariates. The dashed vertical lines are used as a reference to indicate no effect.

**Figure S30.**
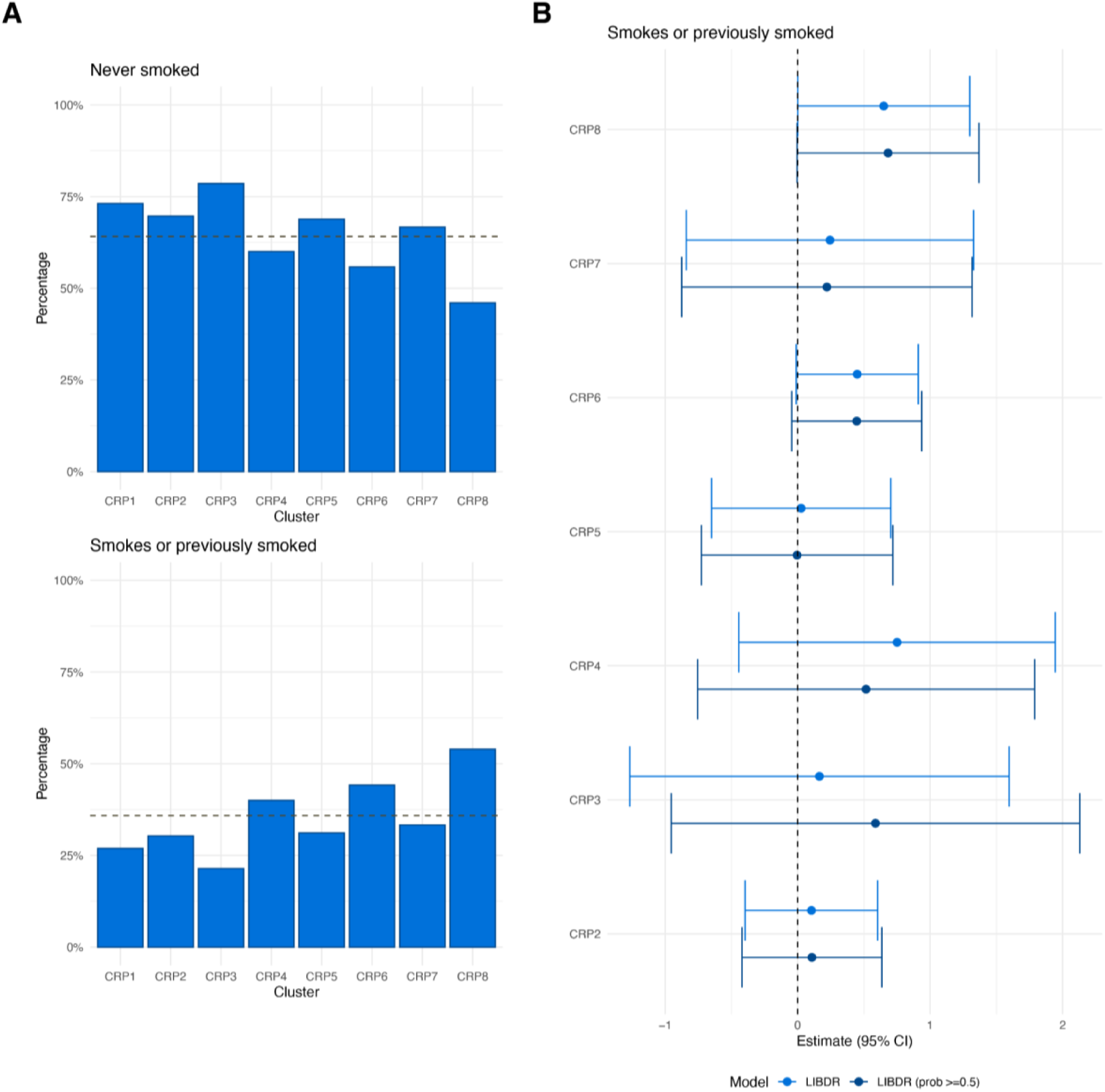
Crohn’s disease patients in the Lothian IBD Registry only. (A) For each CRP cluster, panels show the proportion of individuals with smoking behaviour recorded as “no” (no and never) and “yes” (current or previously smoked) at diagnosis respectively. The dashed horizontal line represents overall proportions across the entire CRP cohort. (B) Forest plot showing the estimated effect sizes and associated 95% confidence intervals for smoking: yes versus no (baseline category) in a multinomial logistic regression model that uses CRP cluster assignment as outcome. Subjects with a posterior probability of belonging to their assigned cluster greater than 0.5 were considered in addition to all subjects in the cohort. Effect sizes are with respect to the reference cluster (in this case CRP1). The multivariate model includes age, sex, smoking, Montreal location (L1, L2, L3), upper gastrointestinal inflammation (L4), and Montreal behaviour (B1, B2/B3) as covariates. The dashed vertical lines are used as a reference to indicate no effect.

**Figure S31.**
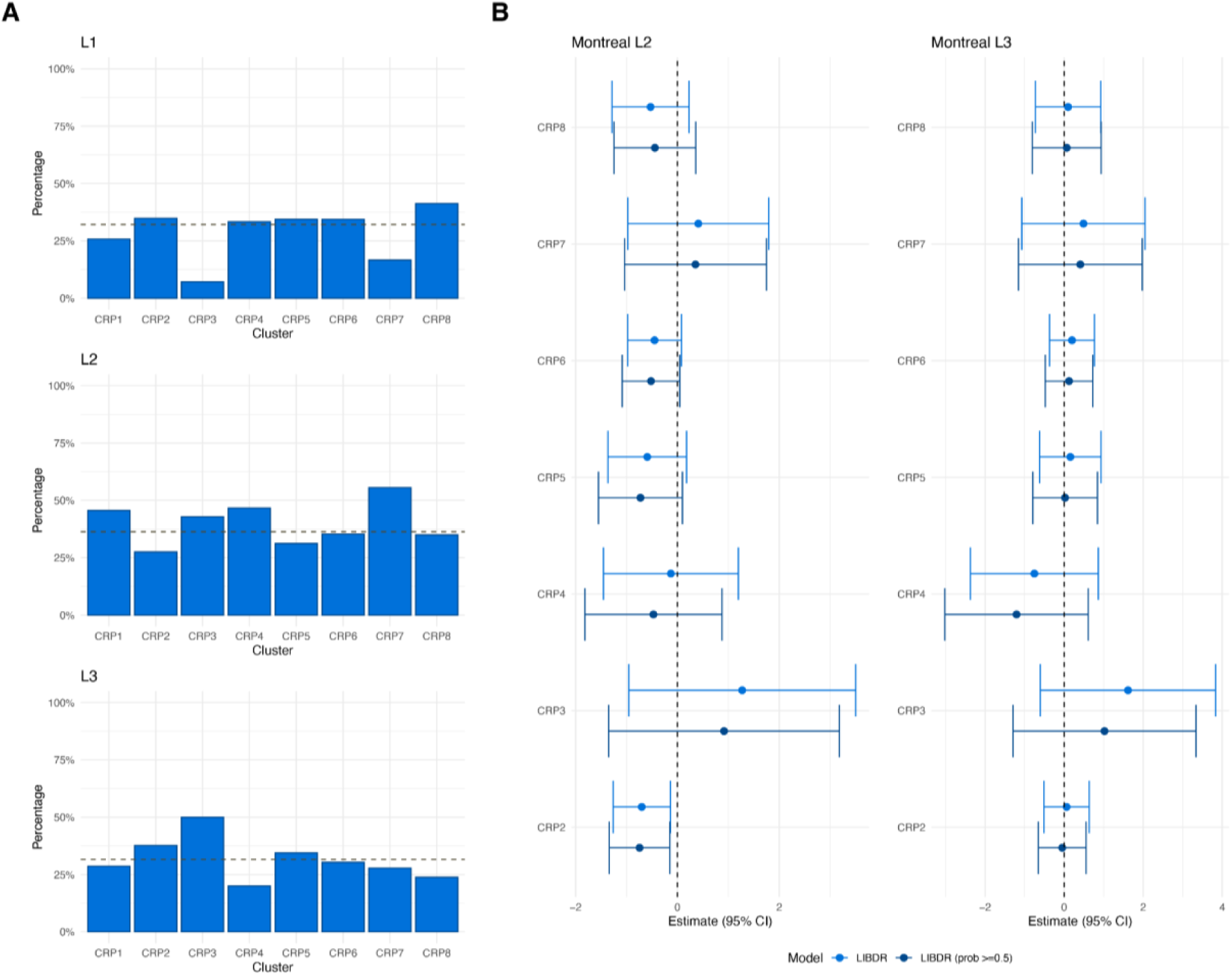
Crohn’s disease patients in the Lothian IBD Registry only. (A) For each CRP cluster, panels show the proportion of individuals with Montreal Location recorded as L1, L2 and L3 respectively. The dashed horizontal line represents overall proportions across the entire CRP cohort. (B) Forest plot showing the estimated effect sizes and associated 95% confidence intervals for Montreal Location: L2 and L3 versus L1 (baseline category) in a multinomial logistic regression model that uses CRP cluster assignment as outcome. Subjects with a posterior probability of belonging to their assigned cluster greater than 0.5 were considered in addition to all subjects in the cohort. Effect sizes are with respect to the reference cluster (in this case CRP1). The multivariate model includes age, sex, smoking, Montreal location (L1, L2, L3), upper gastrointestinal inflammation (L4), and Montreal behaviour (B1, B2/B3) as covariates. The dashed vertical lines are used as a reference to indicate no effect.

**Figure S32.**
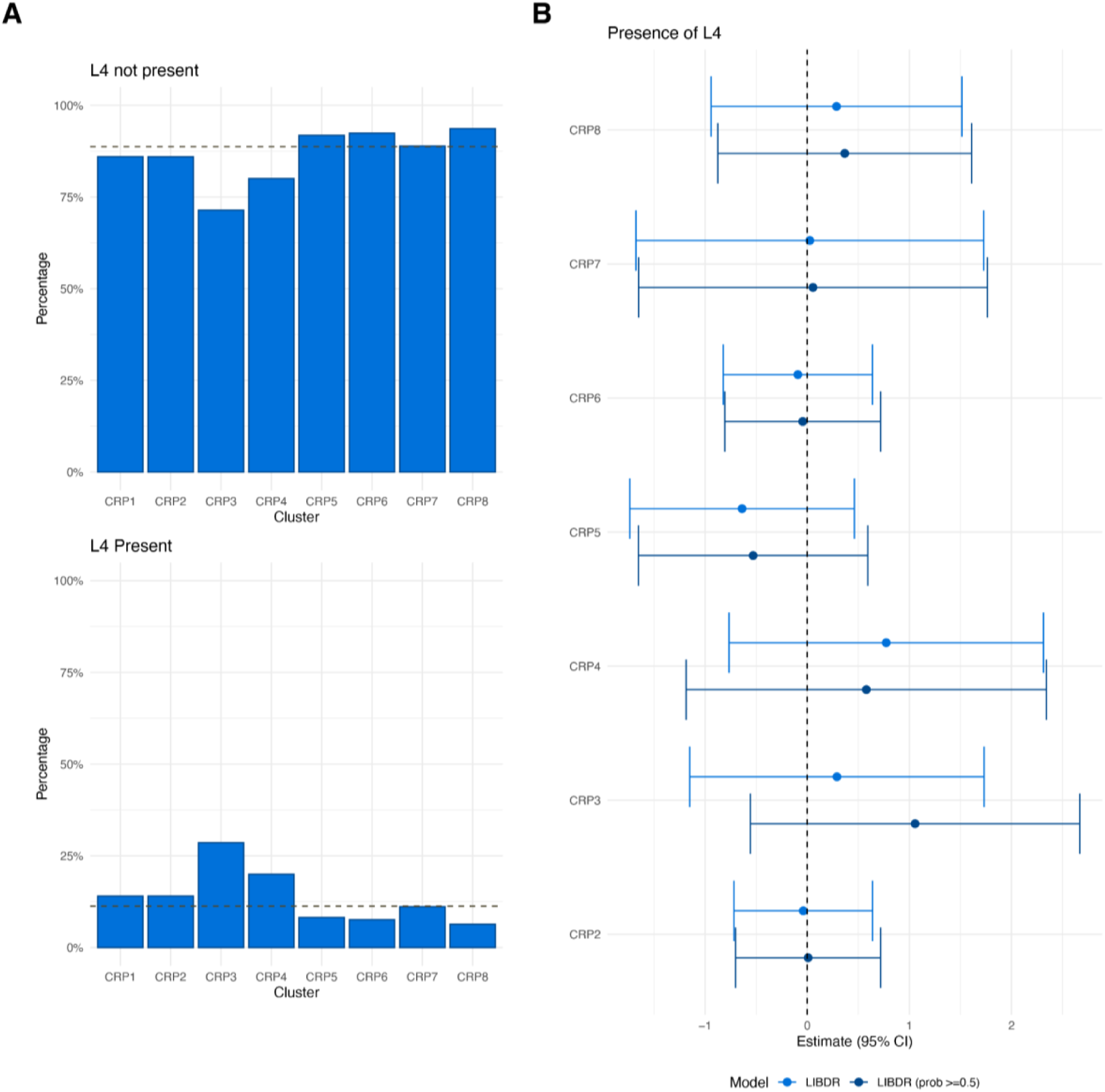
Crohn’s disease patients in the Lothian IBD Registry only. (A) For each CRP cluster, panels show the proportion of individuals with Montreal L4 (upper gastrointestinal inflammation), recorded as present or non present. The dashed horizontal line represents overall proportions across the entire CRP cohort. (B) Forest plot showing the estimated effect sizes and associated 95% confidence intervals for L4: present versus not present (baseline category) in a multinomial logistic regression model that uses CRP cluster assignment as outcome. Subjects with a posterior probability of belonging to their assigned cluster greater than 0.5 were considered in addition to all subjects in the cohort. Effect sizes are with respect to the reference cluster (in this case CRP1). The multivariate model includes age, sex, smoking, Montreal location (L1, L2, L3), upper gastrointestinal inflammation (L4), and Montreal behaviour (B1, B2/B3) as covariates. The dashed vertical lines are used as a reference to indicate no effect.

**Figure S33.**
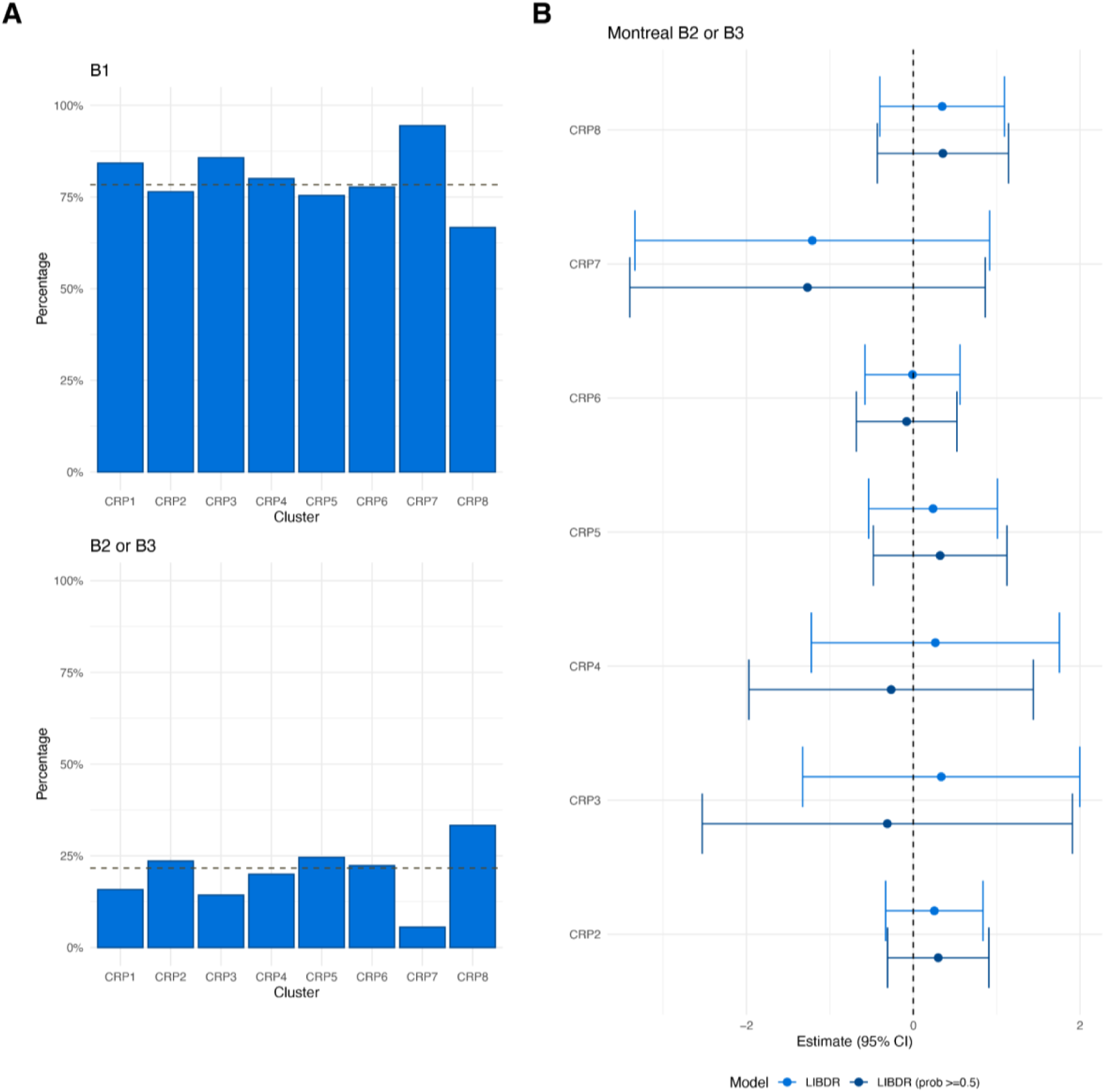
Crohn’s disease patients in the Lothian IBD Registry only. (A) For each CRP cluster, panels show the proportion of individuals with Montreal behaviour recorded as B1 or B2/B3 respectively. The dashed horizontal line represents overall proportions across the entire CRP cohort. (B) Forest plot showing the estimated effect sizes and associated 95% confidence intervals for Montreal behaviour: B2/B3 versus B1 (baseline category) in a multinomial logistic regression model that uses CRP cluster assignment as outcome. Subjects with a posterior probability of belonging to their assigned cluster greater than 0.5 were considered in addition to all subjects in the cohort. Effect sizes are with respect to the reference cluster (in this case CRP1). The multivariate model includes age, sex, smoking, Montreal location (L1, L2, L3), upper gastrointestinal inflammation (L4), and Montreal behaviour (B1, B2/B3) as covariates. The dashed vertical lines are used as a reference to indicate no effect.

**Figure S34.**
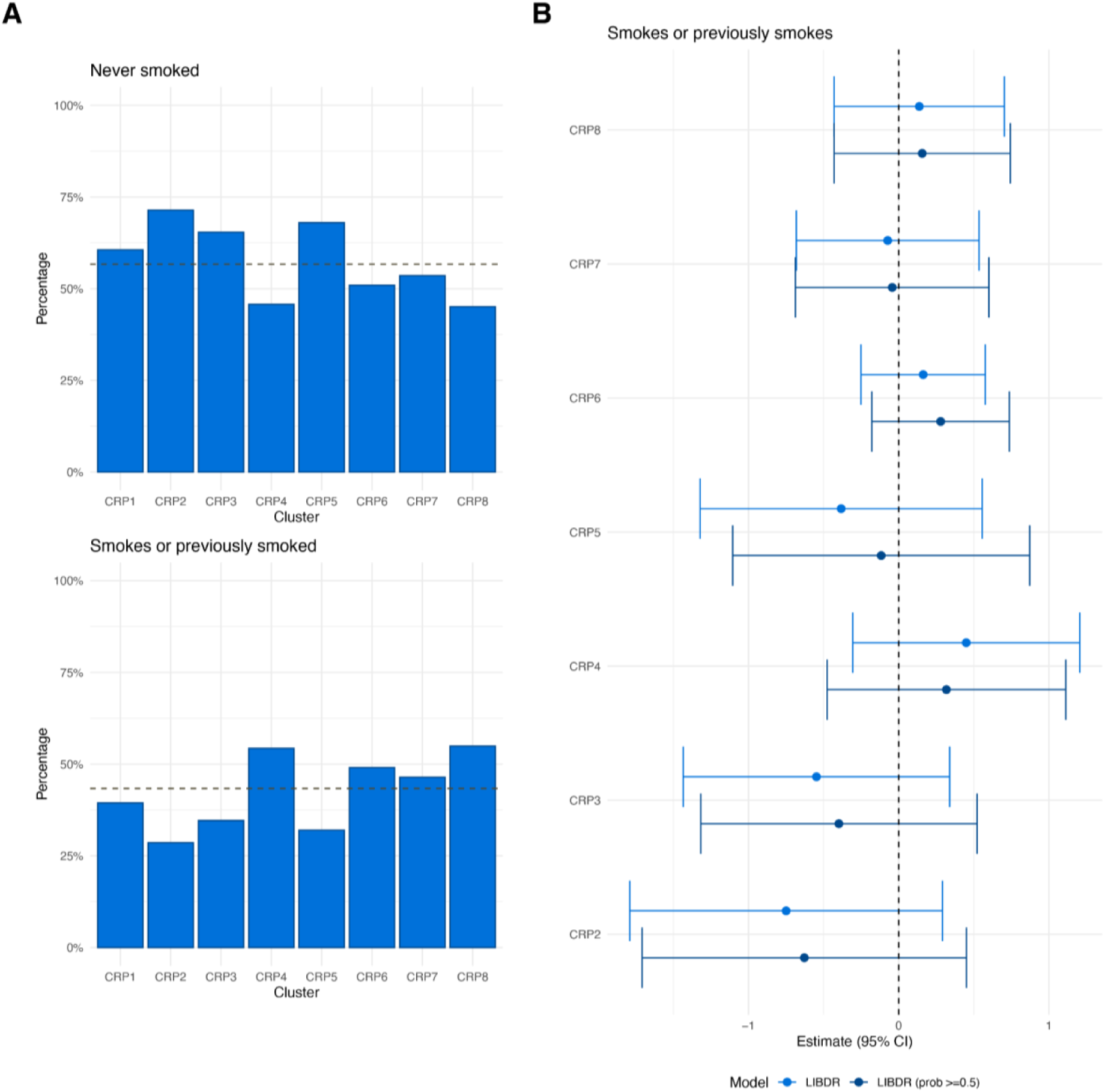
Ulcerative colitis patients in the Lothian IBD Registry only. (A) For each CRP cluster, panels show the proportion of individuals with smoking behaviour recorded as “no” (no and never) and “yes” (current or previously smoked) at diagnosis respectively. The dashed horizontal line represents overall proportions across the entire FC cohort. (B) Forest plot showing the estimated effect sizes and associated 95% confidence intervals for smoking: yes versus no (baseline category) in a multinomial logistic regression model that uses FC cluster assignment as outcome. Subjects with a posterior probability of belonging to their assigned cluster greater than 0.5 were considered in addition to all subjects in the cohort. Effect sizes are with respect to the reference cluster (in this case CRP1). The multivariate model includes age, sex, smoking and Montreal extent (E1, E2, E3). The dashed vertical lines are used as a reference to indicate no effect.

**Figure S35.**
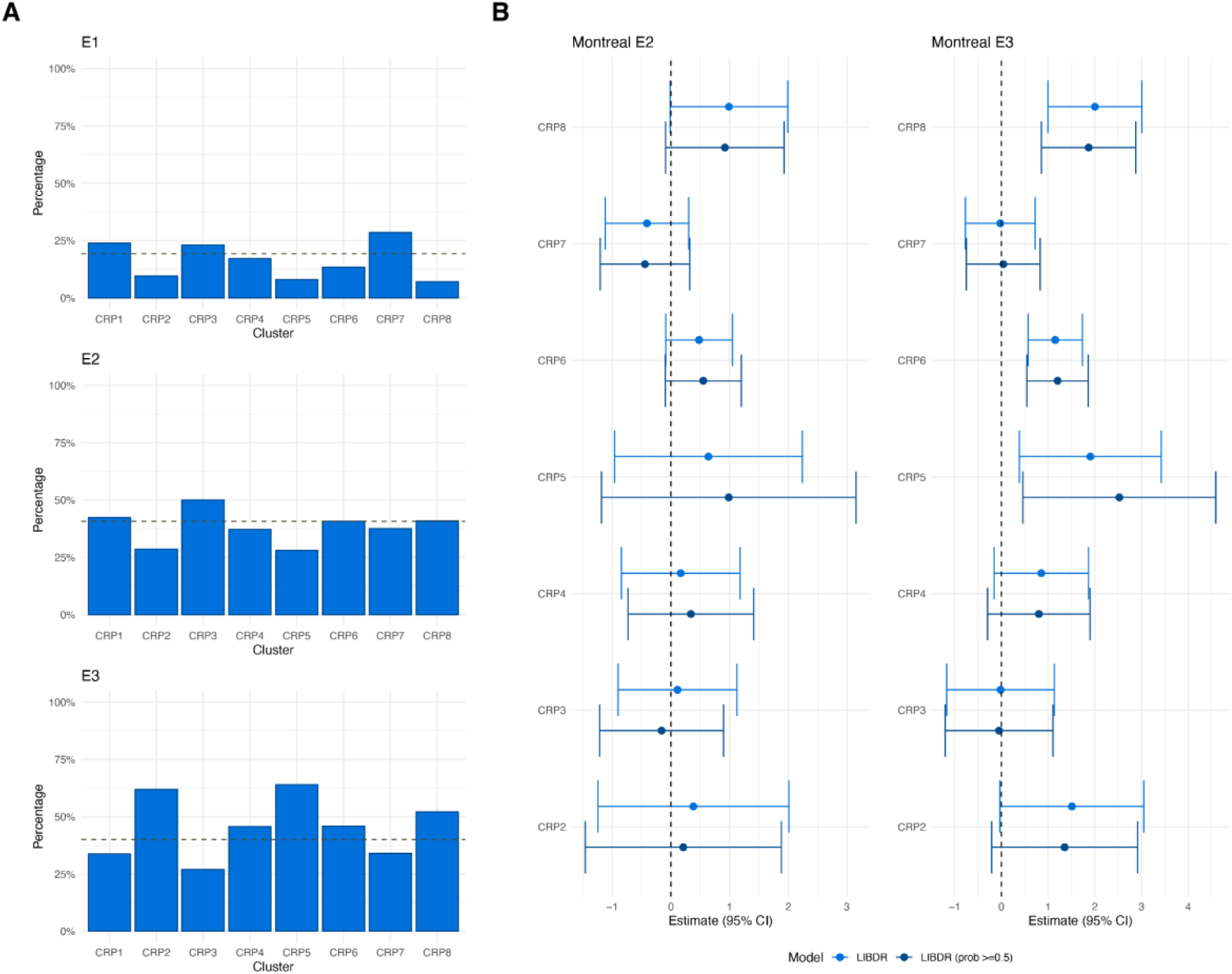
Ulcerative colitis patients in the Lothian IBD Registry only only. (A) For each cluster, panels show the proportion of individuals with Montreal extent recorded as E1, E2 and E3 respectively. The dashed horizontal line represents overall proportions across the entire CRP cohort. (B) Forest plot showing the estimated effect sizes and associated 95% confidence intervals for Montreal extent: E2 and E3 versus E1 (baseline category) in a multinomial logistic regression model that uses FC cluster assignment as outcome. Subjects with a posterior probability of belonging to their assigned cluster greater than 0.5 were considered in addition to all subjects in the cohort. Effect sizes are with respect to the reference cluster (in this case CRP1). The multivariate model includes age, sex, smoking and Montreal extent (E1, E2, E3). The dashed vertical lines are used as a reference to indicate no effect.

**Figure S36.**
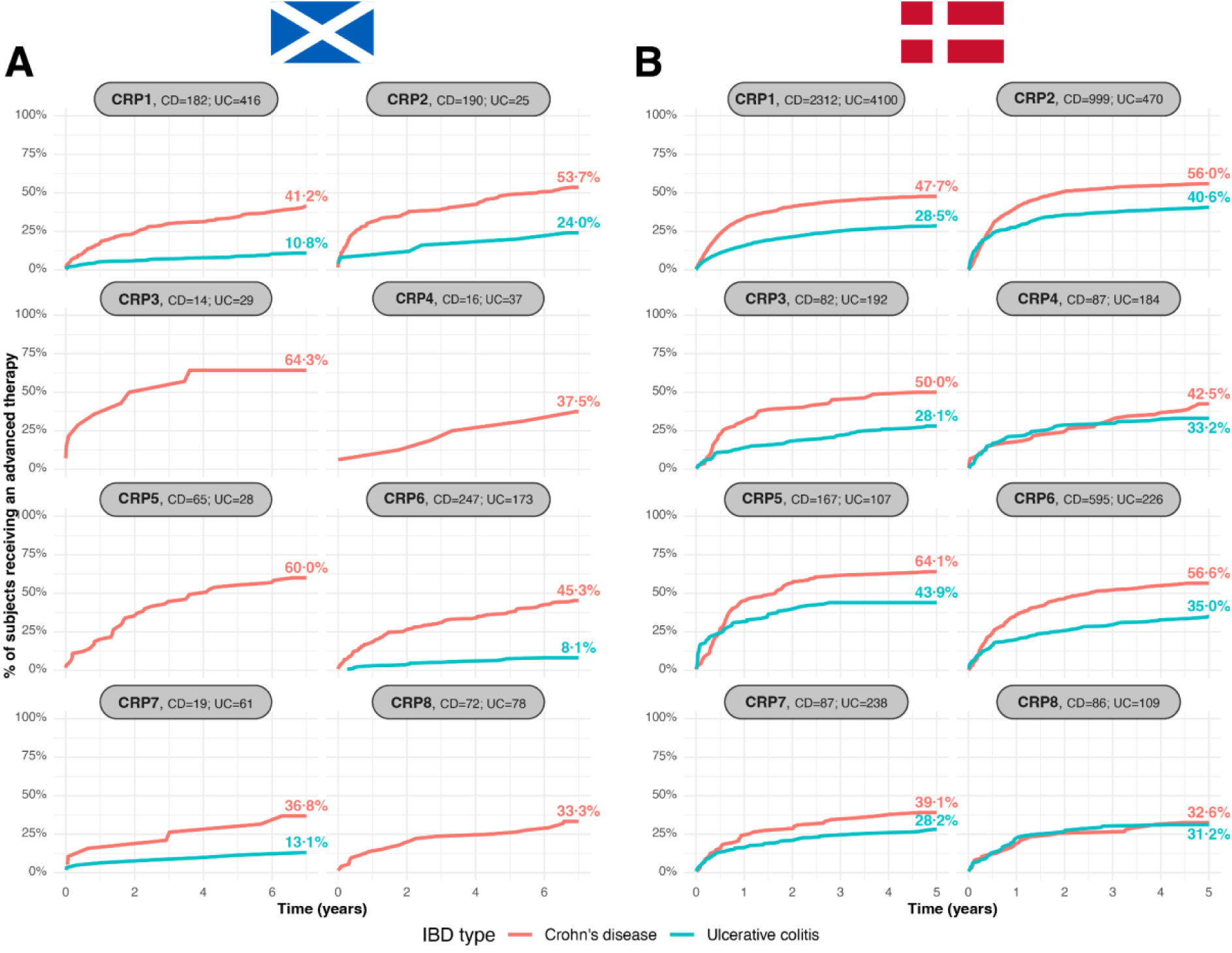
CRP cluster-specific cumulative distribution for first-line advanced therapy prescribing for Crohn’s disease (red) and ulcerative colitis (teal) subjects in (A) the Lothian IBD Registry and (B) national Danish registry data. Clusters are ordered from lowest (CRP1) to highest (CRP8) cumulative inflammatory burden. The number of CD and UC subjects present in each cluster is displayed as panel titles. Curves which would describe fewer than five subjects are not shown.

**Figure S37.**
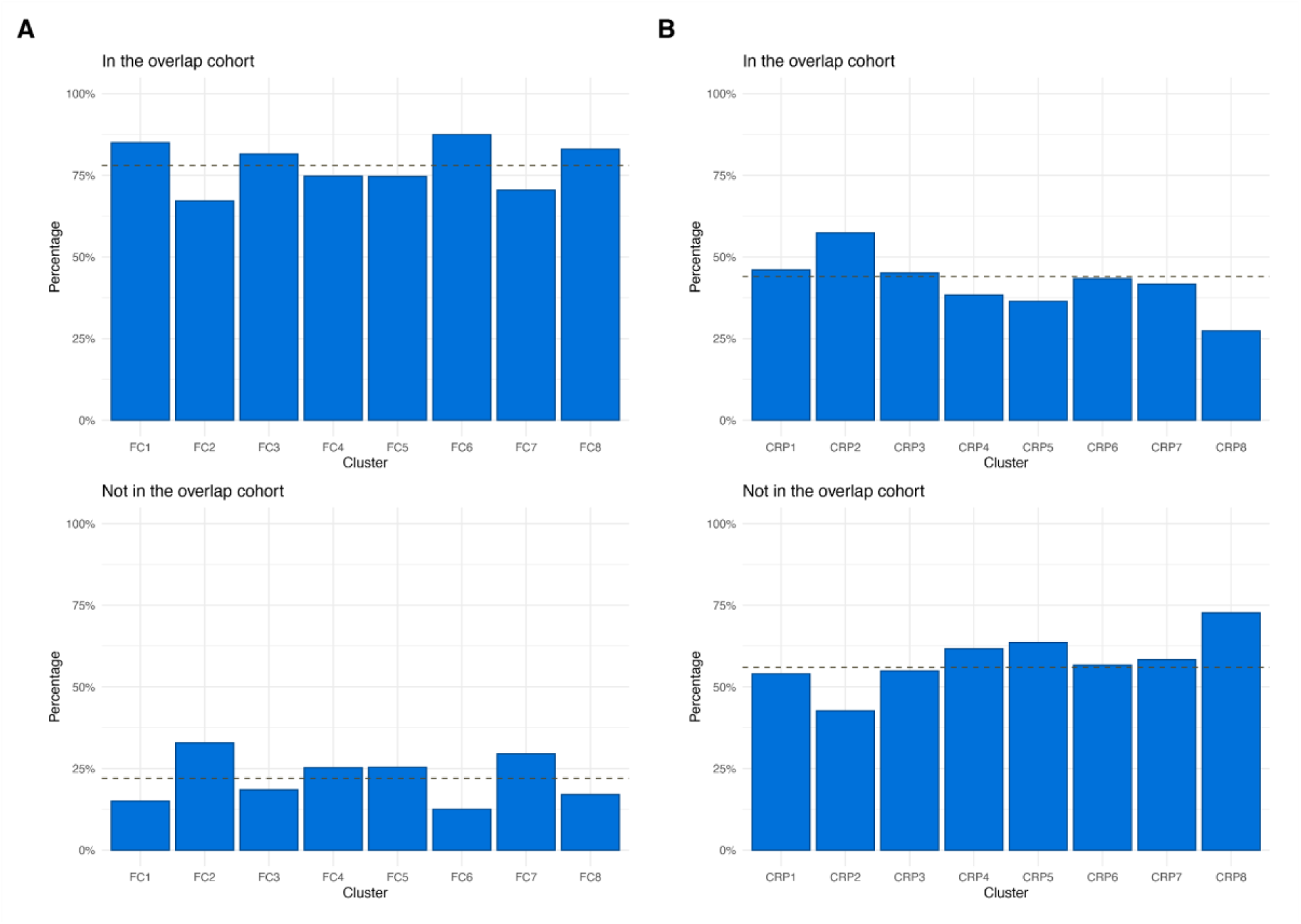
(A) For each FC cluster in the Lothian IBD Registry, panels show the proportion of individuals included in the overlap LIBDR cohort, which consists of LIBDR subjects included in both the FC and CRP analysis. The dashed horizontal line represents overall proportions across the entire FC cohort. (B) As in (A), but focusing on CRP clusters instead. The dashed horizontal line represents overall proportions across the entire CRP cohort.

**Figure S38.**
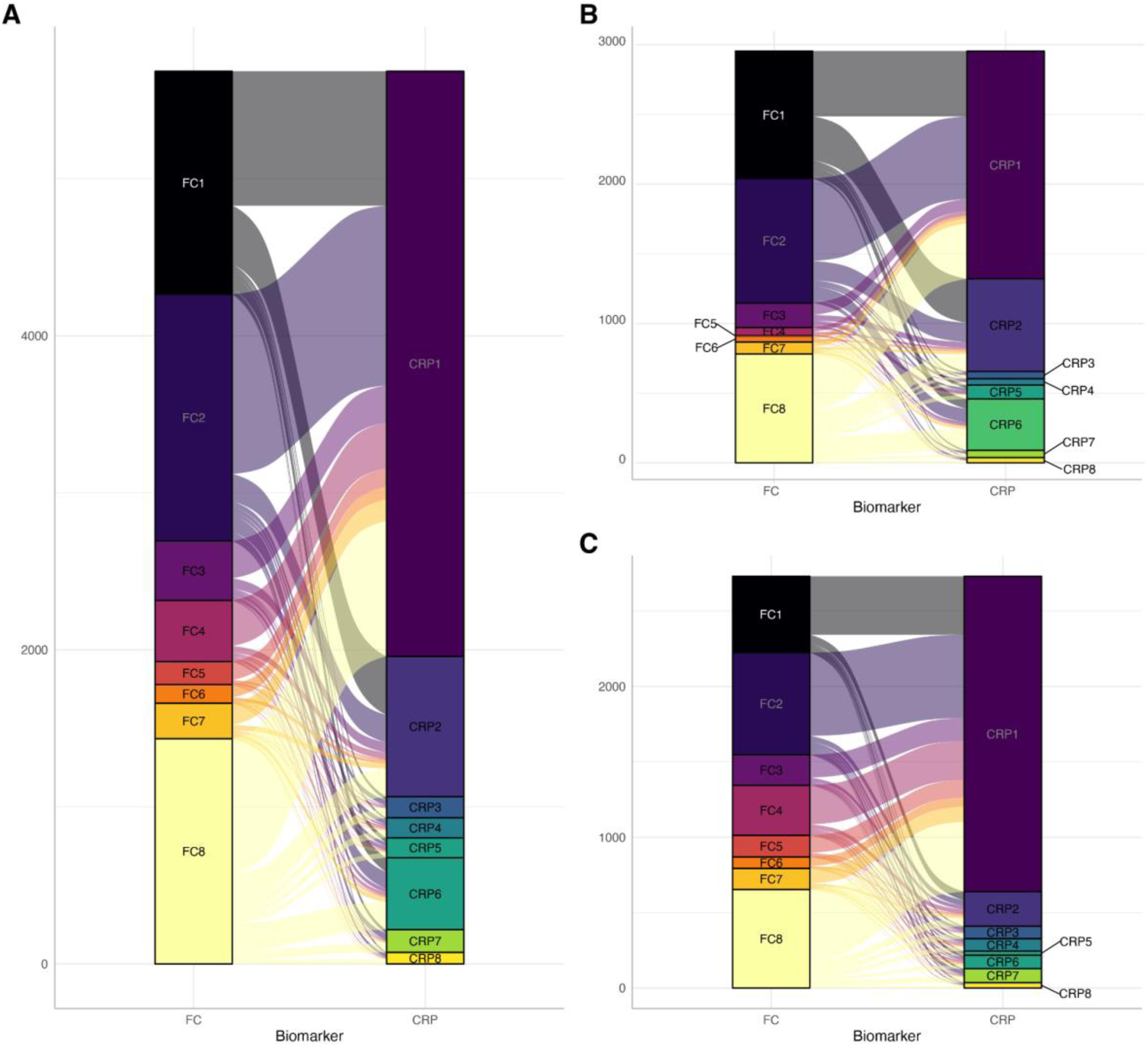
Comparison between faecal calprotectin (FC) and processed CRP cluster assignment for the Danish cohort. Results are reported based on the overlap cohort, consisting of 5685 subjects included in both the FC and CRP analysis. (A) all subjects; (B) Crohn’s disease; and (C) ulcerative colitis. Each segment denotes the size of the cluster whilst the alluvial segments connecting the nodes visualises the number of subjects shared between clusters.

### Supplemental tables

**Table S1.**
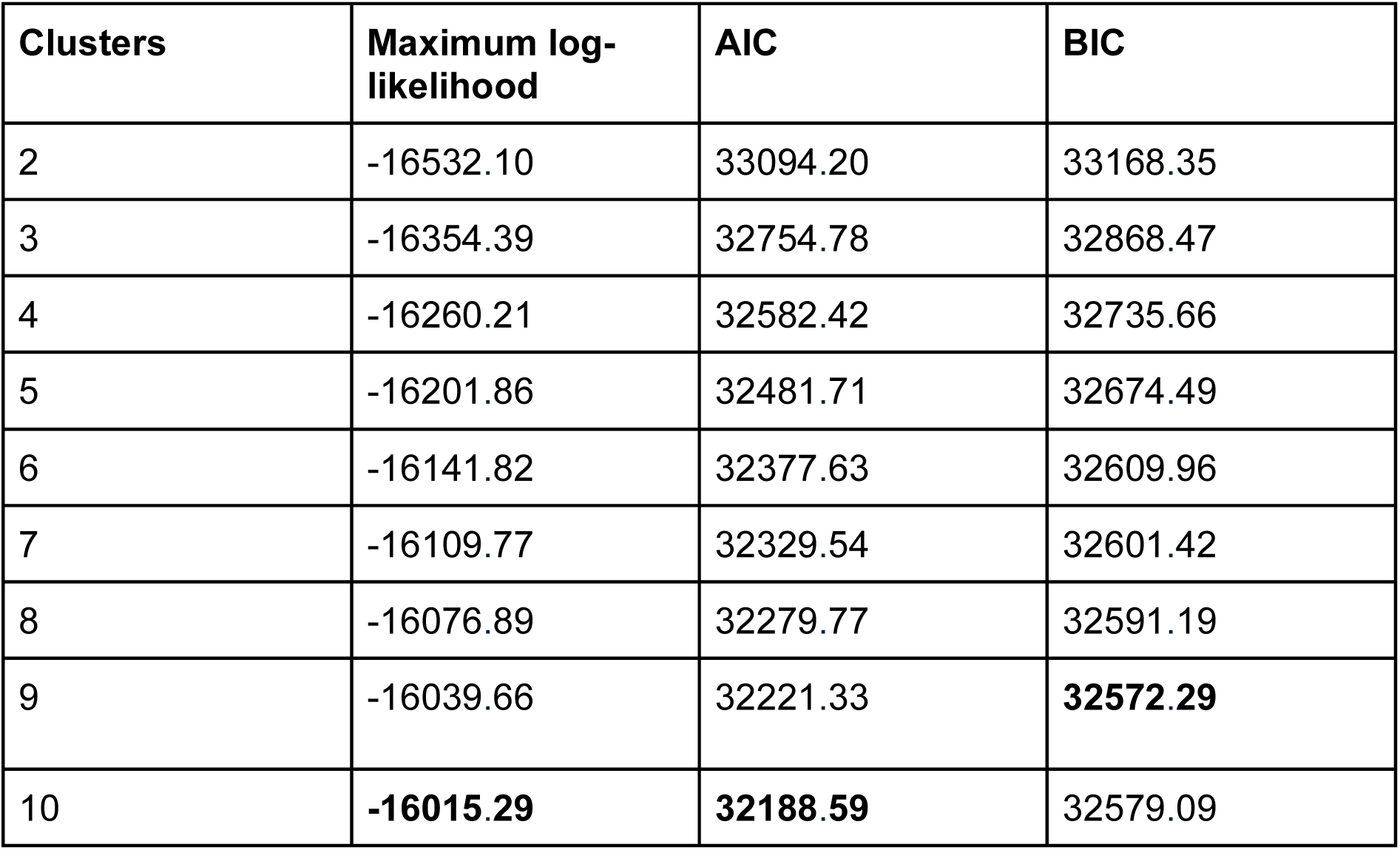
Likelihood-based model statistics for LCMMs fitted to faecal calprotectin data across 2–10 clusters using the chosen model specification. Boldface font indicates optimal values.

**Table S2.** Likelihood-based model statistics for LCMMs fitted to processed CRP data across 2–10 clusters using the chosen model specification. Boldface font indicates optimal values.

| Clusters | Maximum log-likelihood | AIC | BIC |
| --- | --- | --- | --- |
| 2 | -15451.41 | 30932.82 | 31015.57 |
| 3 | -15314.76 | 30675.52 | 30802.40 |
| 4 | -15240.15 | 30542.29 | 30713.30 |
| 5 | -15173.04 | 30424.09 | 30639.23 |
| 6 | -15106.25 | 30306.50 | 30565.78 |
| 7 | -15051.43 | 30212.86 | 30516.27 |
| 8 | -15016.96 | **30159.91** | **30507.45** |
| 9 | -15014.76 | 30171.53 | 30563.20 |
| 10 | **-15014.40** | 30186.81 | 30622.61 |

## Supplementary material for

## Supplemental Note 1: Data definitions

### 1.1 Lothian IBD Registry

#### 1.1.1 Cohort definition

Derivation of the LIBDR cohort was described by Jones et al^1^. In brief, possible IBD cases where first identified from a variety of electronic health record (EHR) resources (e.g. inpatient records with relevant ICD-10 codes). These were subsequently reviewed by a team of eight IBD physicians to exclude non IBD cases. Individuals for which a diagnosis status was unclear were further reviewed by a panel of three IBD physicians. The date of diagnosis, and IBD type for each subject was ascertained as part of the case verification process. This was performed manually and used a priority order of pathology, endoscopy reports and clinic attendance/admission. For some individuals, it was only possible to establish a diagnosis year, without an exact date. In such cases, the date of diagnoses was recorded as January 1st of the corresponding year.

Using capture-recapture sampling, it has been estimated the LIBDR captured 94.3% of all IBD patients treated by NHS Lothian.

Additional information recorded as part of the case verification process includes:

#### 1.1.2 Demographic details

Age and sex for all subjects were obtained as structured data from electronic health records.

#### 1.1.3 Longitudinal biomarker measurements

Longitudinal biomarker data were obtained via an extract from the local biochemistry department. For each observation, test name, test result (including reasons for test failures), sample collection date (when the sample was provided by a subject), and subject identifier were available. The most commonly cited reason for an FC test failing was insufficient sample volume whilst CRP tests most commonly failed due to haemolysis.

Due to limits of detection, the following censoring was used:

- Faecal calprotectin: *<* 20, *>* 1250
- C-reactive protein: *<* 1.

For this study, censored values were mapped to their bounds. For example, *<* 20 was mapped to 20.

#### 1.1.4 Additional phenotyping (at diagnosis)

When available, additional phenotyping information (at diagnosis) was manually extracted by the clinical team from electronic healthcare records (TrakCare; InterSystems, Cambridge, MA). For CD subjects, the following information were extracted:

- **Smoking**: recorded as a binary (yes/no) variable. A high degree of missingness is observed for these data due to smoking status rarely being recorded in electronic health records.
- **Montreal location**:

**–** L1 (Ileal): limited to the ileum, the final segment of the small intestine.
**–** L2 (Colonic): limited to the colon/large intestine.
**–** L3 (Ileocolonic): inflammation is present in both the ileum and colon.

As all Crohn’s disease patients are expected to undergo endoscopic examination at diagnosis, missingness for Montreal location is low.

- Montreal behaviour:

**–** B1 (Inflammatory): non-stricturing and non-penetrating
**–** B2 (Stricturing): where the formation of fibrosis leads to the narrowing of the intestine.
**–** B3 (Penetrating): where the inflammation causes the formation of fistulas or abscesses.

Due to small numbers, B2 and B3 are merged into a single group (complicated CD) when analysing Montreal behaviour. As with Montreal Location, missingness is low due to endoscopic examinations at diagnosis.

- **Upper GI inflammation (L4)**: whether any gastrointestinal inflammation is detected further up than the ileum. Usually, upper inflammation is considered a *modifier* for Montreal location. Recorded as a binary (yes/no) variable. L4 was missing for a high proportion of subjects. This is because the required investigations are only carried out where upper GI inflammation is suspected. As such, we have manually mapped missing L4 values as “No” (i.e. no upper GI inflammation for the associated patients).
- **Perianal disease**: considered a modifier for Montreal behaviour and is a severe complication of Crohn’s disease involving inflammation around the anus. Recorded as a binary (yes/no) variable.

For UC subjects, the following additional phenotyping information was considered:

- **Smoking**: recorded as a binary (yes/no) variable. As with smoking status in Crohn’s disease, a high degree of missingness is observed.
- **Montreal extent**:

**–** E1 (Ulcerative proctitis): limited to the rectum.
**–** E2 (left sided colitis): inflammation is present in the rectum, the sigmoid colon, and possibly the descending colon.
**–** E3 (extensive colitis): inflammation extends beyond the splenic flexure.

Similar to Crohn’s disease, UC patients will undergo endoscopic examination at diagnosis and Montreal extent is rarely missing.

#### 1.1.5 Prescribing and surgery data

The following biologics and small molecules were considered to be advanced therapies relevant for the treatment of IBD. All other advanced therapies were ignored. If one of these advanced therapies were prescribed for a condition other than IBD, for example for rheumatoid arthritis, then these treatments were still considered given they are known to modify IBD outcomes. Only prescriptions given within the observation period (seven years since diagnosis) were considered.

- Adalimumab
- Certolizumab
- Golimumab
- Infliximab
- Risankizumab
- Tofacitinib
- Upadacitinib
- Ustekinumab
- Vedolizumab

Surgery data were not available for this cohort.

### 1.2 National Danish registry data

#### Cohort definition

Agrawal et al. have previously described the derivation of IBD patients from national Danish registry data ^2^. All Danish residents are assigned a personal identification number which can be used to link interactions across the healthcare system. This was used to identify IBD cases. Subjects were also required to have lived in Denmark for at least two years prior to their reported date of diagnosis. Subjects with IBD were defined as either:

1. Having two different secondary care interactions linked to an ICD-10 IBD code (see Table 1.2) within a two year period, and/or
2. Two inpatient interactions assigned an IBD code (no period requirement).

If a subject meets only the first criterion, the first IBD secondary care interaction is used as the date of diagnosis. If a subject only meets the second criterion, then the date of the first inpatient record is used. If a patient meets both criteria, the earliest date is used.

Patients were assigned an IBD subtype (either CD or UC) based on the majority of the ICD-10 codes. If there were an equal number of IBD subtype diagnoses, the latest diagnosis was used.

Table 1.2 details the registry and codes used to obtain the data for each of the variables used in the Danish component of the study.

#### Demographic details

Age and sex for subjects were readily available.

#### Longitudinal biomarker measurements

Measurements of CRP and FC were extracted from the nationwide Register of Laboratory Results for Research (RLRR). The RLRR includes data from 2008, but the database was not nationwide until 2015. We thus only consider patients diagnosed from the 1st of January 2015 and have data available until 1st of December 2022.

Due to limits of detection, the following censoring was used:

- Faecal calprotectin: *>* 1800
- C-reactive protein: *<* 4.

As with the LIBDR, censored values were mapped to their bounds.

FC and CRP were identified via NPU (Nomenclature, Properties and Units) coding ^3^. FC measurements were accessed using the NPU codes NPU19717 and NPU26814. The two NPU codes appeared compatible and were merged. CRP measurements were accessed using the NPU code NPU19748.

#### Additional phenotyping

Montreal classification was not available and smoking status was determined to be missing in too many subjects to be appropiate for inclusion in this study.

#### Prescribing and surgery data

As additional prescribing data were available for the Danish cohort, plots exploring these data were produced. Advanced therapy and immunomodulator plots were generated as the cumulative number of unique treatments over time for each patient. For systemic corticosteroids, one treatment course was defined as 1500 mg with the cumulative number of treatment courses plotted. For major surgeries related to IBD, the cumulative number of surgeries were plotted. For confidentiality reasons, categories with *<* 5 individuals in at each time interval were merged with the category with one fewer treatment.

**Table 1.1:** For each variable used in the Danish analyses, the corresponding registry and codes used to obtain the data.

| Variable | Definition and codes | Source |
| --- | --- | --- |
| **IBD-related hospital interactions** | ICD-10: K50, K51 | Danish National Patient Registry |
| **Biochemistry tests** |  | The Danish nationwide Register of Laboratory Results for Research |
| CRP | NPU: NPU19748 |  |
| Fecal calprotectin | NPU: NPU19717, NPU26814 |  |
| **Biologic therapies and small molecules** |  | Danish National Prescription Registry |
| Infliximab | C_OPR: BOHJ18A1/<br>BOHJ18A |  |
| Adalimumab | C_OPR: BOHJ18A3 |  |
| Ustekinumab | C_OPR: BOHJ18B3 |  |
| Vedolizumab | C_OPR: BOHJ19H4 |  |
| Natalizumab | C_OPR: BOHJ26 |  |
| Certolizumab | C_OPR: BOHJ18A5 |  |
| Golimumab | C_OPR: BOHJ18A4 |  |
| Risankizumab | C_OPR: BOHJ19N1 |  |
| Tofacitinib | C_OPR: BOHJ28D |  |
| Upadacitinib | C_OPR: BWHP107 |  |
| **Immunomodulators** |  | Danish National Prescription Registry |
| Azathioprine | ATC: L04AX01; C_OPR: BWHB83 |  |
| Mercaptopurine | ATC: L01BB02 |  |
| Methotrexate | ATC: L04AX03/L01BA01;<br>C_OPR: BWHA115 |  |
| **Systemic corticosteroids** | ATC: H02AB01, H02AB02, H02AB04, H02AB06, H02AB07, H02AB08, H02AB09 | Danish National Prescription Registry |
| **IBD-related major surgery** |  | Danish National Patient Registry |
| Intestinal resections | NCSP: KJGB, KJFB (excl. KJFB10+13) |  |
| Enteroenterostomy | NCSP: KJFC |  |
| Enterostomy | NCSP: KJFF |  |
| Colectomy | NCSP: KJFH |  |
| Intestinal stricture-plasty | NCSP: KJFA60, KJFA61, KJFA63 |  |
| Other local intestinal surgery | NCSP: KJFA96, KJFA97 |  |
| Other intestinal surgery | NCSP: KJFW |  |
| Stenosis surgery without resection or adhesiolysis | NCSP: KJFL |  |

## Supplemental Note 2: Extended methods

### 2.1 Processing of CRP measurements

When inflammatory cytokines, notably interleukin-6 and tumour necrosis factor-*α*, are produced, CRP is secreted into plasma ^4^. CRP can therefore be viewed as a marker of non-specific inflammation which can be driven by non-IBD related factors such as infection ^5^. As the aim of our analysis is to characterise longitudinal IBD behaviour, we decided to smooth out short-term CRP fluctuations. This smoothing was not required for FCAL, as it is more specific for detecting inflammation at a mucosal level.

For the LIBDR data, CRP measurements (after log transform) were grouped into intervals of *t*: [0, 0.5), [0·5, 1.5), [1.5, 2·5), [2·5, 3·5), [3·5, 4·5), [4·5, 5·5), [5·5, 7], where t = 0 (years) is the time of diagnosis. The median CRP for each interval was calculated for each subject and used as input for subsequent analyses. The centre of each interval was used as the corresponding observation time. A similar approach was used for the Danish data, however intervals [0, 0·5), [0·5, 1·5), [1·5, 2.5), [2·5, 3·5), [3·5, 5] were used instead to take into account the differences in follow-up between the cohorts.

The smoothing process described above was also helpful to reduce computational requirements of LCMM (Section 2.2). Indeed, initial LCMM models run on unprocessed CRP data required very long running times (over a week using 25 CPU cores), complicating the use of a grid search approach to determine the optimal number of clusters (Section 2.3). Furthermore, we observed convergence issues. Model convergence requires measures based on parameter stability, log-likelihood stability, and size of the derivatives to be below a set threshold. The default thresholds with the full Hessian matrix were used.

### 2.2 Latent class mixed models

This note provides a formal definition of latent class mixed models (LCMMs) ^6^ and describes how the most appropriate model specifications were found.

All models were fit using the lcmm R package ^7^. In all cases, models were fit using a maximum of 24,000 Marquardt iterations and all other options left at the default settings. An automatic grid search approach with 50 repetitions was used to attempt to avoid local maxima. All models converged.

We assume a population of *N* individuals is heterogeneous and composed of *G* latent classes (or clusters): each characterised by a distinct mean profile of a marker of disease activity (in logarithmic scale) across time. We assume each subject *i* has a vector of repeated measurements of length *n_i_*, allowing the number of measurements to differ across subjects. We allow each subject *i* to belong to only one latent class and introduce a discrete random variable *c_i_* which is equal to *g* if subject *i* belongs to the latent class *g*, where *g* = 1*, …, G*. LCMM consists of two sub-models which are defined below.

#### Longitudinal sub-model

The logarithm of the biomarker measurement (FC or CRP) for the *i*th subject taken at time *t_ij_* is denoted by *Y_ij_*. Given that the subject *i* belongs to class *g*, the latter is modelled as follows:

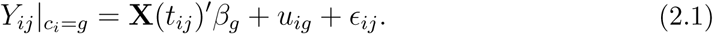

Effectively, this corresponds to a class-specific linear mixed model. In (2.1), the vector of regression coefficients *β_g_* capture class-specific fixed effects. A random effects specification is used to capture intra-individual correlation in measurements. Let *u_ig_* denote random effects distributed such that *u_ig_* ∼ N(0*, σ*^2^) (we assume the variance *σ*^2^ is shared across classes, but a more general form of the model allows for class-specific variance). In our analysis, we assume a single random effect (intercept) *u_ig_*. Finally, *ɛ_ij_* indicates an independently distributed Gaussian error term with zero mean and variance *σ*^2^.

#### Class membership sub-model

The probabilistic approach used by LCMM naturally captures uncertainty in class assignments. The probability of assigning individual *i* to class *g*, i.e. *c_i_* = *g* is defined by a multinomial logistic model:

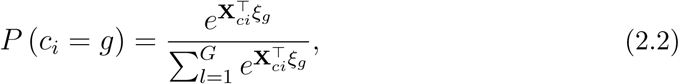

where **X***_ci_* is a vector of covariates and *ξ_g_* is a vector of parameters whose first element corresponds to a class-specific intercept. For identifiability, the intercept associated to class *G* is assumed to be equal to zero. Note that the covariates in Equation (2.2) do not need to match those used in the longitudinal model in (2.1). In our analysis, only IBD type was included as a covariate in this model. This was defined as CD, UC or IBDU for the LIBDR cohort. For the Danish data, this was defined by CD or UC only.

#### Posterior probabilities of class assignment

After inferring all model parameters, posterior class-membership probabilities for each subject are given by:

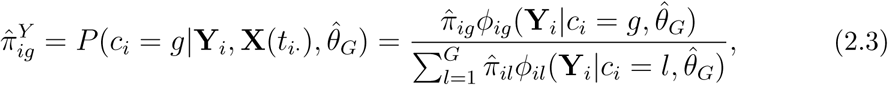

where **Y***_i_* denotes a vector of length *n_i_* containing all longitudinal measurements recorded for subject *i*, **X**(*t_i·_*) is a matrix comprised of all the corresponding time-dependent co-variates for subject *i* (fixed effects), *θ*^^^*_G_* denotes the estimates obtained for all model parameters (*β*_1_*, …, β_G_, σ*^2^*, σ*^2^*, ξ*_1_*, …, ξ_G_*) and *π*^*_ig_* corresponds to Equation (2.2) evaluated on *θ*^^^*_G_*. Finally, *ϕ_ig_*(**Y***_i_*|*c_i_* = *g, θ*^^^*_G_*) denotes a multivariate normal density function with mean **X**(*t_i·_*)*β*^^^*_g_* and variance covariance *σ*^2^**X**(*t_i·_*)**X**(*t_i·_*)*^′^* + *σ*^^2^*I_n_*, where *I_n_* denotes an identify matrix with dimension *n_i_*.

Note that (2.3) considers all the longitudinal measurements available for an individual to assign them to a latent class. As such, cluster allocations are static.

#### Fixed effects specified as natural cubic splines

The vector of time-dependent covariates **X**(*t*) (see Equation (2.1)) was defined using an intercept term and natural cubic splines (NCS) ^8^ to capture non-linear dependency between observations and time. The NCS were calculated as a pre-processing step prior to estimating the model in Equation (2.1) using the ns() function of the splines R library ^9^. Knots were located at quantiles of measurement times across all measurements. To determine the most appropriate number of interior knots for the LIBDR cohort, LCMMs with six clusters were fitted to the LIBDR FC data assuming two to four knots. Visual inspection, Akaike information criterion (AIC), and Bayesian information criterion (BIC) were used to determine the most appropriate NCS specification, which was then used to fit the models used in the main analysis.

The optimal AIC was reported for the NCS model with four interior knots whilst BIC favoured the three-knot specification (Table 2.1). However, visual inspection of the four-knot specification appeared to result in overfitting of the data (Figure 2.1). As such, the three-knot approach was used to specify the fixed effects of the longitudinal sub-model for all further FC and CRP modelling in the LIBDR cohort. As such, the time-dependent covarite vector was defined as **X**(*t*) = (1*, X*_1_(*t*)*, X*_2_(*t*)*, X*_3_(*t*)*, X*_4_(*t*))*^′^* (the first element ensures the model includes an intercept term).

A similar approach was used to define the number of knots when analysing the Danish data. Two knots were chosen for both FC and CRP analyses.

### 2.3 Grid search to define the number of classes ***G***

LCMM requires the number of latent classess *G* to be specified a priori. As *G* is generally unknown, a grid search approach was used. This was applied separately for FCAL and CRP models fitted to the LIBDR dataset (discovery cohort) which has a more precise phenotyping (date of diagnosis, IBD type). The chosen number of clusters was then applied when analysing the Danish data (validation cohort).

Model selection considered the Akaike information criterion (AIC) and the Bayesian information criterion (BIC) ^10^. As previously described by Bauer and Curran^11^, likelihood based statistics such as AIC and BIC have been shown to overestimate the number of ‘true’ clusters when model residuals are not normally distributed. This is a characteristic commonly seen in FC and CRP due to censoring greatly increasing densities at the ends of the distribution. As such, we complemented AIC/BIC-informed model choices with a visual inspection of cluster trajectories. The latter was used to derive parsimonious model choices, ensuring that each cluster captured a distinct longitudinal pattern and to avoid very small clusters (*<*50 individuals) which could be difficult to interpret.

**Table 2.1:** Likelihood-based model statistics for LCMMs assuming six clusters with varying specifications for the fixed effects of the longitudinal sub-model. The models considered used natural cubic splines with varying knots. Boldface font indicates optimal values.

| Specification | Maximum log-likelihood | AIC | BIC |
| --- | --- | --- | --- |
| 2 Knots | -19766.31 | 39614.61 | 39823.41 |
| 3 Knots | -19615.33 | 39324.67 | **39564.02** |
| 4 Knots | **-19604.65** | **39315.30** | 39585.21 |

**Figure 2.1:**
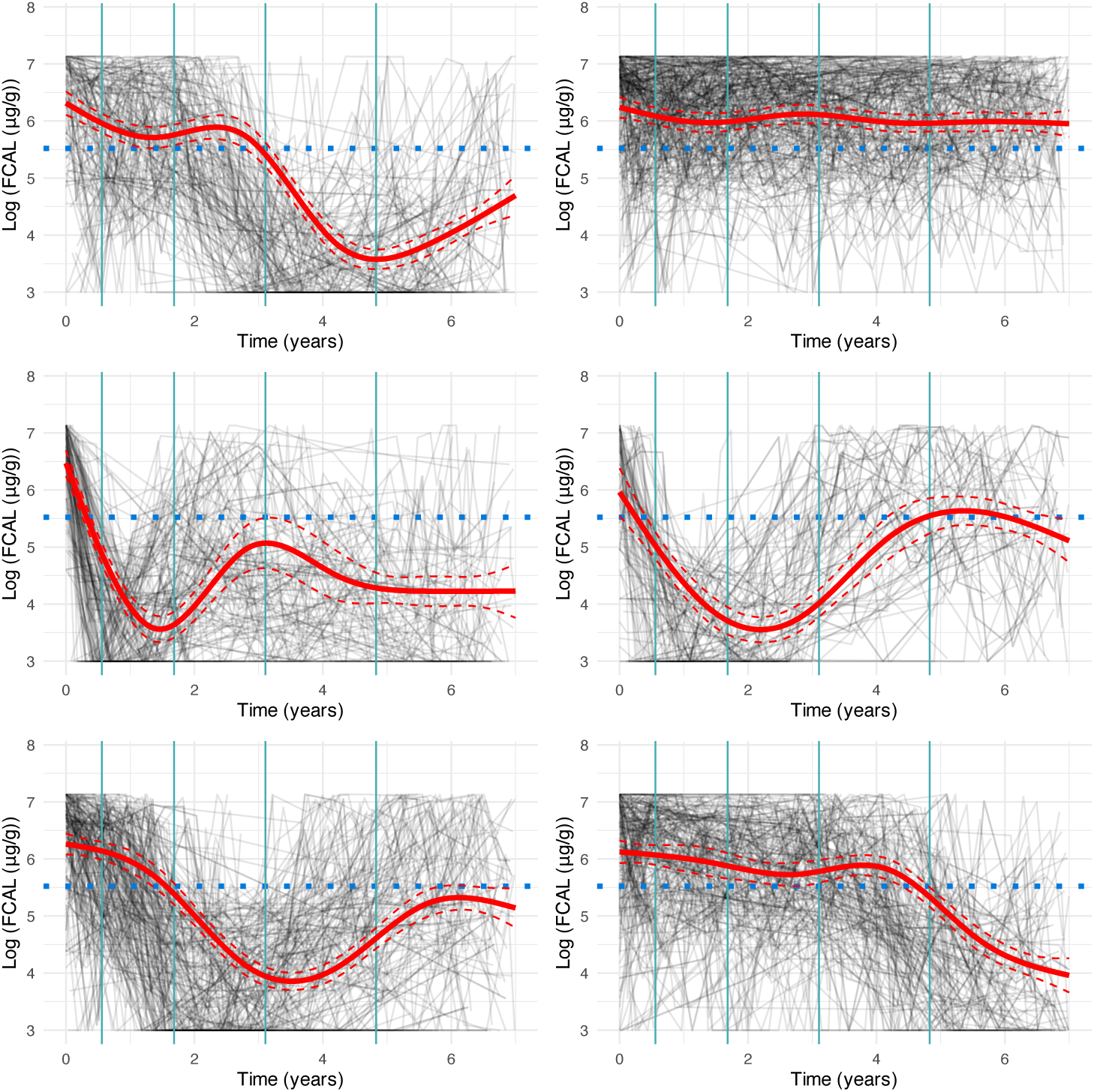
Cluster trajectories of FC for a LCMM using natural cubic splines with 4 knots. Vertical teal lines indicate knot locations. Dotted horizontal lines indicate log(250*µg/g*) FC which is commonly used as a cutoff for disease activity.

### 2.4 Estimating cluster-specific cumulative inflammatory burden

We ordered the clusters found in the LIBDR data according to their associated cumulative inflammatory burden. The latter was calculated based on predicted mean profile for each cluster, using the area under the curve as a proxy for cumulative inflammatory burden. Predicted mean profiles were defined by the fixed effects included in the model defined by Equation 2.1, i.e. **X**(·)*^′^β_g_*, where *β_g_* was replaced by its corresponding maximum likelihood estimate.

The area under the predicted mean profile for each cluster was estimated using the trapezoidal rule with 700 trapezoids placed equidistantly via the trapz R function from the {pracma} package. Let *f* (·) be the function of interest, and *t*_0_*, …, t*_700_ represent a grid of time-points where *t*_0_ and *t*_700_ represent the start and end of the area to be calculated. The area under the curve is estimated using

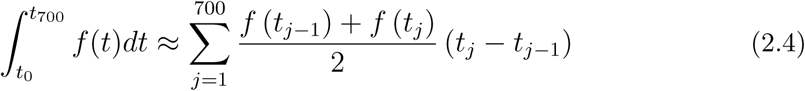

## Supplemental Note 3: Software versions

As LIBDR and Danish analyses were conducted on separate computational platforms, the software versions differed.

### 3.1 Lothian IBD Registry

R (v. 4·4·2), ^9^ extended using the lcmm (v. 2·1·0), ^6^ ggalluvial (v. 0·12·5), ^12^ nnet (v. 7·3-19), ^13^ and datefixR (v.1·7·0) ^14^ packages, was used. Analytical reports have been generated using the Quarto scientific publishing system (v.1.4.551) and are hosted online (https://vallejosgroup.github.io/IBD-Inflammatory-Patterns/). An R package, libdr (v.1·1·0), has also been produced, supporting the reuse of our R code with other datasets.

### 3.2 National Danish registry data

R (v. 4·4·0) extended using the lcmm (2·1·0), ggalluvial 0·12·5, and nnet 7·3·19 packages was used for the Danish analyses. The R code is shared in the VallejosGroup/IBD-Inflammatory-Patterns GitHub repository under the source/dk directory.

## Notes

### Author Declarations

Usage of the Scottish dataset was approved by the local Caldicott Guardian (Project ID: CRD18002, registered NHS Lothian information asset #IAR-954). In Denmark, studies based on registry data alone are not required to obtain permission from the regional ethics committees as confirmed by The Central Denmark Region Committees on Health Research Ethics (legislation: 1-10-72-148-19)

### Summary of Updates

Word count reduced. Minor clarifying edits.

